# Genome-wide study of DNA methylation in Amyotrophic Lateral Sclerosis identifies differentially methylated loci and implicates metabolic, inflammatory and cholesterol pathways

**DOI:** 10.1101/2021.03.12.21253115

**Authors:** Paul J. Hop, Ramona A.J. Zwamborn, Eilis Hannon, Gemma L. Shireby, Marta F. Nabais, Emma M. Walker, Wouter van Rheenen, Joke J.F.A. van Vugt, Annelot M. Dekker, Henk-Jan Westeneng, Gijs H.P. Tazelaar, Kristel R. van Eijk, Matthieu Moisse, Denis Baird, Ahmad Al Khleifat, Alfredo Iacoangeli, Nicola Ticozzi, Antonia Ratti, Jonathan Cooper-Knock, Karen E. Morrison, Pamela J. Shaw, A. Nazli Basak, Adriano Chiò, Andrea Calvo, Cristina Moglia, Antonio Canosa, Maura Brunetti, Maurizio Grassano, Marc Gotkine, Yossef Lerner, Michal Zabari, Patrick Vourc’h, Philippe Corcia, Philippe Couratier, Jesus S. Mora Pardina, Teresa Salas, Patrick Dion, Jay P. Ross, Robert D. Henderson, Susan Mathers, Pamela A. McCombe, Merrilee Needham, Garth Nicholson, Dominic B. Rowe, Roger Pamphlett, Karen A. Mather, Perminder S. Sachdev, Sarah Furlong, Fleur C. Garton, Anjali K. Henders, Tian Lin, Shyuan T. Ngo, Frederik J. Steyn, Leanne Wallace, Kelly L. Williams, BIOS Consortium, Brain MEND Consortium, Miguel Mitne Neto, Ruben J. Cauchi, Ian P. Blair, Matthew C. Kiernan, Vivian Drory, Monica Povedano, Mamede de Carvalho, Susana Pinto, Markus Weber, Guy Rouleau, Vincenzo Silani, John E. Landers, Christopher E. Shaw, Peter M. Andersen, Allan F. McRae, Michael A. van Es, R. Jeroen Pasterkamp, Naomi R. Wray, Russell L. McLaughlin, Orla Hardiman, Kevin P. Kenna, Ellen Tsai, Heiko Runz, Ammar Al-Chalabi, Leonard H. van den Berg, Philip Van Damme, Jonathan Mill, Jan H. Veldink

## Abstract

Amyotrophic lateral sclerosis (ALS) is a fatal neurodegenerative disease with an estimated heritability of around 50%. DNA methylation patterns can serve as biomarkers of (past) exposures and disease progression, as well as providing a potential mechanism that mediates genetic or environmental risk. Here, we present a blood-based epigenome-wide association study (EWAS) meta-analysis in 10,462 samples (7,344 ALS patients and 3,118 controls), representing the largest case-control study of DNA methylation for any disease to date. We identified a total of 45 differentially methylated positions (DMPs) annotated to 42 genes, which are enriched for pathways and traits related to metabolism, cholesterol biosynthesis, and immunity. We show that DNA-methylation-based proxies for HDL-cholesterol, BMI, white blood cell (WBC) proportions and alcohol intake were independently associated with ALS. Integration of these results with our latest GWAS showed that cholesterol biosynthesis was causally related to ALS. Finally, we found that DNA methylation levels at several DMPs and blood cell proportion estimates derived from DNA methylation data, are associated with survival rate in patients, and could represent indicators of underlying disease processes.

## Introduction

Amyotrophic lateral sclerosis (ALS) is a fatal neurodegenerative disorder that is characterized by progressive degeneration of motor neurons in the brain and spinal cord^1^. The disease affects about 1 in 350 people, with death typically occurring within 2 to 5 years after onset. The heritability of ALS is estimated to be around 50%^2^, showing that a considerable portion of the risk could be conferred by environmental and lifestyle risk factors. However, the identification of these factors has proven difficult due to several challenges. Lifestyle factors are typically studied using self reports, or interviews with the risk of recall and selection bias (e.g. because of selection against patients with cognitive impairment)^3,4^. Finally, biomarkers are typically measured after onset of disease making inferences about causality virtually impossible. The combined effect of these challenges has resulted in a large body of literature with conflicting results and only a few established factors related to ALS risk or patient survival^3,5^.

Epigenetic patterns, which regulate gene expression via modifications to DNA, histone proteins, and chromatin, can serve as biomarkers of disease progression and (past) exposures such as smoking and alcohol intake. Moreover, epigenetics patterns can provide a mechanism that mediates genetic and/or environmental risk^6^. The development of standardized assays for quantifying DNA methylation, the best characterized and most stable epigenetic modification, has enabled the systematic analysis of associations between methylomic variation and a wide range of human diseases, including cancer, schizophrenia and various neurodegenerative diseases^6–9^. In addition to a potential mechanistic role in ALS, the quantification of DNA methylation in whole blood DNA can serve as a robust quantifiable biomarker for candidate risk factors including smoking, alcohol intake, BMI, biological age, and various metabolic and inflammatory proteins^10–16^. Moreover, DNA methylation signatures have proven useful as diagnostic and prognostic biomarkers in various settings including common cancers and all-cause mortality^17,18^.

Few studies have investigated the role of DNA methylation in ALS. Most studies have either focused on specific genomic regions such as the *C9orf72* locus^19,20^, or were limited in terms of sample size or coverage^21–24^. The largest epigenome-wide association study (EWAS) to date included nearly 1,500 samples, but identified only a single significant differentially methylated position (DMP)^25^.

We present a blood-based DNA methylation study of ALS incorporating over 10,000 samples, representing the largest case-control study of DNA methylation for any disease to date. First, we implemented a stringent pipeline to meta-analyze EWAS results across multiple strata and identify DMPs associated with ALS. Second, we show how DNA methylation data can be leveraged to serve as a proxy for risk factors and exposures relevant to ALS. Third, we integrate our DNA methylation data with findings from a new genome-wide association study (GWAS) of ALS^26^, identifying potential causal roles for pathways highlighted by our DNA methylation study. Finally, we used overlapping whole-genome-sequencing (WGS) data to assess the influence of genetic variants and leveraged extensive clinical data for survival analyses.

## Results

### EWAS meta-analysis of ALS identifies 45 differentially methylated loci

We quantified genome-wide levels of DNA methylation in whole blood from 10,462 individuals across four strata using the Illumina 450k array (6,275 samples) and the Illumina EPIC array (4,187 samples). A total of 6,763 ALS patients and 2,943 control subjects passed our stringent quality control, which was followed by normalization of signal intensities in each stratum (**Table 1, supplementary file 1 and supplementary tables 1-5**). Samples excluded from our analyses did not show different demographic or clinical characteristics compared to the subset selected for analyses (**supplementary table 5).** We performed an EWAS in each of the four strata using two methods to adjust for known and unknown confounding. First we used a linear model adjusting for known confounders and a calibrated number of principal components (PCs) to adjust for unknown confounding factors (**Methods**; **supplementary figure 1**), followed by correction for residual bias and inflation in test-statistics using *bacon*^27^ (hereafter referred to as the LB model). Second, we used MOA (mixed linear model-based omic association) as implemented in the OSCA software, in which the random effect of total genome-wide DNA methylation captures the correlation structure between probes and directly controls for the genomic inflation^28^. Of note, the MOA algorithm did not converge for the AUS2 stratum, resulting in a total sample size of 9,459 for the MOA results. Test-statistics across strata were combined using an inverse variance-weighted fixed effects meta-analysis^29^. Inflation of the test-statistics was well controlled in both the LB (*λ* = 1.046; **Figure 1c**) and the MOA results respectively (*λ* = 0.984; **Figure 1d**), and we observed little heterogeneity between strata (**Methods**; **supplementary figure 2**). Various technical sensitivity analyses indicated that the results were robust to changes in analysis strategy, including using M-values instead of β-values, using functional normalization^30^ instead of *dasen*^31^, and excluding specific strata or experimental batches (**supplementary figure 3-4).** Finally, application of a method that we recently described^32^ led to the removal of likely cross-hybridizing probes, including four probes that showed high homology to the C9 repeat locus (**supplementary figure 5**). In total, 724,712 sites passed quality control, of which 332,066 were specific to the EPIC array.

**Figure 1.**
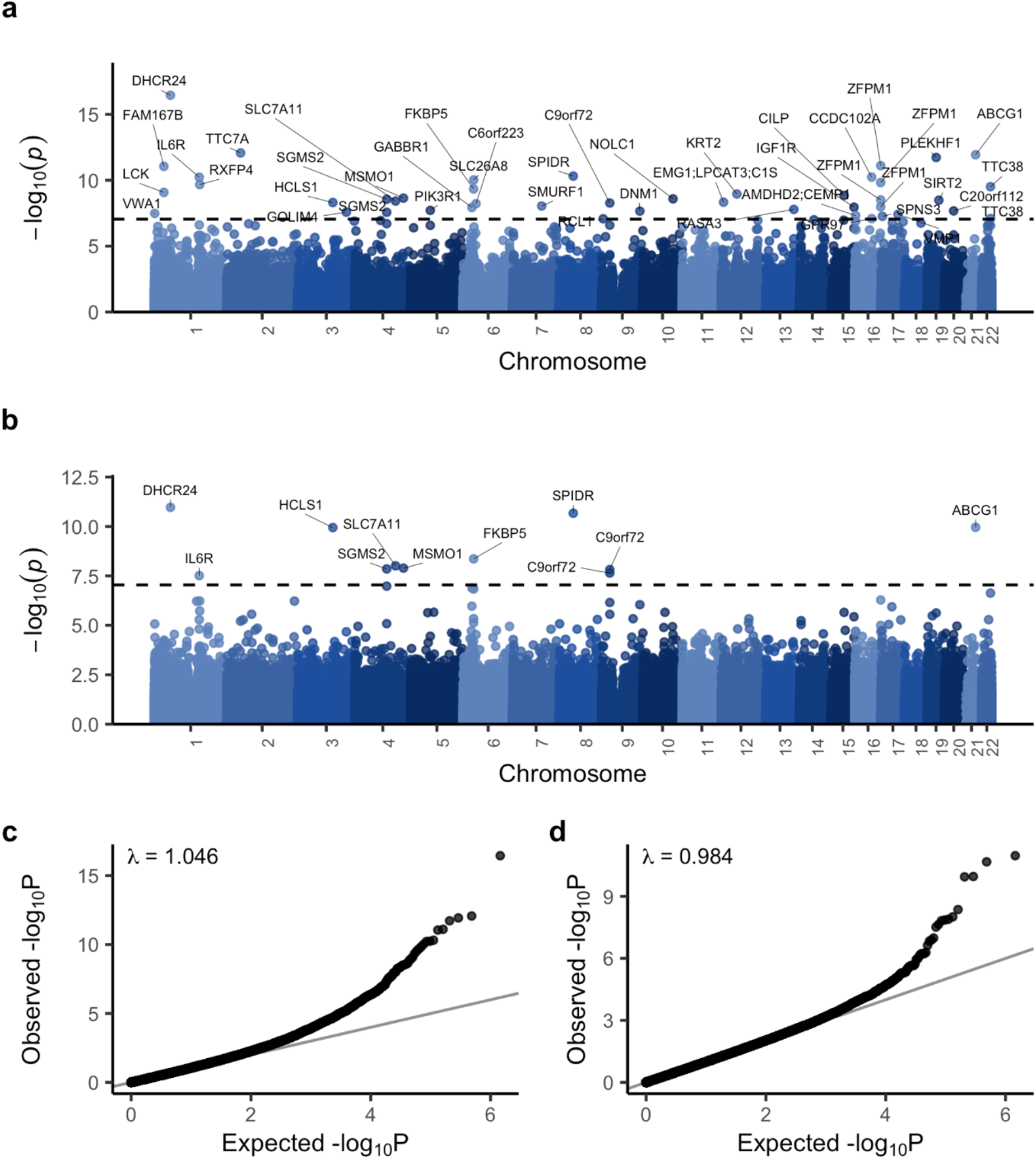
EWAS meta-analysis: Epigenome-wide association study on 6,763 patients and 2,943 controls. **(a,b)** Manhattan plot comparing **(a)** LB (linear model + *bacon*) and **(b)** OSCA MOA association *P*-values (−log 10(*P*), y-axis) and genomic location (x-axis). The dashed line indicates the genome-wide significance threshold (9 × 10^−8^). Sites were annotated with the nearest protein-coding gene in ensembl. **(c,d)** QQ-plot showing observed **(c)** LB and **(d)** OSCA MOA *P*-values (−log 10(*P*), y-axis) against the expected distribution under the null (x-axis).

**Table 1.**
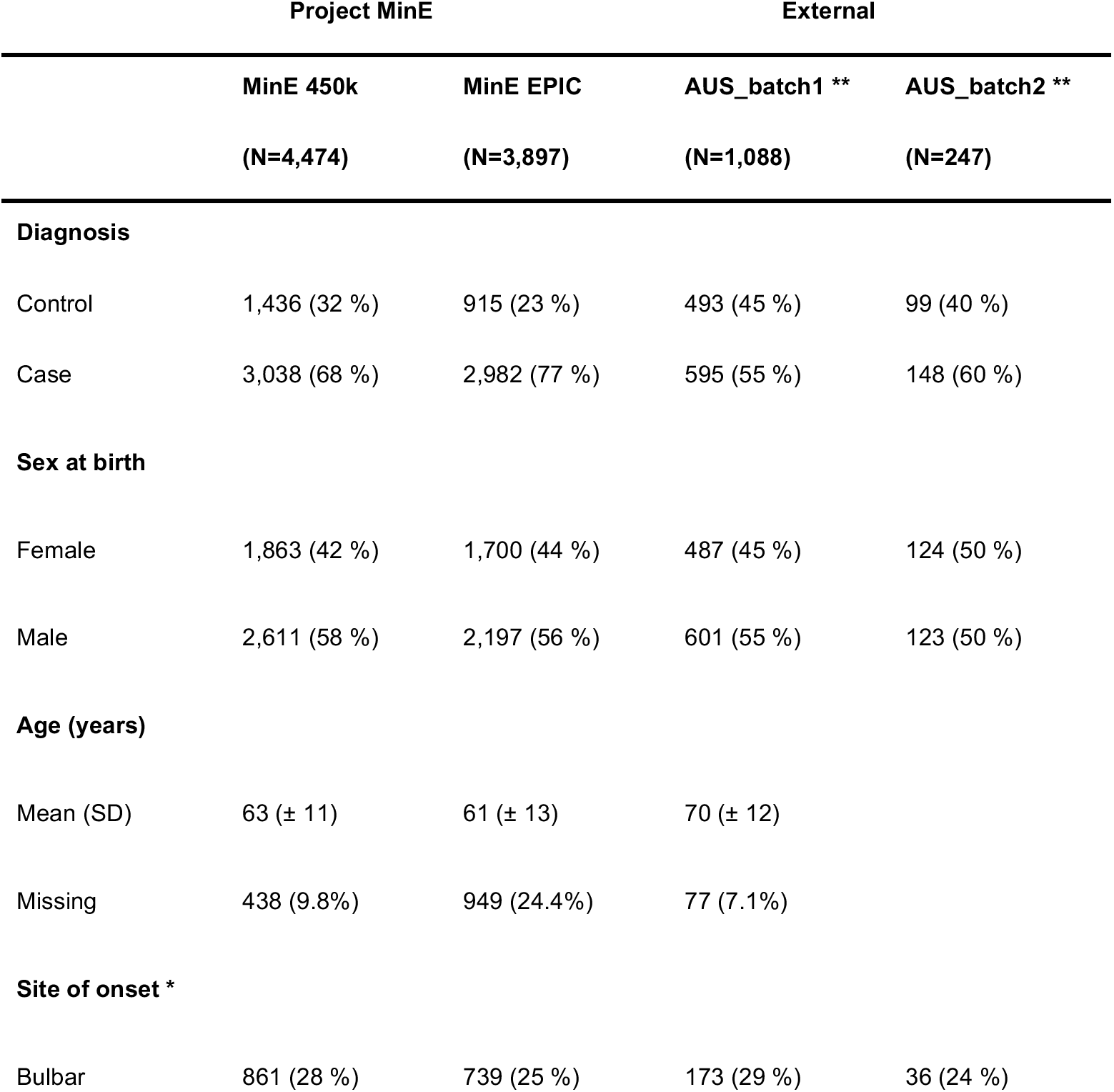
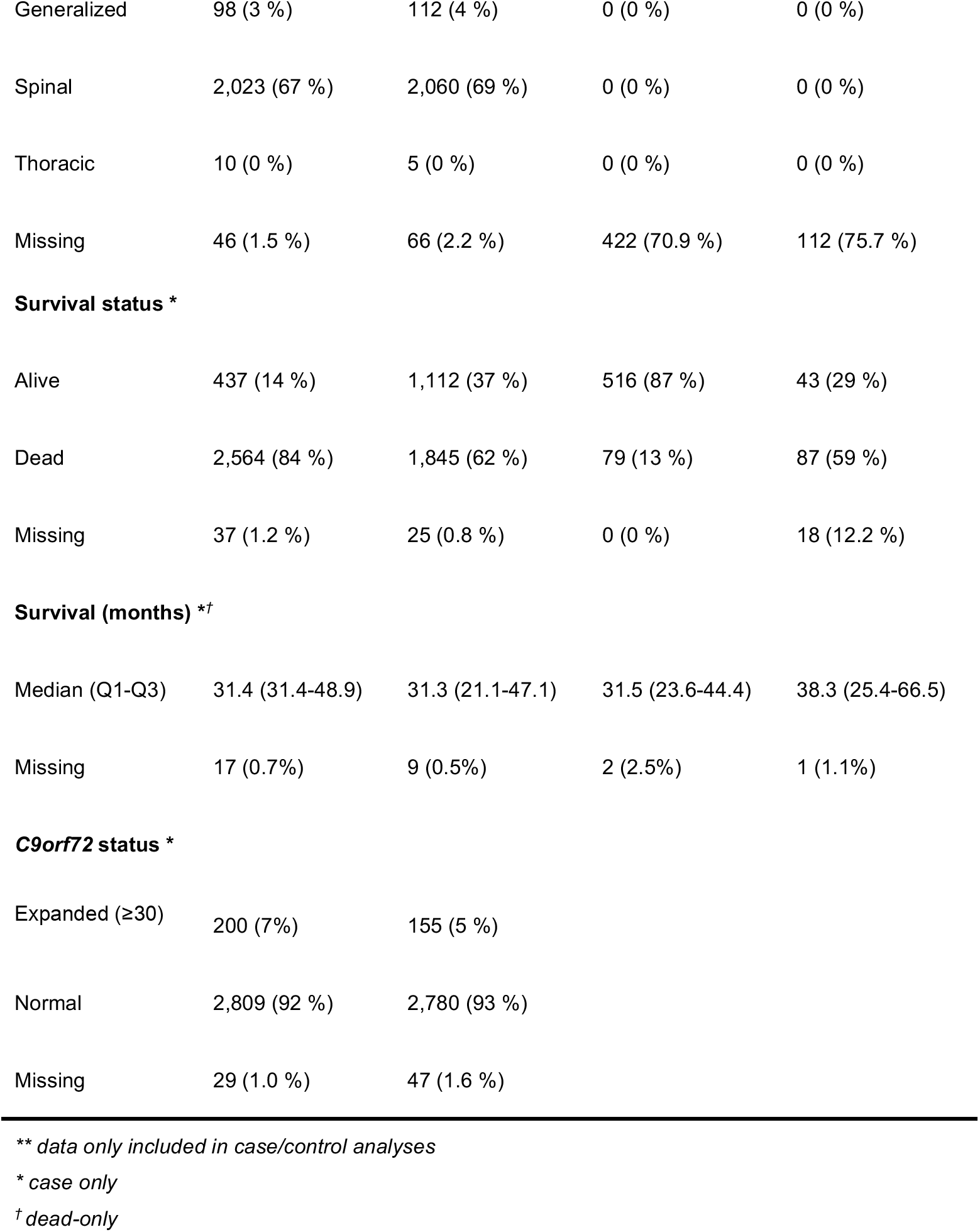
Demographic and clinical characteristics of study population: Shown are numbers (and percentages) of samples that passed qual ty control.

|  | Project MinE |  | External |  |
| --- | --- | --- | --- | --- |
|  | MinE 450k | MinE EPIC | AUS_batch1 ** | AUS_batch2 ** |
|  | (N=4,474) | (N=3,897) | (N=1,088) | (N=247) |
| **Diagnosis** |  |  |  |  |
| Control | 1,436 (32 %) | 915 (23 %) | 493 (45 %) | 99 (40 %) |
| Case | 3,038 (68 %) | 2,982 (77 %) | 595 (55 %) | 148 (60 %) |
| **Sex at birth** |  |  |  |  |
| Female | 1,863 (42 %) | 1,700 (44 %) | 487 (45 %) | 124 (50 %) |
| Male | 2,611 (58 %) | 2,197 (56 %) | 601 (55 %) | 123 (50 %) |
| **Age (years)** |  |  |  |  |
| Mean (SD) | 63 ($\pm 11$) | 61 ($\pm 13$) | 70 ($\pm 12$) | |
| Missing | 438 (9.8%) | 949 (24.4%) | 77 (7.1%) |  |
| **Site of onset \*** |  |  |  |  |
| Bulbar | 861 (28 %) | 739 (25 %) | 173 (29 %) | 36 (24 %) |
| Generalized | 98 (3 %) | 112 (4 %) | 0 (0 %) | 0 (0 %) |
| Spinal | 2,023 (67 %) | 2,060 (69 %) | 0 (0 %) | 0 (0 %) |
| Thoracic | 10 (0 %) | 5 (0 %) | 0 (0 %) | 0 (0 %) |
| Missing | 46 (1.5 %) | 66 (2.2 %) | 422 (70.9 %) | 112 (75.7 %) |
| **Survival status \*** |  |  |  |  |
| Alive | 437 (14 %) | 1,112 (37 %) | 516 (87 %) | 43 (29 %) |
| Dead | 2,564 (84 %) | 1,845 (62 %) | 79 (13 %) | 87 (59 %) |
| Missing | 37 (1.2 %) | 25 (0.8 %) | 0 (0 %) | 18 (12.2 %) |
| **Survival (months) \*†** |  |  |  |  |
| Median (Q1-Q3) | 31.4 (31.4-48.9) | 31.3 (21.1-47.1) | 31.5 (23.6-44.4) | 38.3 (25.4-66.5) |
| Missing | 17 (0.7%) | 9 (0.5%) | 2 (2.5%) | 1 (1.1%) |

**C9orf72 status \***
|  |  |  |
| --- | --- | --- |
| Expanded (≥30) | 200 (7%) | 155 (5 %) |
| Normal | 2,809 (92 %) | 2,780 (93 %) |
| Missing | 29 (1.0 %) | 47 (1.6 %) |

**\*\* data only included in case/control analyses**
**\* case only**
**† dead-only**

The LB meta-analysis resulted in 44 significant DMPs (*P <* 9 × 10^−8^; **Figure 1a, Table 2, supplementary table 6**) and the MOA meta-analysis resulted in 11 significant DMPs (*P <* 9 × 10^−8^; **Figure 1b, supplementary table 7**)^33^. The significant MOA sites comprised a subset of the significant LB sites, with the exception of cg01589155 which is annotated to the *C9orf72* locus; this site was significant in MOA (*P* = 1.51 × 10^−8^) and just below the significance threshold in the LB results (*P* = 2.59 × 10^−7^) (**supplementary figure 6**). Effect sizes were generally small and we observed both hypermethylated (51%) and hypomethylated (49%) DMPs associated with ALS (**supplementary figure 7**). Based on nearest gene mapping, these DMPs were annotated to 42 unique genes. Additionally, we annotated each site with *cis* expression associations in blood (eQTMs) calculated in an external dataset (BBMRI^34^). This revealed that DNA methylation at 18 sites was significantly associated with the expression levels of at least one nearby gene, which included the nearest gene in 14 out of 18 sites (**Table 2**, **supplemental table 8)**.

**Table 2.** Top ten significant sites: Details of the ten most significant sites identified with the LB algorithm. Position = Chromosome:bp (GRCh37), Nearest gene = nearest gene based on Ensembl GRCh37 (75), eQTM = the most significant eQTM for the respective probe, eQTM *FDR* = *p*-value corresponding to the most significant eQTM, FDR-corrected for the number of tests for the respective probe, PMS = Indicates that the probe is part of the respective PMS (poly-methylation score), Trait = Overlap with significantly enriched traits from the MRC-IEU and NGDC EWAS databases (showing a maximum of five traits). Abbreviations: HGF = Hepatocyte growth factor, N.CDase = Neutral ceramidase, FGF.21 = Fibroblast growth factor 21.

| Probe | Position | Nearest gene | eQTM (direction) | eQTM *FDR* | b | se | P-value | PMS | Traits |
| --- | --- | --- | --- | --- | --- | --- | --- | --- | --- |
| cg17901584 | 1: 55353706 | *DHCR24* | *DHCR24* (-) | $2.9 \times 10^{-62}$ | 0.0090 | 0.00110 | $3.6 \times 10^{-17}$ | BMI, HDLchol, HGF | Hepatic fat, BMI, Metabolic trait, (serum) Triglycerides |
| cg06528816 | 2: 47242277 | *TTC7A* | *TTC7A* (-) | 0.13 | 0.0035 | 0.00049 | $8.5 \times 10^{-13}$ | | Allergic sensitization, Total serum IgE |
| cg06500161 | 21: 43656587 | *ABCG1* | *ABCG1* (-) | $1.6 \times 10^{-25}$ | -0.0052 | 0.00073 | $1.2 \times 10^{-12}$ | BMI, HDLchol, N.CDase, FGF.21 | Hepatic fat, BMI, Metabolic trait, (serum) Triglycerides |
| cg14945937 | 19: 30162771 | *PLEKHF1* | *PLEKHF1* (-) | 0.02 | -0.0041 | 0.00058 | $1.9 \times 10^{-12}$ | | |
| cg08940169 | 16: 88540241 | *ZFPM1* | *PIEZO1*<sup>†</sup> (-) | 0.08 | 0.0037 | 0.00054 | $7.8 \times 10^{-12}$ | | Allergic sensitization, Total serum IgE, Childhood asthma, Schizophrenia |
| cg07571745 | 1: 32715428 | *FAM167B* | *CCDC28B*<sup>†</sup> (-) | 0.26 | -0.0033 | 0.00048 | $8.9 \times 10^{-12}$ | | |
| cg14195992 | 8: 48265917 | *SPIDR* | *SPIDR* (-) | 0.0059 | -0.0053 | 0.00080 | $4.8 \times 10^{-11}$ | | Birth weight |
| cg08851837 | 16: 57558820 | *CCDC102A* | *GPR56*<sup>†</sup> (+) | 0.84 | 0.0045 | 0.00069 | $5.8 \times 10^{-11}$ | | |
| cg09257526 | 1: 154379696 | *IL6R* | *ATP8B2*<sup>†</sup> (-) | 0.0031 | -0.0023 | 0.00035 | $5.9 \times 10^{-11}$ | | Alcohol consumption per day |
| cg15782984 | 6: 35993792 | *SLC26A8* | *SLC26A8* (-) | 0.007 | -0.0046 | 0.00070 | $9.5 \times 10^{-11}$ | | |

<sup>†</sup>The association between DNA methylation and the nearest gene was not significant (*FDR* > 0.05)

### Sensitivity analyses indicate that ALS-associated differential methylation is not driven by genetic variation in *cis* or *trans*, riluzole use or *C9orf72* status

We performed sensitivity analyses to evaluate whether our results were driven by known biological factors associated with ALS or by genetic variation. First, we examined the effects of the *C9orf72* (C9) repeat expansion by performing an EWAS meta-analysis excluding 371 carriers of this mutation. Overall the results were highly correlated (**supplementary figure 8)**, except for a subset of sites that were strongly driven by *C9orf72* carrier status. These included two genome-wide significant DMPs (cg01589155 and cg23074747) located within the C9 repeat and in a CpG island just upstream of the C9 repeat respectively. Second, to delineate whether DMPs were influenced by riluzole use, we performed an EWAS on riluzole use in ALS patients (N users = 1,803, N non-users = 451), finding no evidence of shared signals between the ALS EWAS and the riluzole EWAS (**supplementary figure 9**). Finally, we investigated whether results were driven by genetic variation. For each significant site we iteratively adjusted for all genetic variants in *cis* (*<*250 Kb) as detected in our overlapping whole-genome-sequencing data^26^ (WGS, N = 7,939) and blood *trans*-mQTLs as reported in the GoDMC consortium^35^. We found no evidence that the significant sites were driven by either genetic variants in *cis* or in *trans* (supplementary figure 10).

### Enrichment analyses of genes annotated to ALS-associated differential DNA methylation implicate metabolic, inflammatory and cholesterol pathways

#### Gene set analysis

To characterize the EWAS results we performed gene set enrichment analyses based on both nearest genes and eQTMs annotated to each site^36,37^. We considered both the default threshold used in the *methylGSA* package (*P* < 0.001) and the stringent genome-wide significance threshold (9 x 10^−8^) to select DMPs for enrichment analyses. We identified two main categories of enriched pathways.

First, in both the LB and MOA results we identified cholesterol/steroid biosynthesis-related pathways. These included the KEGG pathway steroid biosynthesis and the GO pathways cholesterol biosynthetic process, sterol biosynthetic process, organic hydroxy compound biosynthetic process and secondary alcohol biosynthetic process which were enriched among the MOA results (**Table 3**). In addition, we found that these and related pathways were enriched among annotated eQTMs in both the LB and MOA results (**supplementary table 9)**. The enrichments were mainly driven by four DMPs: cg17901584 (DHCR24), cg05119988 (MSMO1), cg06500161 (ABCG1) and cg06690548 (SLC7A11). Of these, cg17901584, cg05119988 and cg06500161 were strongly associated with the expression of the nearest gene in blood (DHCR24, MSMO1 and ABCG1 respectively; **Table 2**, **supplementary table 8**).

**Table 3.** Gene set enrichments: Details of the gene sets that were significantly enriched among the MOA and LB results based on nearest genes annotated to each site. Method = EWAS method and *p-*value cutoff applied to the respective EWAS test-statistics resulting in the input probes for the shown enrichment analyses, N overlap = Number of significant genes that overlap with genes in the respective pathway, N genes = Total number of genes in the pathway, *FDR* = FDR-controlled (False discovery rate) *P*-values.

| Method | Database | Pathway | N overlap | N genes | FDR |
| --- | --- | --- | --- | --- | --- |
| LB ($p < 0.001$) | KEGG | Cytokine-cytokine receptor interaction | 36 | 262 | 0.0012 |
|  | KEGG | Natural killer cell mediated cytotoxicity | 22 | 108 | 0.036 |
| MOA ($p < 0.001$) | - | - | - | - | - |
| LB ($p < 9 \times 10^{-8}$) | - | - | - | - | - |
| MOA ($p < 9 \times 10^{-8}$) | KEGG | Steroid biosynthesis | 2 | 18 | 0.015 |
|  | GO BP | cholesterol biosynthetic process | 3 | 71 | 0.021 |
|  | GO BP | sterol biosynthetic process | 3 | 77 | 0.021 |
|  | GO BP | organic hydroxy compound biosynthetic process | 4 | 251 | 0.021 |
|  | GO BP | secondary alcohol biosynthetic process | 3 | 71 | 0.021 |

Second, the immune-related KEGG pathways Cytokine-cytokine receptor interaction and natural killer cell mediated cytotoxicity were enriched in the LB results (at *P* < 0.001), but not in the MOA results (**Table 3**).

#### EWASdb enrichments

To further characterize the results we assessed whether the significant sites overlapped with trait-associated sites reported in publicly available EWAS databases^38,39^. For the LB results we found a significant overlap (*FDR* < 0.05) with 23 traits in the MRC-IEU database (**Figure 2a; Table 4**) and 20 traits in the NGDC database (**supplementary figure 11**, **supplementary table 10-11**), with a total of 23 out of 44 probes overlapping with one or more enriched traits. For the MOA results we found a significant overlap (*FDR* < 0.05) with 20 traits in the MRC-IEU database (**supplementary figure 12**, **supplementary table 10-11**) and 14 traits in the NGDC database (**supplementary figure 13**, **supplementary table 10-11**), with a total of 8 out of 11 probes overlapping with one or more of the enriched traits.

**Figure 2.**
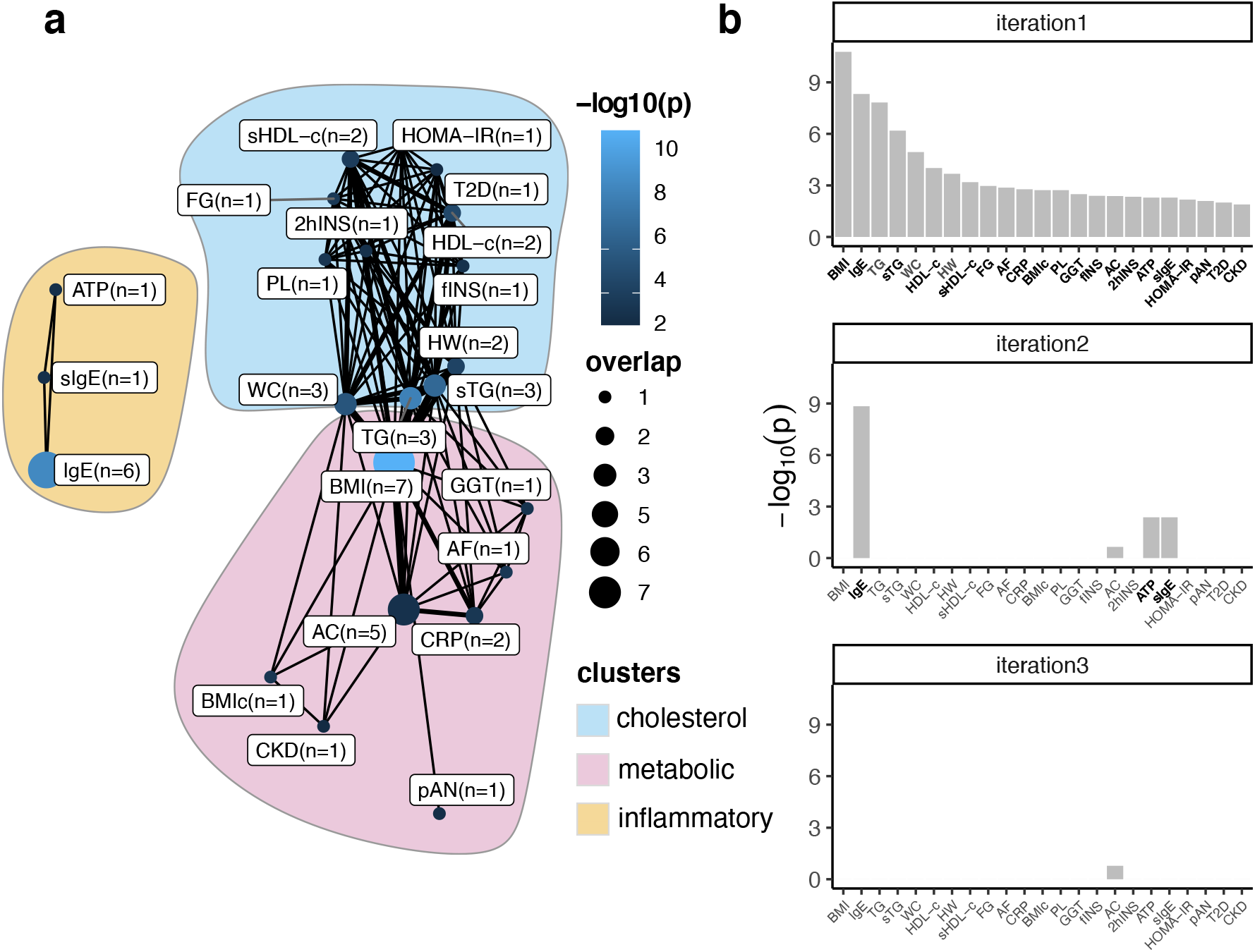
EWAS database enrichments: Significant overlap between traits included in the MRC-IEU EWAS database and ALS-associated sites identified using the LB model **(a)** Network showing the traits that significantly overlap with the ALS-associated sites. Nodes indicate the overlap between ALS-associated sites and sites associated with indicated traits, with larger nodes indicating more overlap, and lighter shades of blue indicating stronger associations. Edges indicate probe overlap between the traits, with thicker lines indicating more overlapping probes. Colored surfaces indicate the clusters (cholesterol, metabolic and inflammatory) identified using the Louvain clustering algorithm. **(b)** Identification of independent clusters of traits. The first iteration shows the traits that significantly overlap with the ALS-associated probes at *FDR <* 0.05. In subsequent iterations the probes belonging to the most significant trait were excluded and enrichments tests were performed using the remaining traits. This algorithm was repeated, retaining traits that were nominally significant (*P <* 0.05, indicated in bold), until at most one trait remained significant. At the third iteration no traits remained significant, showing that both BMI and related traits (including triglycerides and HDL-c) and IgE and related traits (Atopy) show independent overlap with the ALS-associated sites. Abbreviations: IgE = total serum IgE, TG = triglycerides, sTG= serum triglycerides, WC = waist circumference, sHDL-c = serum HDL-c, HW = Hypertriglyceridemic waist, FG = fasting glucose, AF = atrial fibrillation, BMIc = BMI change, PL = postprandial lipemia, GGT = Gamma-glutamyl transferase, fINS=fasting insulin, AC = alcohol consumption per day, 2hINS = 2-hour insulin, ATP = atopy, sIgE = High serum IgE, pAN = Plasma adiponectin, T2D = Type II diabetes, CKD = Chronic kidney disease, HOMA-IR = Homeostatic Model Assessment of Insulin Resistance.

**Table 4.** EWAS database enrichments: Ten most significant trait enrichments within the MRC-IEU EWAS database. *FDR* = FDR-corrected *P*-values (False discovery rate). Effect directions = indicate whether the ALS EWAS and trait EWAS effect sizes share the same direction of effect (e.g. an opposite direction of effect for Body mass index indicates that DNA methylation changes at overlapping sites associated with a *lower* BMI are also associated with a *higher* ALS risk); EWAS method = indicates whether significant sites identified with respective method were enriched for the given trait.

| Trait | *FDR* | Effect directions | EWAS method |
| --- | --- | --- | --- |
| Body mass index | $1.36 \times 10^{-9}$ | opposite | LB & MOA |
| Total serum IgE | $1.93 \times 10^{-7}$ | opposite | LB |
| Triglycerides | $4.02 \times 10^{-7}$ | opposite | LB & MOA |
| Serum triglycerides* | $1.32 \times 10^{-5}$ | opposite | LB & MOA |
| Waist circumference | $1.85 \times 10^{-4}$ | opposite | LB & MOA |
| High-density lipoprotein cholesterol | 0.0013 | equal | LB & MOA |
| Hypertriglyceridemic waist | 0.0024 | equal | LB & MOA |
| Serum high-density lipoprotein cholesterol* | 0.0066 | equal | LB & MOA |
| Fasting glucose | 0.0097 | opposite | LB & MOA |
| Atrial fibrillation | 0.011 | opposite | LB & MOA |

\*note that we adhered to the trait descriptions as provided in the database: serum, plasma and whole-blood measurements are included as distinct traits ("Triglycerides" and "High-density lipoprotein cholesterol" refer to whole-blood measurements).

Among the most significant enrichments in the MRC-IEU database (all results shown in **supplementary table 10-11**) were BMI, Total Serum IgE (only enriched among the LB results), (Serum) Triglycerides, Waist circumference and HDL-cholesterol, of which all showed effect directions opposite to those found for ALS, except for HDL (**Table 4**). Using the Louvain clustering algorithm^40^, we found that the overlapping traits clustered into two (MOA) to three (LB) clusters respectively. These included two connected cholesterol (including HDL-c and triglycerides) and metabolism-related (including BMI and alcohol consumption) clusters which were identified in the results from both EWAS methods. Additionally, in the LB results we identified an inflammation-related trait cluster that included traits such as total serum IgE and atopy. We found that this inflammation-related cluster was independent of the other clusters, as indicated by iterative analyses presented in **figure 2b**, showing that only the immune-related traits remained significant after excluding BMI-related probes (**supplementary figures 11-13**).

### Poly-methylation scores for BMI, HDL-c, alcohol intake and white blood cell proportions are associated with ALS

To gain further insight into potential intermediate phenotypes associated with ALS, we utilized published poly-methylation scores (PMS) as proxies for various traits and exposures, including BMI, HDL-c, LDL-c, total cholesterol, alcohol consumption, smoking, white blood cell proportions (CD4T, CD8T, monocytes, granulocytes, NK-cells), biological age and a collection of immunological and neurological proteins^11–14,16,41–43^.

First, we performed a validation analysis for each of the PMSs for which we had relevant clinical/exposure data available (see Methods, **supplementary table 4**). We selected PMSs with an explained variance of ≥ 5%, as indicated by an incremental R^2^ between the null model (including known covariates and control probe PCs) and the model including the respective PMS (figure 3a). Two out of the thirteen validated PMS did not meet the implemented threshold of ≥ 5% (LDL-c and total cholesterol).

**Figure 3.**
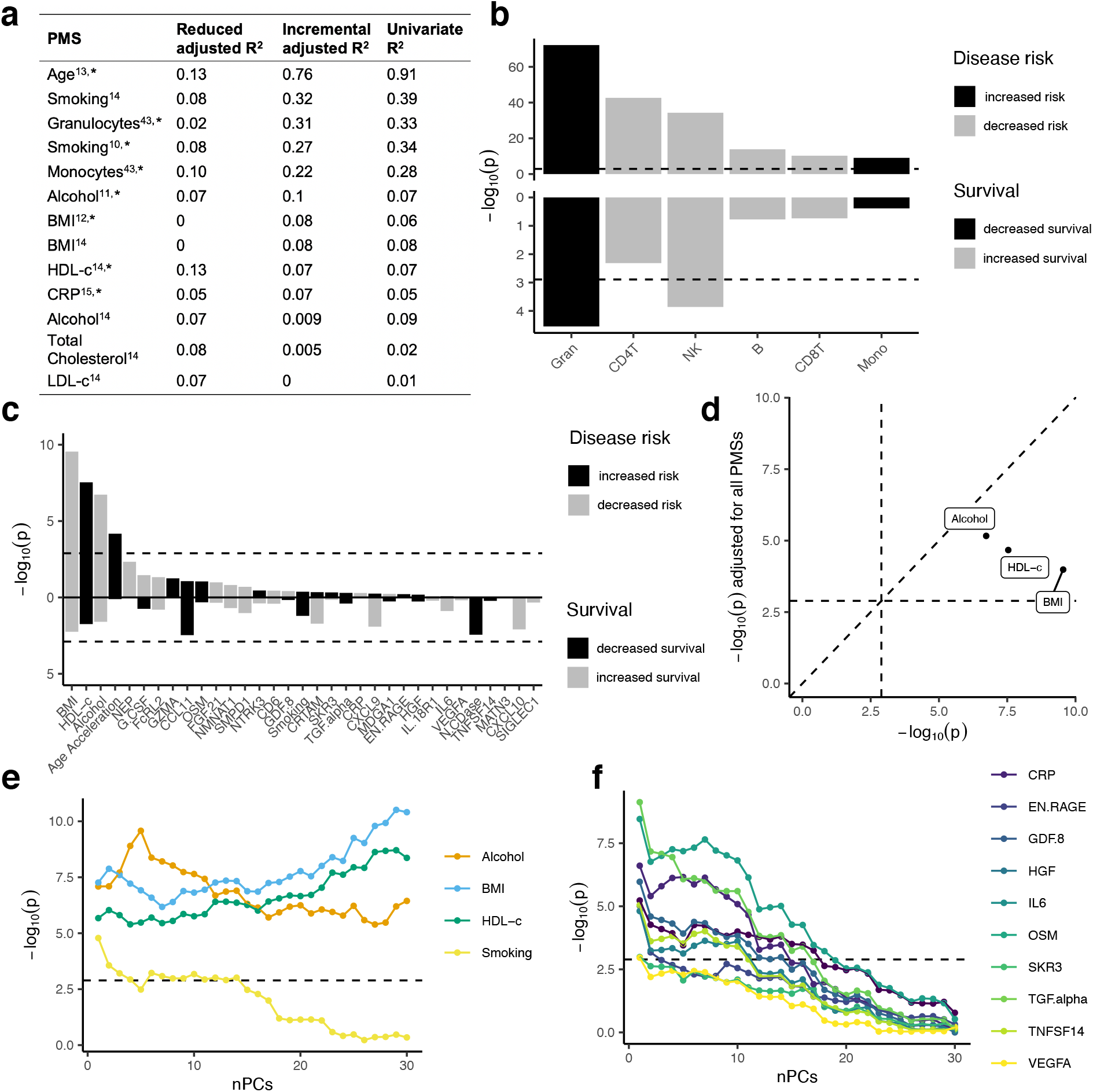
Poly-methylation score analyses on disease risk and patient survival: Poly-methylation scores (PMS) were determined as proxies for various traits, exposures, proteins and white blood cell proportions, calculated as weighted sums based on probes and weights derived from published papers respectively. **(a)** Explained variance of PMSs calculated in samples for which both DNA methylation data and biomarker/clinical data were available (N=800/2000). Reduced R^2^ represents the variance explained by the null model while the incremental R^2^ represents the additional variance explained by the PMS over the null model. Lastly, the explained variance of the univariate model of the respective PMS is displayed (see Methods). The asterisk indicates that the PMS was used in the association tests. **(b,c)** The upper panel shows association *P*-values (−log10(*P*), y-axis) for each PMS (x-axis), **(b)** white blood cell proportions and **(c)** various traits and exposures, colored by whether a higher score is associated with increased (black) or decreased (grey) disease risk. The lower panel shows the Cox proportional hazard *P*-values (−log10(*P*), y-axis) for each PMS (x-axis), colored by whether a higher score is associated with decreased (black) or increased (grey) survival respectively. The dashed line indicates the significance threshold (1.3 x 10^−3^). **(d)** Original *P*-values (−log10(*P*), x-axis) compared to *P*-values after including all PMSs in the logistic regression model (−log10(*P*), y-axis) for the significant traits/exposures. **(e, f)** Associations *P*-values (−log10(*P*), y-axis) upon incrementally adding principal components (PCs) to the logistic regression model.

We found that PMSs for HDL cholesterol, monocyte cell proportion and granulocyte cell proportion were positively associated with ALS (*P <* 1.3 × 10^−3^; **Figure 3 b-c, supplementary table 12**), and the PMSs for alcohol intake, BMI and the other white blood cell proportions (CD4T, CD8T, NK, B-cells) were negatively associated with ALS, a result that reflects the nature of proportion data given the positive associations of other cell types (*P <* 1.3 × 10^−3^; **Figure 3 b-c, supplementary table 12**). Although we did find a significant association for epigenetic age acceleration (Zhang *et al.* clock^13^, adjusted for chronological age), there was significant heterogeneity between strata (Cochran’s Q test *P* < 0.1/39), **supplementary table 12**), which led us to exclude age acceleration for further consideration. Additionally, we considered the multi-tissue Horvath clock^44^ and the Hannum *et al.* clock^45^, but found no significant associations for either (**supplementary figure 14**).

In addition to the PC-adjusted models, we also evaluated less stringent models, showing that various immunological and neurological proteins such as CRP, IL6, TGF-ɑ and CCL11 as well as smoking were significantly associated with ALS when PCs were excluded (**Figure 3e-f, supplementary figure 15**).

Conditional analyses showed that the significant PMSs were independently associated with ALS, although the HDL-c and BMI associations were attenuated after mutual adjustment (**Figure 3d**, **supplementary figure 16**). Adjustment for significant EWAS sites showed that signal is shared between several CpG sites and significant PMSs (**supplementary figure 17**), most notably, the alcohol intake association became insignificant upon adjustment for cg06690548 (*SLC7A11*) and cg18120259 (*C6orf223*), and the HDL-c association became insignificant upon adjustment for cg17901584 (*DHCR24*) and cg06500161 (*ABCG1*). We assessed whether the associations were primarily driven by carriers of the *C9orf72* repeat expansion, but found no evidence that this was the case (**supplementary figure 18**). Finally, we evaluated whether the PMS associations were primarily driven by specific strata or experimental batches by performing leave-one-out analyses. We found no evidence that one stratum or experimental batch mainly contributed to the observed signals, and as expected, the largest decrease in significance was seen when leaving out the larger experimental batches and strata (**supplementary figure 19-22**).

### Survival analyses indicate that white blood cell proportions and DNA methylation at five ALS-associated DMPs are associated with disease progression

We performed multivariate Cox PH analyses on the 45 ALS-associated DMPs identified using the MOA and LB models. Test-statistics were combined using inverse variance-weighted fixed effects meta-analysis and checked for heterogeneity (Cochran’s Q test *P* < 0.1/45). A total of five probes showed a significant association with survival after correcting for known confounders and PCs (0.05/45=*P <* 1.11 × 10^−3^) and cross-validation between three sensitivity analyses. Effect sizes were moderate and showed both shorter and longer survival time between DNA methylation and overall survival (**supplementary table 13**).

All reported sites were not significantly affected by the addition of time-varying effects in the Cox PH model or by applying a restricted cubic spline with varying complexity to model the baseline log cumulative hazard (**supplementary figure 23)**. Moreover, after adjusting for *C9orf72* carrier status in the multivariate Cox PH model, the significant sites (besides the *C9orf72* mapped probe) remained significantly associated with survival (**supplementary figure 23)**. Four sites showed a significant eQTM effect with *FKBP5*, *ATP8B2*, *SPIDR*, and *DHCR24* (**Table 5**).

**Table 5.**
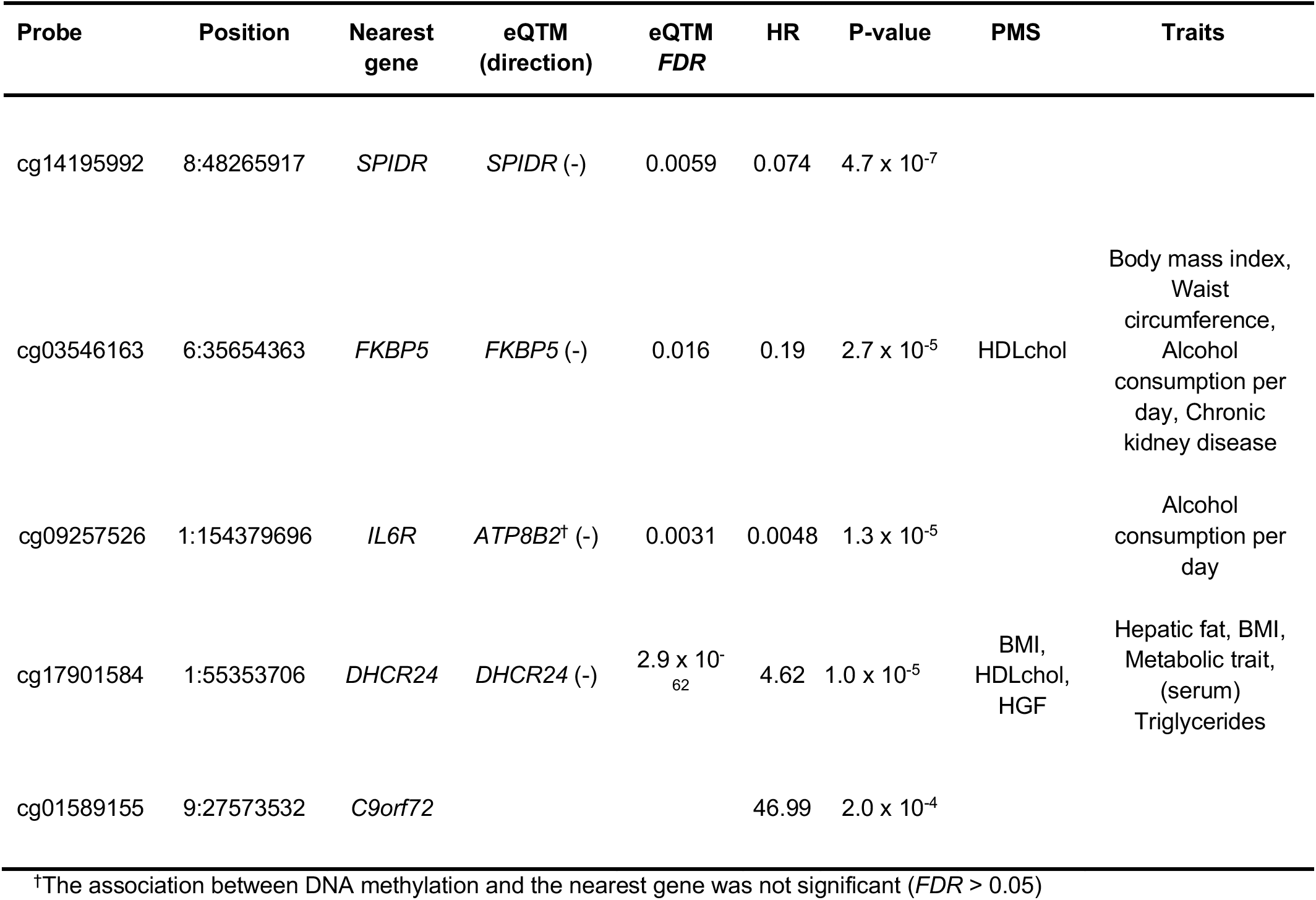
Significant sites associated with survival: Details of the sites significantly associated with survival. Position = Chromosome:bp (GRCh37), Nearest gene = nearest gene based on Ensembl GRCh37 (75), eQTM = the most significant eQTM for the respective probe, eQTM *FDR* = *p*-value corresponding to the most significant eQTM, FDR-corrected for the number of tests for the respective probe, PMS = Probe is part of the respective PMS (poly-methylation score), HR = Hazard Ratio, Trait = Overlap with significantly enriched traits from the MRC-IEU and NGDC EWAS databases (showing a maximum of five traits). Abbreviations: HGF = Hepatocyte growth factor.

| Probe | Position | Nearest gene | eQTM (direction) | eQTM *FDR* | HR | P-value | PMS | Traits |
| --- | --- | --- | --- | --- | --- | --- | --- | --- |
| cg14195992 | 8:48265917 | *SPIDR* | *SPIDR* (-) | 0.0059 | 0.074 | $4.7 \times 10^{-7}$ | | |
| cg03546163 | 6:35654363 | *FKBP5* | *FKBP5* (-) | 0.016 | 0.19 | $2.7 \times 10^{-5}$ | HDLchol | Body mass index, Waist circumference, Alcohol consumption per day, Chronic kidney disease |
| cg09257526 | 1:154379696 | *IL6R* | *ATP8B2*<sup>†</sup> (-) | 0.0031 | 0.0048 | $1.3 \times 10^{-5}$ | | Alcohol consumption per day |
| cg17901584 | 1:55353706 | *DHCR24* | *DHCR24* (-) | $2.9 \times 10^{-62}$ | 4.62 | $1.0 \times 10^{-5}$ | BMI, HDLchol, HGF | Hepatic fat, BMI, Metabolic trait, (serum) Triglycerides |
| cg01589155 | 9:27573532 | *C9orf72* | | | 46.99 | $2.0 \times 10^{-4}$ | | |

<sup>†</sup>The association between DNA methylation and the nearest gene was not significant (*FDR* > 0.05)

We also assessed whether the PMSs were associated with survival, finding that a higher proportion of granulocytes was significantly associated with decreased survival and a higher proportion of natural-killer cells was associated with increased survival (*P* < 1.3 × 10^−3^; lower panels **Figure 3b-c, supplementary table 14**). These results were robust in different models: a Cox PH model stratified on experimental batch and with time-varying effects and a restricted cubic spline to model the baseline log cumulative hazard (**supplementary figure 24-25**). Finally, the associations were not affected upon adjustment for C9orf72 status (**supplementary figure 26)**.

## Discussion

Our study represents the largest case-control EWAS of any disease performed to date. Our unique dataset includes genome-wide DNA methylation data on over 10,000 individuals, with extensive clinical data and whole genome sequencing data available for the majority of samples. Following thorough quality control and extensive sensitivity analyses, we identified a total of 45 DMPs at which DNA methylation was significantly and robustly associated with ALS, with the majority representing novel findings not previously reported. Both gene set and trait enrichment analyses revealed enrichments for metabolic, cholesterol and inflammatory pathways. These findings were corroborated using a hypothesis-driven approach that leveraged published DNA methylation-based proxies for risk factors and exposures relevant to ALS. These analyses found that PMSs for BMI, HDL-cholesterol were independently associated with ALS. Moreover, we found a strong association between white blood cell PMSs and ALS, and show that an increased proportion of granulocytes is associated with worse patient survival. Finally, we show that several of the DMPs associated with ALS were also associated with survival, and could therefore be of interest as and could represent indicators of underlying disease processes.

By utilizing enrichment analyses, poly-methylation scores and survival analyses we highlight various pathways and potential disease-modifiers in ALS.

First, genes annotated to DMPs were enriched for pathways related to cholesterol biosynthesis. The main drivers of these enrichments include cg17901584 (*DHCR24*), cg06500161 *(ABCG1*), cg05119988 (*MSMO1*), and cg06690548 (*SLC7A11*). Of these, we found that DNA methylation levels at the first three sites were significantly associated with gene expression in blood. These genes are all involved in cholesterol biosynthesis and lipid transport, and DNA methylation at these sites has been robustly linked to HDL- and total cholesterol, triglyceride levels and BMI-related traits such as diabetes and hepatic fat levels^46–52^. Moreover, both cg17901584 (*DHCR24*) and cg06500161 *(ABCG1*) are part of the HDL-c PMS, and explain a considerable part of the significant association we found between elevated HDL-cholesterol levels and ALS.

Interestingly, cg06690548 (annotated to *SLC7A11*) has also been previously associated with alcohol intake and related factors such as GGT and phosphatidylethanol levels^11,53–55^, and the association between the alcohol PMS and ALS was primarily driven by this site. Although we did not find a significant DNA methylation-gene expression relationship (eQTM) for this site, previous work suggests that increased DNA methylation at cg06690548 is associated with downregulation of *SLC7A11* in the brain. Interestingly, *SLC7A11* encodes xCT, a cystine-glutamate antiporter that imports cystine while exporting glutamate^56^, the former being an essential precursor of glutathione, the major antioxidant in the brain. It is possible therefore, that the association found in *SLC7A11 --* and by extension the alcohol PMS -- is related to glutamate excitotoxicity and/or oxidative stress, rather than reflecting alcohol use.

Second, both the EWAS trait enrichments and PMS analyses indicate that lower BMI levels are associated with ALS. Importantly, the BMI association remained significant after adjustment for other PMSs, including those of HDL-c and alcohol intake, although these PMSs are not perfect proxies of the respective covariates. Lower BMI has been consistently reported in ALS, and may be explained by several factors including hypermetabolism^57^, increased levels of physical activity prior to disease onset^58^ or muscle loss and eating difficulties after onset of disease. It is also interesting to note that our findings support the hypothesis that ALS is associated with a favorable cardiovascular profile^59,60^. Supporting findings include lowered BMI and higher HDL-c levels in ALS patients as indicated by the PMS analyses, and opposite effects at overlapping sites between ALS and traits such as BMI, triglycerides, waist circumference, fasting glucose, and type II diabetes as indicated by the trait enrichment analyses. However, since our study cannot reliably distinguish whether phenomena occur prior to or after onset of disease, we need to be cautious to conclude that a favourable cardiovascular profile is a risk factor for developing ALS. The Mendelian randomization (MR) analyses in our accompanying GWAS study^26^ do not support a causal role for cardiovascular traits including BMI, triglycerides, blood pressure, smoking, physical activity, BMI, coronary artery disease, or stroke, but do support a causal role for cholesterol. This shows that the causal role for cholesterol in ALS might be independent from a potential cardiovascular mechanism, for example through the interplay between lipid levels and autophagy^61^ as illustrated by a recent study showing that high cholesterol levels lead to increased protein aggregation through autophagy impairment in mouse models of Alzheimer’s disease^62^. Whether the other independently related traits (BMI and alcohol intake) are causally associated with ALS, or are indicators for ALS pathophysiology, needs further study. These relations can be complex, for example we recently showed that BMI, smoking, alcohol intake and physical activity can have a genotype and time dependent relationship with ALS^63^.

Third, our results point towards a role of the immune system in ALS. The EWAS results were enriched for immune-related traits including IgE and allergic sensitization, and importantly, these results were independent of predicted white blood cell proportions. Sites driving these enrichments included, among others, cg06528816 (*TTC7A*) and a cluster of three probes in the *ZFPM1* gene, both implicated in immune-related traits such as IgE^64^, asthma^65,66^ and allergic sensitization^67^. Our PMS analyses corroborate the role of immunity in ALS as we found that white blood cell proportions were altered in ALS, with a higher ratio of granulocytes and a lower ratio of lymphocytes in ALS patients (CD4T-cells, CD8T-cells, and NK-cells). Interestingly, a recent study showed that white blood cell alterations are shared between neurodegenerative diseases (Alzheimer’s, Parkinson’s disease and ALS)^68^. These findings indicate that WBC alterations are not specific to ALS and are therefore more likely to be a consequence of disease, and thus their value is more likely related to disease activity rather than etiological. Indeed, in our study we found that increased granulocyte proportions are associated with worse prognosis, whereas NK-cell proportions are associated with better prognosis, indicating that WBC proportions have prognostic value. The role of immunity is further supported by our observation that various inflammatory protein PMSs such as CRP, IL6, TGF-ɑ and CCL11 were elevated in ALS patients, although these differences became insignificant upon adjustment for principal components. Our findings are in line with previous studies that identified higher ratios of neutrophils and/or granulocytes to lymphocytes in ALS patients^69,70^, elevated levels of inflammatory proteins^71^, and an association between higher neutrophil proportions and worse prognosis^72^. Although immune alterations could be part of a systemic aspect of ALS, there is evidence that suggests that the peripheral immune system contributes to neuroinflammation, the latter being an established phenomenon in ALS as well as other neurodegenerative diseases^73,74^. Especially interesting in this regard is that recent evidence shows that mast cells infiltrate skeletal muscles at the neuromuscular junction and degranulate to help recruit neutrophils^75,76^, which prevent reinnervation capacity and may thus be a potential mechanism causing worse prognosis. In line with this, we identified an enrichment for IgE (and related traits such as allergy and atopy), which activate mast cells, and found that increased proportions of granulocytes were associated with ALS and patient survival. Thus, these findings could be of interest for new treatments, especially given that mast cell activity can be influenced^77^.

Finally, 8 out of the identified 45 sites were reported in a recent study on shared DNA methylation alterations across Parkinson’s disease, Alzheimer’s disease and ALS^68^. Of these ALS-associated DMPs we identified associations between DNA methylation and survival within 5 of the 45 DMPs annotated to FKBP5, ATP8B2, SPIDR, and DHCR24. Interestingly, the majority (4 out of 5) of sites that we found to be associated with disease progression were among the overlapping sites. It could therefore be speculated that these sites represent shared pathways involved in neurodegeneration and could therefore have clinical utility.

Several recent findings are relevant to our current study.

First, the largest ALS EWAS up to this date^25^ has reported that white blood cell proportions were associated with ALS. We replicate these findings, and additionally show that WBC proportions are associated with prognosis.

Second, the DNA methylation changes that we identified in the CpG island just upstream of the *C9orf72* G_4_C_2_ repeat and within the repeat itself have been previously reported in carriers of the *C9orf72* repeat expansion^78–80^. In line with this, we found that these associations were driven by the *C9orf72* carriers in our data.

Third, we do not replicate the recently reported association between epigenetic age acceleration and survival^81^. Importantly, in our analyses we adjusted for sampling age, as it has been shown to be crucial when studying epigenetic age acceleration^82^, especially given that age of onset affects disease progression in ALS^83^. As we show in **supplementary figure 27** both survival and age of onset were significantly associated with age acceleration when sampling age was not accounted for, but became insignificant upon adjustment. Additionally, in our case/control analysis we observed substantial heterogeneity among strata (Cochran’s Q test *P* = 5.8 x 10^−4^), hence our results do not support a unambiguous role for age acceleration.

One limitation of this study is that the cross-sectional nature of our study hinders inferences about causality. Mendelian randomization analyses, presented in our accompanying GWAS, can aid in identifying causal relations in observational data, as evidenced by the finding that cholesterol levels are causally related to ALS. The GWAS MR analyses (SMR^84^) did not find significant evidence for a causal role of the sites identified in this study. Although this may reflect a lack of power, it could indicate that the results reflect the consequences of disease processes rather than causal mechanisms. In that case, the value of the identified DNA methylation changes would lie primarily in revealing underlying disease processes in ALS. Furthermore, our survival analyses show that the results could be of interest as potential starting points for new disease-modifying treatments. Future studies could further investigate causality in longitudinal samples, which although scarce and often small, are expected to become more readily available with the increasing number of larger-sized prospective cohorts.

We further note that we collected DNA from whole blood rather than from brain tissue, given that our aim was to identify peripheral biomarkers reflecting underlying disease processes and traits/exposures related to disease. In contrast to brain tissue, blood DNA methylation is easily accessible and allows for sampling close to disease onset. Combined with the ever growing body of literature on, and biomarkers of, various traits and exposures, it makes blood DNA methylation ideal for biomarker purposes^85^. However, given the tissue-specificity of DNA methylation, further studies are needed to assess ALS-associated DNA methylation changes in the brain.

Finally, we note that although the stringent adjustment for confounding we applied by using PCs and random effects models (i.e. OSCA^28^) is key in combating test statistic inflation in EWAS, it may have obscured biological signals of interest. For example, we show that the recent OSCA MOA algorithm results in a substantially lower (but overlapping) number of identified DMPs compared to the often used method of including principal components followed by adjustment for inflation/bias in test-statistics (termed the LB algorithm in this study). Interestingly, our results indicate that the additional DMPs identified using the LB algorithm are enriched for inflammatory pathways and traits, which corroborates previous findings that suggest that uncaptured variation can be explained by cell type heterogeneity and related immune processes^86,87^. Similarly, we show that various immunological proteins such as CRP, IL6, TGF-ɑ and CCL11 became insignificant upon PC adjustment. More generally, this relates to the discussion on whether to treat variables such as cell type proportions as nuisance variables in an EWAS, or view them as variables that provide valuable information in themselves^88,89^. In this study we therefore struck a balance by opting for a two-way approach, combining a stringently corrected EWAS with a more targeted approach where we studied “confounders’’ such as WBC proportions, smoking and BMI as outcomes of interest, assessing them with both stringent (i.e. including PCs) as well as more lenient models.

To conclude, we present the largest case-control DNA methylation study to date, employing a comprehensive approach integrating methylomic, genomic and clinical data. We identify 45 differentially methylated sites, and by utilizing enrichment analyses, poly-methylation scores and survival analyses we highlight a potential disease-modifying role for caloric intake, cholesterol metabolism and inflammation. In our accompanying GWAS paper, we show that cholesterol levels are causally related to ALS. Regardless of the exact etiologic role of the other identified pathways, these constitute potential targets for intervention in ALS patients.

## Methods

### Cohorts

We obtained DNA methylation and phenotypic/lifestyle data from 10,462 individuals (cases N = 7,344, controls N = 3,118) in fourteen different cohorts. Patients were diagnosed with definite, probable, and probable lab-supported ALS according to the revised El Escorial Criteria^90^. Population-based controls were matched for age, sex, and geographical region in a 1:2 ratio and not screened for (subclinical) signs of ALS. Detailed descriptions of each cohort are provided in the **Supplementary Note**. Experimental batches were processed in the same lab and sequenced in the same series, resulting in 44 independent batches after quality control. Strata for analyses were defined as samples within the Project MinE sequencing consortium separated by array technology (MinE 450k and MinE epic) and Australian data separated by signal intensities (AUS 1 and AUS 2).

### DNA methylation profiling

For the Project Mine samples, venous blood was drawn from patients and controls from which genomic DNA was isolated using standard methods. We set the DNA concentrations at 100ng/l as measured by a fluorometer with the PicoGreen© dsDNA quantitation assay. DNA integrity was assessed using gel electrophoresis. Genomic DNA (∼1 µg) was bisulfite-treated at a central site using Zymo Bisulfite Conversion Kits (Zymo Research, Orange, CA). DNA methylation was analyzed using the Infinium Methylation450k array (N=4,903) or Infinium EPIC array (N=4,300), according to the standard Infinium HD array methylation protocol (Illumina, San Diego, CA, USA).

The AUS ALS datasets were profiled as described in ref^25^.

### QC & Normalization

#### Sample QC

Quality control and normalization was performed separately for the four strata (MinE 450k, MinE EPIC, AUS1 and AUS2). The following metrics were used to exclude samples, the exact thresholds used for each stratum are listed in **supplementary table 1,** and the number of samples failed on each metric are listed in **supplementary table 2.** QC figures for each stratum are provided in **supplementary file 1.**

1. Median methylated and unmethylated intensities.
2. Median red/green intensity ratios calculated in type I probes.
3. Discordance between reported sex and predicted sex based on the *getSex* function in the *minfi* R package^91^.
4. The OP (non-polymorphic controls) and Hyb (hybridization controls) metrics as implemented in the *MethylAid* R package^92^.
5. Bisulfite conversion rate based on the *bscon* metric as implemented in the *wateRmelon* R package^31^.
6. Fraction of probes with high detection *P*-values and/or measured by a low number of beads.
7. Samples that failed on the inbreeding metric in the corresponding whole-genome-sequencing (WGS) data. Quality control of the Project MinE WGS data was performed as described earlier^93^.
8. Genotype concordance. Briefly, we used the *omicsPrint* R package to select 413 probes that reliably measured underlying SNPs and were present in the WGS-derived SNP data^94^. We performed identity-by-state (IBS) between the DNAm-inferred SNPs and the WGS-derived SNP data using the *allelesharing* function.
9. After removing samples that failed on any of the steps listed above, we performed PCA on the control probes present on the array. Samples that had values larger than 3.5 standard deviations of the mean on the first two principal components (PCs) were excluded.

#### Normalization

After quality control, signal intensities for all strata except AUS1 were normalized using the *dasen* function as implemented in the *wateRmelon* package^31^. Since the signal intensities in the AUS1 stratum showed dye bias (**supplementary file 1: QC figure 50**), this stratum was normalized using the *nanes* function, which is similar to the *dasen* function, except that it corrects for dye bias prior to normalizing the signal intensities.

#### Probe Filtering

After normalization, we set all the measurements with detection *P*-value *>* 1 × 10^−16^ or measured by *<*3 beads to missing^95^. Within each stratum we then removed probes with *>*5% missing data.

#### Post-normalization filtering

After sample QC, normalization and probe filtering, we further excluded samples based on the following criteria (**supplementary table 3)**:

1. Discordance between chronological (reported) age and predicted age^13^. In each stratum we regressed DNA methylation age on chronological age, and excluded samples whose residuals from this regression were more than 4.5 standard deviations from the mean.
2. We performed principal component analyses on the normalized *β*-values. Samples that had values larger than 4 standard deviations from the mean on the first two PCs were excluded.
3. Samples with non-MND diagnosis or missing phenotype info were excluded.
4. For each related pair of individuals (identical or first-degree) we excluded one individual to obtain a set of unrelated individuals.

### EWAS

We used two approaches to test for an association between disease status and variable DNA methylation while controlling for known and unknown confounding factors.

#### Principal components + bacon (LB model)

We performed linear regression at each site, adjusting for sex, experimental batch, predicted age, predicted white blood cell (WBC) fractions, 30 control probe PCs and a variable number of PCs derived from all measured sites. PCs were calculated by regressing the *β*-values on the covariates listed above, followed by performing principal component analysis on the residuals from this regression. We optimized the number of PCs included in each stratum by evaluating the sample-size normalized inflation factors (*λ*_1000_)^96^:

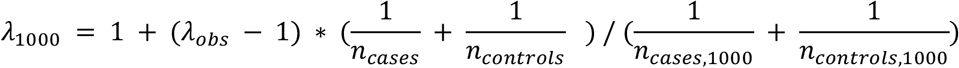

after employing the following linear regression at each site:

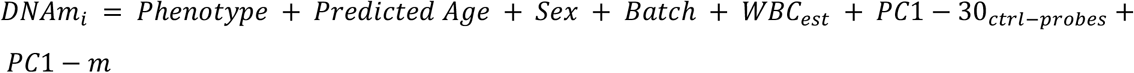

where *m* is in {0,5,10,15,20,25,30}.

The number of PCs were chosen so that for each stratum *λ*_1000_ ≤ 1.15. We then corrected for remaining inflation and/or bias in test-statistics of each stratum using *bacon* (implemented in the *bacon* R package)^27^. Hereafter we refer to this model (linear model followed by *bacon*) as the LB model.

#### Mixed linear model

We used a mixed-linear model approach as implemented in the MOA algorithm included in the OSCA software (v0.45)^28^. Briefly, this algorithm tests for an association between the methylation status of a CpG-site (*β*-value) and a trait (case/control status) while fitting all the other probes as random effects. This method is based on the assumption that including distal probes as random effects accounts for correlations induced by (unobserved) confounding factors. In addition to the random effects, we included sex, predicted age and experimental batch as fixed effects.

#### Meta-analysis

Test-statistics across strata were combined using an inverse variance-weighted fixed-effects meta-analysis as implemented in the *metagen* function in the *meta* package^29^. We assessed heterogeneity using Cochran’s Q test, and considered sites significantly heterogeneous if Cochran’s Q test *P*-value (corrected for the number of significant sites) was < 0.1. Sites where *P <* 9 × 10^−8^ were considered genome-wide significant, as recommended for the EPIC array based on an empirical estimate of the independent number of tests^33^.

### EWAS sensitivity analyses

We performed various sensitivity analyses to assess whether the EWAS results were robust to varying analysis strategies or driven by measured biological factors.

#### Technical Sensitivity Analyses

The following technical sensitivity analyses were performed:

1. We performed an EWAS on *M*-values (defined as 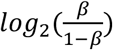) instead of *β*-values.
2. We performed an EWAS including only the Project MinE samples (N=8371; excluding the external Australian samples).
3. We performed an EWAS where we normalized the signal intensities using functional normalization^30^ instead of *dasen* and *nanes*^31^.
4. We performed an EWAS where we left out experimental batches that included only ALS patients and no controls.

#### Biological Sensitivity Analyses

The following biological sensitivity analyses were performed:

1. To assess whether results were influenced by riluzole use, we performed an EWAS on riluzole usage (yes/no) in 2,254 ALS patients for which data was available. These included 1,803 patients that used riluzole and 451 patients that did not use riluzole at time of blood draw respectively.
2. To assess whether results were driven by patients carrying the *C9orf72* (C9) repeat expansion (the most common genetic cause of ALS), we performed an EWAS where we excluded individuals carrying the C9 repeat expansion. We restricted the analysis to individuals for which C9 status was available, and who did not carry the C9 repeat expansion (N = 7,839, removed 371 C9 carriers; based on a ≥30 repeat cutoff as determined by ExpansionHunter^97^). We compared this C9-negative EWAS to an EWAS including C9-carriers where we randomly downsampled the number of samples to match the sample size of the C9-negative EWAS.
3. To assess whether the results were driven by genetic variation, we employed the available whole genome sequencing (WGS) data (available for 7,939 samples). We considered two sets of genetic variants: (1) all variants (SNVs) in *cis* of the CpG-site (<250Kb) and (2) variants reported as *trans*-mQTLs in blood for the respective CpG in the *GoDMC* consortium atlas of genetic effects (for 11 DMPs at least one *trans*-mQTL was reported)^35^. For each SNP (*j*) annotated to CpG-site (*i*), we ran the following regression:

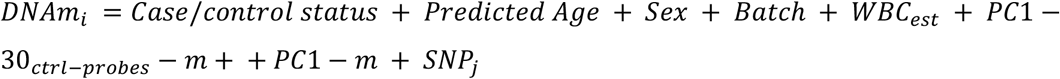

Where *m* = 30 for the MinE 450k stratum and *m* = 15 for the MinE EPIC stratum.

### Probe filtering

Probes were filtered based on the following criteria:

1. A ≥14bp 3’-subsequence inexact match to the C9 repeat expansion^32^
2. A ≥30bp 3’-subsequence inexact off-target match to the reference genome.
3. Low mapping quality based as determined by Zhou *et al.*^98^.
4. Given that we have genetic data available for most samples, we tested whether significant probes were driven by nearby (*<*250Kb) genetic variants, instead of removing probes containing SNPs *a priori*. Non-significant probes (used as background for enrichment analyses) were filtered based on containing a SNP with MAF *>* 0.01 (European population) within 5bp of the 3’-end of the probe^98^.

### Gene mapping & eQTMs

We mapped CpG-sites to the nearest protein-coding gene within 250kb based on Ensembl (GRCh37).

We tested whether the DNA methylation levels of the significant CpG-sites were associated with the expression of nearby genes in blood using an external dataset (BBMRI; https://www.ebi.ac.uk/ega/studies/EGAS00001001077) for which gene expression (RNAseq) and DNA methylation data (450k array) are available for 3,251 samples^34^. For each CpG-site, we tested for an association between DNA methylation and gene expression levels of nearby genes (*<*250kb), correcting for age, sex, strata, white blood cell composition and 20 PCs (10 PCs derived from gene expression data, and 10 PCs derived from the DNA methylation data).

We corrected the test-statistics for bias and inflation (estimated based on the association between DNA methylation and expression levels of all genes, using the R package *bacon*^27^).

### Enrichment analyses

#### Gene Set Analysis

GO and KEGG enrichment analyses were performed using the *methylgometh* function in the *methylGSA* R package^37,99^. This method takes into account that the number of CpGs assigned to each gene differs by accounting for the probability of a gene being selected using Wallenius’ noncentral hypergeometric distribution. We restricted the analysis to GO/KEGG categories that included at least 10 and at most 500 genes. All tested probes were used as background, and the *P*-values were adjusted for the number of categories tested using the Benjamin-Hochberg method. We considered both the default threshold used in the *methylGSA* package (*P* < 0.001) and the stringent genome-wide significance threshold (9 x 10^−8^) to select DMPs for enrichment analyses. Enrichment analyses were performed using both nearest gene annotations as well as eQTM annotations. In the eQTM enrichment analyses we restricted the analysis to sites that had a significant eQTM-association after accounting for multiple testing using a two-step approach as described previously^100^. This resulted in 79,441 eQTM-annotated sites that were considered for the enrichment analysis.

#### EWASdb enrichments

We tested whether the identified sites significantly overlapped with CpG-sites associated with other traits using publicly available EWAS databases (EWASdb^39^ and the MRC-IEU EWAS Catalog^38^). For each trait present in the EWAS databases, we overlapped the trait-associated CpG-sites with the ALS-associated CpG-sites. Fisher’s exact test was used to test for a significant enrichment (*FDR <* 0.05) relative to a background consisting of all probes included in the database. The resulting *P*-values were adjusted for the number of traits tested using the Benjamin-Hochberg method. We limited the analysis to traits that were associated with more than 5 and less than 5,000 CpGs and that were studied in blood-related tissues. Since the great majority of the EWASs in these databases are based on the 450k array, we limited the analyses to 450k array probes.

The Louvain algorithm as implemented in the *igraph* R package was used to identify clusters among the significantly enriched traits^40,101^. We further identified independent clusters of traits by removing the probes linked to the most significant trait, followed by retesting for enrichment among the remaining probes. We repeated this algorithm, retaining traits that were nominally significant (*P <* 0.05), until at most one trait remained significant.

### Poly-methylation scores

#### Derivation of poly-methylation scores

We considered the following poly-methylation scores: smoking^14,42^, alcohol intake^11,14^, BMI^12,14^, HDL-c^14^, LDL-c^14^, total cholesterol^14^, CRP^15^, white blood cell proportions^102^ (CD8T-cells, CD4T-cells, NK-cells, Monocytes, Granulocytes, and B-cells), DNA methylation age^13,44,45^, and a collection of 27 immunological and neurological plasma proteins^16,103^.

The WBC proportions were estimated using the robust partial correlations (RBC) method as implemented in the *EpiDISH* R package. The Zhang *et al.* age predictions were calculated using the scripts provided by the authors (https://github.com/qzhang314/DNAm-based-age-predictor). The Horvath age predictions were calculated using the supplementary code^44^. The *EpiSmokEr* smoking PMS were calculated using the *EpiSmokEr R* package (https://github.com/sailalithabollepalli/EpiSmokEr). The Liu *et al.* alcohol PMS were calculated using the *dnamalci* R package (https://github.com/yousefi138/dnamalci)^41^.

For all other PMSs, we downloaded the respective coefficients and calculated the PMS by multiplying each coefficient by the respective DNA methylation levels and summing these values:

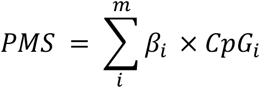

#### Validation of poly-methylation scores

Intermediate phenotypes were validated in a Dutch cohort of approximately 2000 individuals (2:1 case/control ratio) of which measured biomarker data was collected for 800 individuals (**supplementary table 4**). Collected biomarker data <1 week and phenotypic data < 52 weeks after the collection of blood for DNA methylation profiling were excluded from all analyses. Furthermore, CRP values >100 mg/L were considered an indicator of significant infection^104^ and were therefore not included for validation of the poly-methylation scores (PMS).

Linear regression models were used to identify the proportion of phenotypic variance explained by the corresponding PMS. Diagnosis, predicted age,sex, batch, cellcounts and 30 control probe PCs were considered as covariates in the null model, whereby the phenotypic measure was the dependent variable, if available:

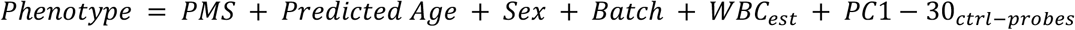

Incremental R² estimates were calculated between the null model and the models with the PMS of interest. The incremental R² estimates were used to determine whether the PMS increased the predictive ability above and beyond that provided by an existing model. Therefore, our analysis was limited to PMS with an incremental R² of at least 0.05 when intermediate phenotype/biomarker data was available.

Intermediate phenotypes/biomarker data were available for age (years), cigarettes (self-reported number of cigarettes in a year), alcohol (self-reported units of alcohol a week), Granulocytes (absolute counts), Monocytes (absolute counts), BMI (kg/m2), CRP (mg/L), total cholesterol (mmol/l), LDL-cholesterol(mmol/l), HDL-cholesterol (mmol/l).

#### Association testing

For each stratum, we tested for an association between the PMS and case/control status using logistic regression, the following regression models were used for HDL-c, BMI, alcohol intake, smoking, CRP and the immunological/neurological proteins:

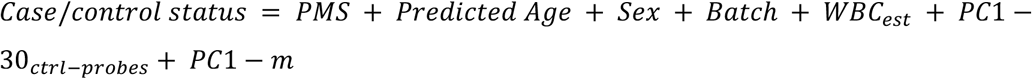

Where *m* = 30 for the MinE 450k stratum, *m* = 15 for the MinE EPIC stratum, *m* = 25 for the AUS2 stratum and *m* = 30 for the AUS2 stratum.

for DNA methylation age we additionally adjusted for age at blood draw, thus representing age acceleration^82^:

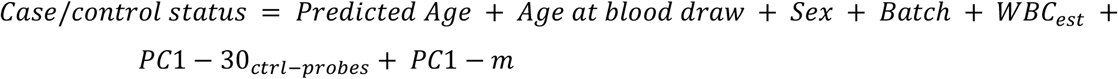

Where *m* = 30 for the MinE 450k stratum, *m* = 15 for the MinE EPIC stratum, *m* = 25 for the AUS1 stratum and *m* = 30 for the AUS2 stratum.

For the white blood cell proportions we did not adjust for array-wide principal components, given that the top principal components essentially represent white blood cell proportions^87^:

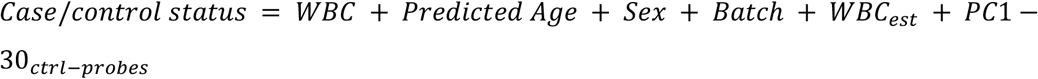

Strata test-statistics were combined using an inverse-variance-weighted, fixed-effects meta-analysis using the *metagen* function in the *meta* R package^29^. We corrected for the number of PMSs tested using the Bonferroni correction (0.05/39 = *P <* 1.3 × 10^−3^).

#### Survival

We used a multivariate Cox proportional hazards regression model to test for an association between survival and PMSs, adjusting for predicted age, sex, batch, estimated white blood cell proportions, 30 control probe PCs and a variable number of principal components, as described in the ‘Principal components + *bacon’* section.

### Survival analyses

#### Association testing

To investigate whether PMSs and the 45 significant sites were associated with survival, we applied multivariate Cox proportional hazards (PH) regression models utilizing the coxph function of the *survival* R package. Individuals with known overall survival, defined as the time from the date of a patient’s onset to the date of death or last known follow-up were included (450k: N = 2,892, EPIC: N = 2,363). Overall survival in months was defined as outcome of interest in the multivariate Cox PH model.

For the individual sites, we used multivariate models and corrected for bias and inflation similar as described in the ‘Principal components + *bacon’* section. Significant sites from the MOA and LB model were selected for survival analysis.

For the PMSs, we used multivariate models as described in the ‘Poly-methylation Scores’ section.

The proportional hazards (PH) assumption of the Cox model was checked using Schoenfeld and martingale residuals. Test-statistics were combined using inverse variance-weighted fixed effects meta-analysis. Sites were considered significantly heterogeneous when Cochran’s Q P-values < 0.1 (corrected for the number of tests performed).

#### Sensitivity analyses

Sensitivity analyses were performed to assess robustness of the Cox PH output after recognition of a violation in the proportional hazards assumptions. We applied two sensitivity analyses proposed to detect and model such time-varying effects ^105,106^.

First, the multivariate cox model, as described above, was stratified by batch allowing the underlying hazard function to vary across the experimental batch levels. Moreover, the variable predicted age was added as a log transformed time-varying covariate.

Second, flexible survival regression using the Royston-Parmar (RP) spline model was performed utilizing the flexsurvspline function from the R package *flexsurv*. This model utilizes restricted cubic splines permitting the estimation of a continuous function instead of a step function which takes into account time varying issues encountered when using the Cox PH. Model complexity was assessed by the addition of up to five knots compared to one single knot. Lastly, we evaluated the predictive effect of *C9orf72* carrier status on survival by adjusting for C9 in the multivariate model described above.

## Data Availability

All summary statistics are available as supplementary data. Raw data will become available after peer review.

https://bigd.big.ac.cn/ewas

http://www.ewascatalog.org/

https://www.bbmri.nl/acquisition-use-analyze/bios

http://geneontology.org/

https://www.genome.jp/kegg/pathway.html

http://www.godmc.org.uk/

## Acknowledgments

E.H. and J.M. were supported by Medical Research Council (MRC) grant K013807 (awarded to J.M.).

G.S. was supported by a PhD studentship from the Alzheimer’s Society.

W.v.R. is supported by funding provided by the Dutch Research Council (NWO) [VENI scheme grant 09150161810018] and Prinses Beatrix Spierfond (neuromuscular fellowship grant W.F19-03).

J.J.F.A.v.V. and J.H.V. acknowledge the Prinses Beatrix Spierfonds (W.OR20-08) for funding.

A.A.K. is funded by The Motor Neurone Disease Association (MNDA) and NIHR Maudsley Biomedical Research Centre.

A.N.B. is grateful to Suna and Inan Kirac Foundation and Koc University for the excellent research environment created and for their financial support.

G.A.R. is supported by the Canadian Institutes of Health.

J.P.R. is funded by the Canadian Institutes of Health Research (FRN 159279).

R.J.P. is funded through the Gravitation program of the Dutch Ministry of Education, Culture, and Science and the Netherlands Organization for Scientific Research (BRAINSCAPES).

V.S. is supported by the Italian Ministry of Health, AriSLA, and E-Rare Joint Transnational Call.

K.P.K. is supported by funding provided by the Dutch Research Council (NWO) [VIDI grant 91719350].

D.B., E.T. and H.R. are employees of Biogen.

L.H.v.d.B. reports grants from the Netherlands ALS Foundation, grants from The Netherlands Organization for Health Research and Development (Vici scheme), grants from The European Community’s Health Seventh Framework Programme (grant agreement n° 259867 (EuroMOTOR)), grants from The Netherlands Organization for Health Research and Development) the STRENGTH project, funded through the EU Joint Programme – Neurodegenerative Disease Research, JPND), during the conduct of the study. Several authors of this publication are members of the Netherlands Neuromuscular Center (NL-NMD) and the European Reference Network for rare neuromuscular diseases EURO-NMD. Project MinE Belgium was supported by a grant from IWT (n° 140935), the ALS Liga België, the National Lottery of Belgium and the KU Leuven Opening the Future Fund. PVD holds a senior clinical investigatorship of FWO-Vlaanderen and is supported by the E. von Behring Chair for Neuromuscular and Neurodegenerative Disorders, the ALS Liga België and the KU Leuven funds “Een Hart voor ALS”, “Laeversfonds voor ALS Onderzoek” and the “Valéry Perrier Race against ALS Fund”.

French ALS patients of the Pitié-Salpêtrière hospital (Paris) have been collected with ARSla funding support.

This work was supported by the Italian Ministry of Health (Ministero della Salute, Ricerca Sanitaria Finalizzata, grant RF-2016-02362405), the Progetti di Rilevante Interesse Nazionale program of the Ministry of Education, University and Research (grant 2017SNW5MB); the European Commission’s Health Seventh Framework Programme (FP7/2007–2013 under grant agreement 259867), and the Joint Programme–Neurodegenerative Disease Research (Strength, ALS-Care and Brain-Mend projects), granted by Italian Ministry of Education, University and Research. This study was performed under the Department of Excellence grant of the Italian Ministry of Education, University and Research to the “Rita Levi Montalcini” Department of Neuroscience, University of Torino, Italy.

We acknowledge funding from the Australian National Health and Medical Research (NHMRC) Council: 1151854, 1083187, 1173790, 1078901, 1113400, 1095215, 1176913 Enabling Grant #402703. Additional funding was provided by the Motor Neurone Disease Research Institute of Australia Ice Bucket Challenge grant for the SALSA-SGC consortium. The Older Australian Twins Study (OATS, used for controls) was facilitated through Twins Research Australia, a national resource in part supported by a Centre for Research Excellence from the Australian National Health and Medical Research Council (NHMRC 1079102). Funding for this study was awarded by the (NHMRC)/Australian Research Council Strategic Award (Grant 401162) and NHMRC grants (1405325, 1024224, 1025243, 1045325, 1085606, 568969, 1093083). We acknowledge the OATS research team: https://cheba.unsw.edu.au/project/older-australian-twins-study. We thank the participants and their informants for their time and generosity in contributing to this research.

This project has received funding from the European Research Council (ERC) under the European Union’s Horizon 2020 research and innovation programme (grant agreement n° 772376 - EScORIAL.

The collaboration project is co-funded by the PPP Allowance made available by Health∼Holland, Top Sector Life Sciences & Health, to stimulate public-private partnerships.

This study was supported by the ALS Foundation Netherlands.

This is an EU Joint Programme - Neurodegenerative Disease Research (JPND) project. The project is supported through the following funding organisations under the aegis of JPND - www.jpnd.eu (United Kingdom, Medical Research Council (MR/L501529/1; MR/R024804/1) and Economic and Social Research Council (ES/L008238/1)) and through the Motor Neurone Disease Association. This study represents independent research part funded by the National Institute for Health Research (NIHR) Biomedical Research Centre at South London and Maudsley NHS Foundation Trust and King’s College London. A.A-C. is supported by an NIHR Senior Investigator Award. Samples used in this research were entirely/in part obtained from the UK National DNA Bank for MND Research, funded by the MND Association and the Wellcome Trust. We would like to thank people with MND and their families for their participation in this project. We acknowledge sample management undertaken by Biobanking Solutions funded by the Medical Research Council at the Centre for Integrated Genomic Medical Research, University of Manchester.

## Author Contributions

### Sample ascertainment & data generation

P.J.H, R.A.J.Z, E.H, G.L.S, M.F.N, E.M.W, W.v.R, J.J.F.A.v.V, A.M.D, H-J.W, G.H.P.T, K.R.v.E, M.M, A.A.K, A.I, N.T, A.R, J.C-K, K.E.M, P.J.S, A.N.B, A.C, A. Calvo, C.M, A. Canosa, M.B, M.G, M. Gotkine, Y.L, M.Z, P.V’h, P.C, P. Couratier, J.S.M.P, T.S, P.D, J.P.R, R.D.H, S.M, P.A.M, M.N, G.N, D.B.R, R.P, K.A.M, P.S.S, S.F, F.C.G, A.K.H, T.L, S.T.N, F.J.S, L.W, K.L.W, B.C, B.M.C, M.M.N, R.J.C, I.P.B, M.C.K, V.D, M.P, M.d.C, S.P, M.W, G.R, V.S, J.E.L, C.E.S, P.M.A, A.F.M, M.A.v.E, R.J.P, N.R.W, R.L.M, O.H, K.P.K, A.A-C, L.H.v.d.B, P.V.D, J.M, J.H.V. **Whole-genome sequencing:** P.J.H, R.A.J.Z, W.v.R, J.J.F.A.v.V, A.M.D, G.H.P.T, K.R.v.E, M.M, J.C-K, K.P.K, A.A-C, L.H.v.d.B, P.V.D, J.H.V. **WGS quality-control:** P.J.H, R.A.J.Z, W.v.R, J.J.F.A.v.V, M.M, K.P.K, P.V.D, J.H.V. **Data analysis:** P.J.H, R.A.J.Z, E.H, J.M, J.H.V. **Writing the manuscript:** P.J.H, R.A.J.Z, J.M, J.H.V. **Revising manuscript:** P.J.H, R.A.J.Z, M.F.N, W.v.R, J.J.F.A.v.V, H-J.W, D.B, R.J.P, N.R.W, H.R, E.T, P.V.D, J.M, J.H.V.

## Consortia

### BIOS consortium (Biobank-based Integrative Omics Study)

**Management Team** Bastiaan T. Heijmans^62^ (Chair), Peter A.C. t Hoen^63^, Joyce van Meurs^64^, Rick Jansen^65^, Lude Franke^66^

**Cohort collection** Dorret I. Boomsma^67^, René Pool^67^, Jenny van Dongen^67^, Joukje J. Hottenga^67^ (Netherlands Twin Register); Marleen M.J. van Greevenbroek^68^, Coen D.A. Stehouwer^68^, Carla J.H. van der Kallen^68^, Casper G. Schalkwijk^68^ (Cohort study on Diabetes and Atherosclerosis Maastricht); Cisca Wijmenga^66^, Lude Franke^66^, Sasha Zhernakova^66^, Ettje F. Tigchelaar^66^ (LifeLines Deep); P. Eline Slagboom^62^, Marian Beekman^62^, Joris Deelen^62^, Diana van Heemst^69^ (Leiden Longevity Study); Jan H. Veldink^1^, Leonard H. van den Berg^1^ (Prospective ALS Study Netherlands); Cornelia M. van Duijn^70^, Bert A. Hofman^71^, Aaron Isaacs^70^, André G. Uitterlinden^64^ (Rotterdam Study)

**Data Generation**, Joyce van Meurs^64^ (Chair), P. Mila Jhamai^64^, Michael Verbiest^64^, H. Eka D. Suchiman^62^, Marijn Verkerk^64^, Ruud van der Breggen^62^, Jeroen van Rooij^64^, Nico Lakenberg^62^

**Data management and computational infrastructure** Hailiang Mei^72^ (Chair), Maarten van Iterson^62^, Michiel van Galen^63^, Jan Bot^73^, Dasha V. Zhernakova^66^, Rick Jansen^65^, Peter van ’t Hof^72^, Patrick Deelen^66^, Irene Nooren^73^, Peter A.C. t Hoen^63^, Bastiaan T. Heijmans^62^, Matthijs Moed^62^

**Data Analysis Group** Lude Franke^66^ (Co-Chair), Martijn Vermaat^63^, Dasha V. Zhernakova^66^, René Luijk^62^, Marc Jan Bonder^66^, Maarten van Iterson^62^, Patrick Deelen^66^, Freerk van Dijk ^74^, Michiel van Galen^63^, Wibowo Arindrarto^72^, Szymon M. Kielbasa^75^, Morris A. Swertz^74^, Erik W. van Zwet^75^, Rick Jansen^65^, Peter A.C. t Hoen^63^ (Co-Chair), Bastiaan T. Heijmans^62^ (Co-Chair)

### Affiliations

62: Molecular Epidemiology, Department of Biomedical Data Sciences, Leiden University Medical Center, Leiden, The Netherlands.

63: Department of Human Genetics, Leiden University Medical Center, Leiden, The Netherlands.

64: Department of Internal Medicine, ErasmusMC, Rotterdam, The Netherlands.

65: Department of Psychiatry, VU University Medical Center, Neuroscience Campus Amsterdam, Amsterdam, The Netherlands.

66: Department of Genetics, University of Groningen, University Medical Centre Groningen, Groningen, The Netherlands.

67: Department of Biological Psychology, VU University Amsterdam, Neuroscience Campus Amsterdam, Amsterdam, The Netherlands.

68: Department of Internal Medicine and School for Cardiovascular Diseases (CARIM), Maastricht University Medical Center, Maastricht, The Netherlands.

69: Department of Gerontology and Geriatrics, Leiden University Medical Center, Leiden, The Netherlands.

70: Department of Genetic Epidemiology, ErasmusMC, Rotterdam, The Netherlands. 71: Department of Epidemiology, ErasmusMC, Rotterdam, The Netherlands.

72: Sequence Analysis Support Core, Department of Biomedical Data Sciences, Leiden University Medical Center, Leiden, The Netherlands.

73: SURFsara, Amsterdam, the Netherlands.

74: Genomics Coordination Center, University Medical Center Groningen, University of Groningen, Groningen, the Netherlands.

75: Medical Statistics, Department of Biomedical Data Sciences, Leiden University Medical Center, Leiden, The Netherlands.

### BRAIN MEND Consortium

#### Biological Resource Analysis to Identify New MEchanisms and phenotypes in Neurodegenerative Diseases consortium

Ammar Al-Chalabi^9, 61^, Naomi R. Wray^3, 45^, Gilbert Bensimon^45, 76^, Orla Hardiman^60^, Adriano Chiò^19^, Jan H. Veldink^1^, George Davey Smith^77, 78^, Jonathan Mill^2^

### Affiliations

76: Département de Pharmacologie Clinique, Hôpital de la Pitié-Salpêtrière, UPMC Pharmacologie, AP-HP, Paris, France.

77: MRC Integrative Epidemiology Unit, University of Bristol, Bristol BS8 1TH, UK. 78: Population Health Science, Bristol Medical School, Bristol, Bristol BS8 1TH, UK.

