## Supplementary information for "Genome-wide study of DNA methylation in Amyotrophic Lateral Sclerosis identifies differentially methylated loci and implicates metabolic, inflammatory and cholesterol pathways"

#### Supplementary Data

**Supplementary Information** 2
- Cohort descriptions 2
- Description of cohorts in Project MinE whole-genome sequencing. 2

**Supplementary Tables** 6

**Supplementary Figures** 11

**References** 35

### Supplementary Information

#### Cohort descriptions

#### Description of cohorts in Project MinE whole-genome sequencing.

**The Netherlands (Veldink, van den Berg)**. ALS patients were diagnosed with ALS at the tertiary referral clinic for motor neuron disease at the University Medical Center Utrecht (Dutch ALS Center) or were included in the Prospective ALS Study in The Netherlands. Patients were not pre-screened for any mutations related to ALS. Control subjects were population based controls matched for sex, age and geographic region within the Netherlands. The Prospective ALS study in The Netherlands has been described in detail previously[1]. All participants gave written informed consent and the University Medical Center Utrecht Medical Ethics Committee, Utrecht, Netherlands approved this protocol.

**UK MNDA Biobank (C. Shaw, P. Shaw, Al-Chalabi, Morrison)**. Cases were diagnosed with ALS in one of 20 UK hospitals by neurologists specialized in motor neuron diseases. Patients had no family history for ALS. All participated in the UK National Biobank for Motor Neuron Disease Research. Patients had no family history for ALS and were of self-reported European descent. All participants gave written informed consent and the Trent University Medical Ethics Committee and Yorkshire and the Humber - Sheffield Research Ethics Committee approved this protocol.

**Turkey (Basak)**. ALS patients were recruited from hospitals across Turkey between 2002 and 2019. DNA samples were collected at the Boğaziçi University. A full description is provided in reference ref. [2]. All participants gave written informed consent and the Ethics Committee on Research with Human Participants (INAREK) at Bogazici University, Istanbul, Turkey approved this protocol.

**Belgium (van Damme).** Patients were diagnosed with ALS at the tertiary referral clinic for motor neuron diseases at the University Hospitals in Leuven. Patients were pre-screened for mutations in *C9orf72*, *SOD1*, *TARDBP* and *FUS*, but were not always excluded in case of pathogenic mutation[3]. Control subjects were often spouses of patients, supplemented with age- and sex-matched controls from other local studies[4]. All participants gave written informed consent and the Ethical Committee of University Hospital Leuven approved this protocol.

**Ireland (Hardiman, McLaughlin).** Cases were diagnosed with probable or definite ALS according to the 1994 El-Escorial Criteria by neurologists specialized in motor neurone diseases at Beaumont Hospital in Dublin. Patients were referred from all regions in Ireland and were part of an ongoing population-based prospective ALS registry. Patients were selected for sequencing such that all areas of Ireland were adequately represented. Control samples were neurologically healthy volunteers matched for geography, sex and age sampled from the community. All individuals reported Irish ancestry for at least three generations. All participants gave written informed consent and the Beaumont Hospital Research & Ethics Committee approved this protocol.

**Spain (Mora Pardina, Povedano).** ALS patients diagnosed with definite or probable ALS according to the El Escorial criteria. Patients were seen by neurologists and neurophysiologists at the tertiary referral centers: the Bellvitge hospital and Carlos III hospital for Catalonia and Madrid respectively. Controls were healthy individuals, without familial history of ALS, matched for age and sex. All participants gave written informed consent and the Comité de Ética de la Investigación del Hospital Carlos III and Bellvitge University Hospital Ethics Committee, Barcelona, Spain approved this protocol.

**United States (Landers, Glass).** All samples were taken from patients seen at the Emory ALS Center in Atlanta, Georgia, USA. The Emory Center is a tertiary care ALS clinic caring for a large proportion of patients in Georgia and surrounding states[5]. Diagnoses were made by neurologists specialized in neuromuscular diseases and motor neuron diseases. After informed consent, full demographic and clinical information was stored into the clinic database. DNA was collected and stored. All participants gave written informed consent and the Committee for the Protection of Human Subjects in Research of the University of Massachusetts Medical School, Worcester, USA approved this protocol.

**France (Corcia, Couratier, Vourc'h).** ALS patients were diagnosed with probable or definite ALS according to the El Escorial Criteria by neurologists specialized in motor neuron diseases at the Reference centers for ALS of the University Hospitals of Limoges and Tours (LITORALS federation), members of the French FILSLAN networks. All participants gave written informed consent and the Medical Research Ethics Committee of “Assistance Publique-Hôpitaux de Paris”, The ethics committee of Tours Hospital, France, and Ethics committee of Limoges University Hospital, France approved this protocol.

**Sweden (Andersen).** Cases were diagnosed with probable or definite ALS according to the revised El-Escorial Criteria by neurologists specialized in motor neuron diseases. Control individuals were free of any neuromuscular disease and matched for age, gender and ethnicity. Healthy controls were spouses of ALS patients or patients with other neurological diseases. All participants were from Swedish descent all reporting Northern Swedish citizenship for at least three generations. All participants gave written informed consent and the Regional Ethical Review Board in Umeå approved this protocol.

**Israel (Gotkine, Drory)**. ALS patients were diagnosed with probable or definite ALS according to the El Escorial Criteria and in follow-up at the tertiary referral ALS clinic at the Hadassah University Hospital, Jerusalem or Tel-Aviv Sourasky Medical Center in Tel-Aviv. Patients were not pre-screened for any mutations related to ALS. Patients were referred from all regions in Israel and participated in a prospective ALS database and sample repository. All participants gave written informed consent and the Hadassah University Hospital IRB board and The Institutional Review Board of Tel Aviv Sourasky Medical Center, Israel approved this protocol.

**Portugal (deCarvalho, Pinto).** Patients were diagnosed with possible, probable or definite ALS according to the revised El-Escorial criteria by neurologists specialized in motor neuron diseases. Both cases with and without a family history (third degree relatives) were included. Control subjects were spouses or those accompanying patients to the clinic. All participants gave written informed consent and the The Local Research Ethics Committee at the Faculty of Medicine, University of Lisbon, Lisbon, Portugal approved this protocol.

**Italy (Chio, Ticozzi, Silani).** Patients were diagnosed with ALS according to the El Escorial revised criteria at the ALS tertiary referral center of Istituto Auxologico Italiano IRCCS. All patients had probable or definite familial ALS according to the Byrne criteria for FALS[6]. Patients were pre-screened for mutations in the *SOD1*, *TARDBP*, *FUS* and *C9orf72* genes. All participants gave written informed consent and the Ethical Committee of Città della Salute Hospital, Torino, Italy approved this protocol.

**Switzerland (Weber).** ALS patients were diagnosed at the Muskelzentrum/ALS clinic at the Kantonsspital St. Gallen, a tertiary referral center in Northern Switzerland. Patients fulfilled the El-Escorial Criteria for probable lab supported, probable or definite or ALS. Control subjects were healthy blood donors matched for age and gender. All participants gave written informed consent and the Kantonale Ethikkomission des Kantons St. Gallen, Switzerland approved this protocol.

**Australia (Wray)**. The Australian ALS cohort (AUS) and DNA methylation assays have been previously described elsewhere[7,8]. Part of the Australian sample comprised patients and controls that were ascertained from the University of Sydney as part of the Australian MND DNA bank, which recruited participants from April 2000 to June 2011. Cases were white Australians older than 25 years recruited from around Australia via state-based MND associations with diagnosis verified by a neurologist. Control individuals were either partners or friends of patients with ALS or community volunteers. The remainder of Australian cases were recruited from clinics across Australia between 2015 and 2017 diagnosed with definite or probable ALS according to the revised El Escorial criteria. Control subjects were healthy individuals free of neuromuscular diseases, recruited as either partners or friends of patients with ALS or community volunteers or from the Older Australian Twins Study (OATS)[9]. ALS cases with a recorded family history of ALS were excluded. All participants gave written informed consent and the protocol was approved by Sydney South West Area Health Service Human Research Ethics Committee; HREC at the different sites: University of Sydney, Western Sydney Local Health District, Royal Brisbane and Women Hospital Metro North, South Metropolitan Health Service, Macquarie University, QIMR Berghofer Medical Research Institute, University of New South Wales and the University of Melbourne. DNA methylation were measured using Illumina Infinium HumanMethylation450 BeadChip. These data are available at dbGAP phs002068.v1.p1.

### Supplementary Tables

**Supplementary table 1.** Thresholds applied for each QC metric in each stratum.

| Stratum | MU | RG ratio | GR ratio | OP | HC | bscon | detP | beadNr | XY diff | IBS mean | IBS var | additional |
|---|---|---|---|---|---|---|---|---|---|---|---|---|
| MinE 450k | 2000 | 0.5 | 0.5 | 11.75 | 12 | 80 | 0.05 | 0.05 | -2 | 1.9 | 0.1 | WGS inbreeding |
| MinE EPIC | 2000 | 0.4 | 0.4 | 12 | 12.75 | 80 | 0.05 | 0.05 | -2 | 1.9 | 0.1 | WGS inbreeding |
| AUS1 | 1000 | 0.35 | 0.5 | 11.75 | 13 | 80 | 0.05 | 0.05 | -2 | 1.9 | 0.1 | - |
| AUS2 | 1500 | 0.5 | 0.5 | 11.75 | 12 | 80 | 0.05 | 0.05 | -2 | 1.9 | 0.1 | - |

**Supplementary table 2.** Number of samples that failed QC across the different metrics based on the thresholds supplied in Supplementary table 1.

| Stratum | MU | RG ratio | GR ratio | HC | OP | bscon | bead Nr | detectionP | Sex check | WGS concordance (IBS) | Inbreeding failures (WGS) | Tissue (non-blood) | control PCs | Total |
|---|---|---|---|---|---|---|---|---|---|---|---|---|---|---|
| MinE 450k | 86 | 0 | 0 | 14 | 31 | 84 | 0 | 65 | 46 | 91 | 53 | 47 | 26 | 302 |
| MinE EPIC | 48 | 0 | 3 | 3 | 4 | 7 | 0 | 65 | 64 | 40 | 61 | - | 6 | 230 |
| AUS1 | 10 | 19 | 0 | 4 | 0 | 0 | 0 | 0 | 0 | - | - | - | 0 | 27 |
| AUS2 | 17 | 0 | 0 | 0 | 0 | 1 | 0 | 0 | 0 | - | - | - | 1 | 19 |

**Supplementary table 3.** Number of samples that were removed based on various metrics applied after the first phase of QC and normalization.

| Stratum | Age discordance | PCA | missing phenotype | sample swap | relatedness | Total |
|---|---|---|---|---|---|---|
| MinE 450k | 6 | 22 | 16 | 3 | 80 | 127 |
| MinE EPIC | 17 | 5 | 94 | 2 | 55 | 173 |
| AUS1 | 2 | 11 | 0 | 0 | 1 | 14 |
| AUS2 | 0 | 0 | 0 | 0 | 0 | 0 |

**Supplementary table 4.** Demographic and clinical characteristics of samples with intermediate phenotype/biomarker data.

| | mine_450k (N=2733) | mine_epic (N=313) |
|---|---|---|
| **Diagnosis** | | |
| Control | 1003 (37 %) | 125 (40 %) |
| Cases | 1730 (63 %) | 188 (60 %) |
| **Age (years)** | | |
| Mean (SD) | 64 (± 10) | 65 (± 12) |
| Missing | 33 (1.2%) | 0 (0%) |
| **Body mass index (kg/m2)** | | |
| Mean (SD) | 25 (± 4.0) | 25 (± 3.6) |
| Missing | 693 (25.4%) | 25 (8.0%) |
| **Cigarettes year survey** | | |
| Mean (SD) | 1.6 (± 5.2) | 1.2 (± 4.3) |
| Missing | 694 (25.4%) | 25 (8.0%) |
| **Units of alcohol year survey** | | |
| Mean (SD) | 6.6 (± 9.6) | 5.9 (± 8.6) |
| Missing | 689 (25.2%) | 25 (8.0%) |
| **CRP (mg/L)** | | |
| Mean (SD) | 3.7 (± 5.3) | 4.9 (± 13) |
| Missing | 1974 (72.2%) | 191 (61.0%) |
| **Total cholesterol (mmol/l)** | | |
| Mean (SD) | 5.5 (± 1.2) | 5.6 (± 1.1) |
| Missing | 2196 (80.4%) | 194 (62.0%) |
| **HDL cholesterol (mmol/l)** | | |
| Mean (SD) | 1.5 (± 0.38) | 1.4 (± 0.35) |
| Missing | 2196 (80.4%) | 194 (62.0%) |

| | | |
|---|---|---|
| **LDL cholesterol (mmol/l)** | | |
| Mean (SD) | 3.3 (± 0.97) | 3.5 (± 0.92) |
| Missing | 2208 (80.8%) | 194 (62.0%) |
| **Monocytes (absolute counts)** | | |
| Mean (SD) | 0.51 (± 0.18) | 0.49 (± 0.17) |
| Missing | 1869 (68.4%) | 190 (60.7%) |
| **Granulocytes (absolute counts)** | | |
| Mean (SD) | 4.9 (± 1.6) | 4.7 (± 1.7) |
| Missing | 1959 (71.7%) | 190 (60.7%) |

**Supplementary table 5.** Demographic and clinical characteristics of samples per experimental batch and excluded samples.
[Table in Excel file]

**Supplementary table 6.** Test statistics for all tested sites (LB algorithm).
[table in zipped csv file]

**Supplementary table 7.** Test statistics for all tested sites (OSCA MOA algorithm).
[table in zipped csv file]

**Supplementary table 8.** Associations (eQTMs) between the DNA methylation levels of significant sites and the expression of nearby genes (<250Kb).
[table in Excel file]

**Supplementary table 9** Details of the gene sets that were significantly enriched among the MOA and LB results based on 79,441 sites that had a significant eQTM association. Method = EWAS method and *p*-value cutoff applied to the respective EWAS test-statistics resulting in the input probes for the shown enrichment analyses, N overlap = Number of significant genes that overlap with genes in the respective pathway, N genes = Total number of genes in the pathway, *FDR* = FDR-controlled (False discovery rate) *P*-values.

| **Method** | **Database** | **Pathway** | **N overlap** | **N genes** | ***FDR*** |
|---|---|---|---|---|---|
| **LB ($p < 0.001$)** | - | - | - | - | - |
| **MOA ($p < 0.001$)** | - | - | - | - | - |
| **LB ($p < 9 \times 10^{-8}$)** | GO BP | steroid biosynthetic process | 4 | 96 | 0.042 |
| | GO BP | cholesterol biosynthetic process | 4 | 44 | 0.0047 |
| | GO BP | cholesterol metabolic process | 4 | 87 | 0.042 |
| | GO BP | sterol metabolic process | 4 | 95 | 0.045 |
| | GO BP | sterol biosynthetic process | 4 | 48 | 0.0047 |
| | GO BP | alcohol biosynthetic process | 4 | 91 | 0.042 |
| | GO BP | secondary alcohol metabolic process | 4 | 93 | 0.042 |
| | GO BP | secondary alcohol biosynthetic process | 4 | 44 | 0.0047 |
| **MOA ($p < 9 \times 10^{-8}$)** | KEGG | Steroid biosynthesis | 2 | 14 | 0.015 |
| | GO BP | steroid biosynthetic process | 3 | 96 | 0.028 |
| | GO BP | cholesterol biosynthetic process | 3 | 44 | 0.0067 |
| | GO BP | cholesterol metabolic process | 3 | 87 | 0.028 |
| | GO BP | sterol metabolic process | 3 | 95 | 0.030 |
| | GO BP | sterol biosynthetic process | 3 | 48 | 0.0067 |
| | GO BP | alcohol biosynthetic process | 3 | 91 | 0.028 |
| | GO BP | organic hydroxy compound biosynthetic process | 3 | 126 | 0.048 |

| | | | | |
|---|---|---|---|---|
| GO BP | secondary alcohol metabolic process | 3 | 93 | 0.028 |
| GO BP | secondary alcohol biosynthetic process | 3 | 44 | 0.0067 |

**Supplementary table 10.** EWASdb enrichment test-statistics for both the MRC and the NGDC EWAS databases, and for both the sites identified with the LB and OSCA MOA algorithm respectively.
[Table in Excel file]

**Supplementary table 11.** Overview of probe overlap with the significantly enriched traits from the MRC and NGDC EWAS databases.
[Table in Excel file]

**Supplementary table 12.** Poly-methylation score (PMS) test statistics.
[Table in Excel file]

**Supplementary table 13.** Test statistics of the 45 significant MOA and LB probes in association with survival (Cox PH) and various sensitivity analyses.
[Table in Excel file]

**Supplementary table 14.** Poly-methylation score (PMS) survival (Cox PH) test statistics.
[Table in Excel file]

### Supplementary Figures

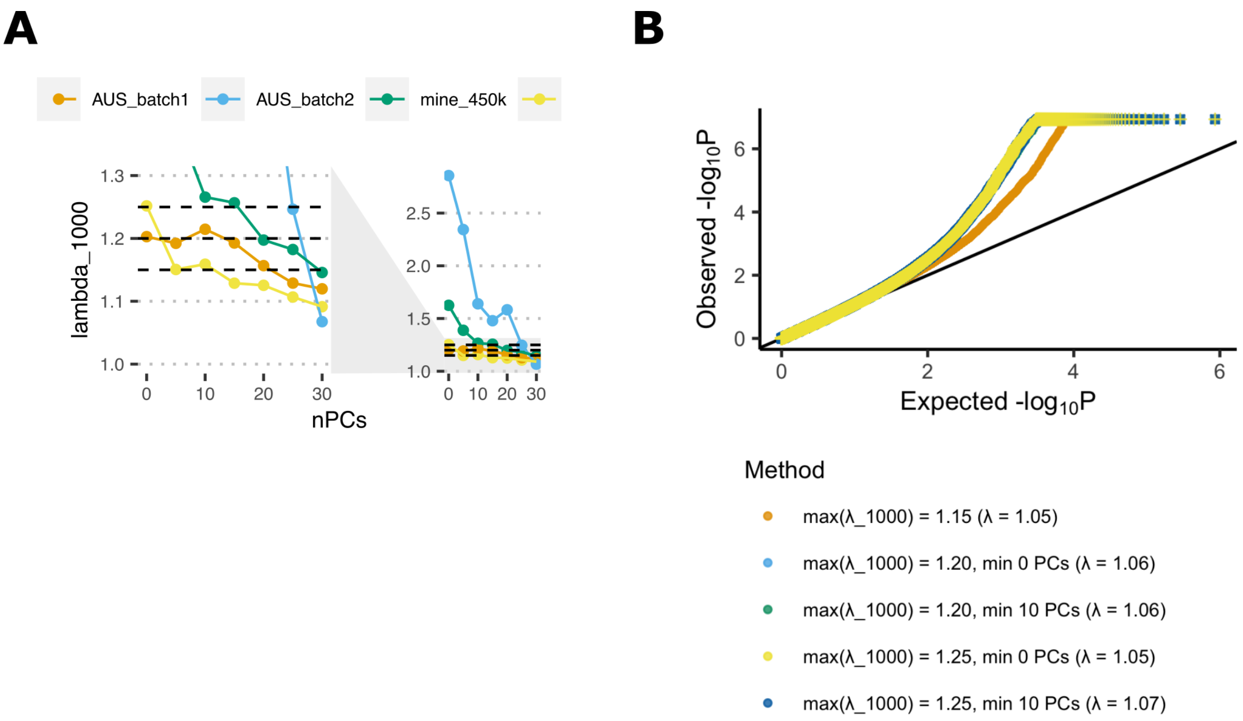


**Supplementary Figure 1.** Calibration of the number of principal components (PCs) included in the linear models.
**(A)** Sample-size normalized inflation factors ($\lambda_{1000}$; y-axis) against the number of PCs included (x-axis) in each stratum. In each model a core set of covariates including predicted age, sex, experimental batch and predicted white blood cell proportions are included. The left figure represents a zoomed-in version of the figure on the right.
**(B)** QQ-plots of the inverse variance-weighted fixed effects meta-analyses of the four strata for different thresholds of $\lambda_{1000}$ in the individual strata. *bacon*[10] was applied to the test-statistics of each stratum to correct for residual bias and inflation prior to performing the meta-analysis.

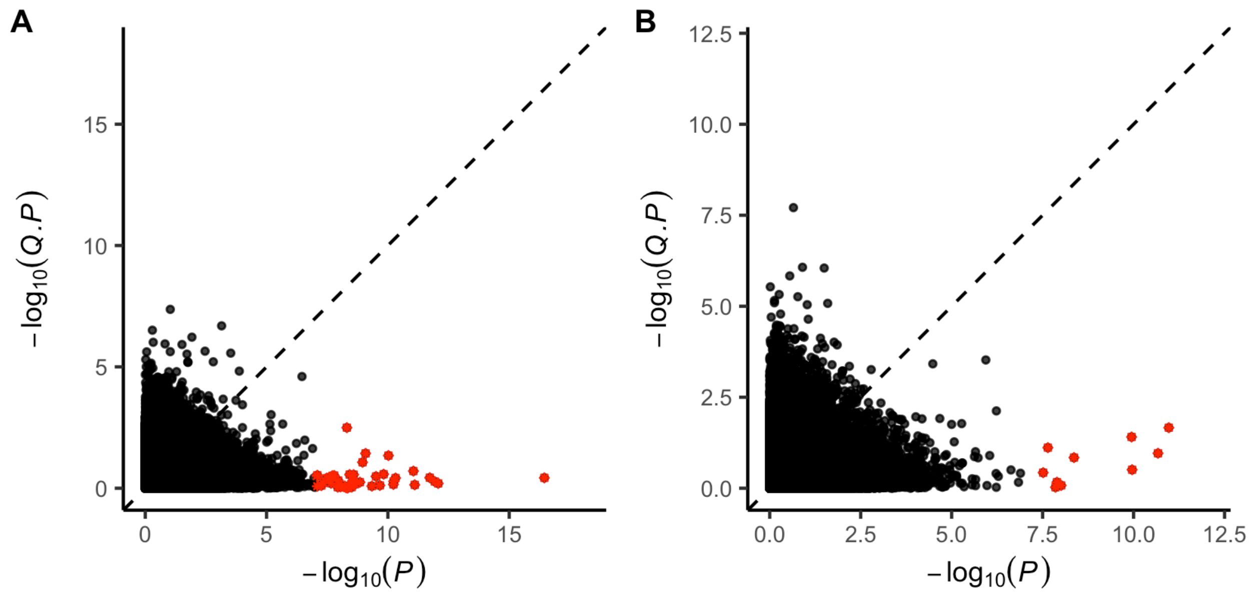


**Supplementary Figure 2.** Comparison of EWAS meta-analysis *P*-values with *P*-values from Cochran's Q test for heterogeneity. Differentially methylated sites (at $P < 9 \times 10^{-8}$) are highlighted in red. **(A)** LB meta-analysis test statistics, **(B)** MOA meta-analysis test statistics.

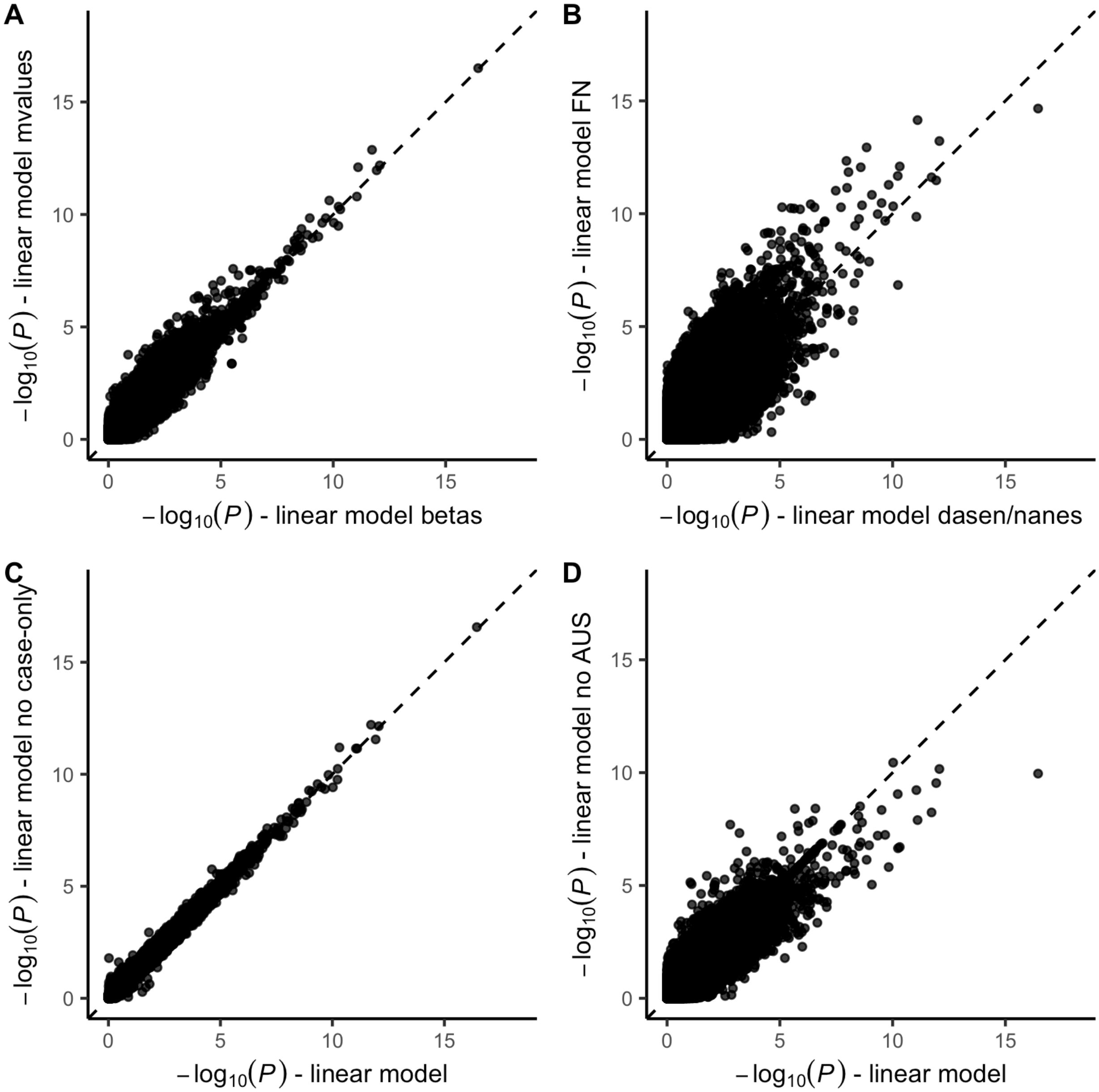


**Supplementary Figure 3.** EWAS sensitivity analyses for LB results.
**(A)** Test-statistics using M-values (y-axis) compared to $\beta$-values (x-axis).
**(B)** Test-statistics after normalizing the data using functional normalization (y-axis) instead of *dasen* (and *nanes for* the AUS1 stratum) (x-axis).
**(C)** Test-statistics after removing all experimental batches that contain only patients (y-axis), compared to the original test-statistics (x-axis).
**(D)** Test-statistics excluding the non-Project MinE data (y-axis), compared to the original test-statistics including all data (x-axis).

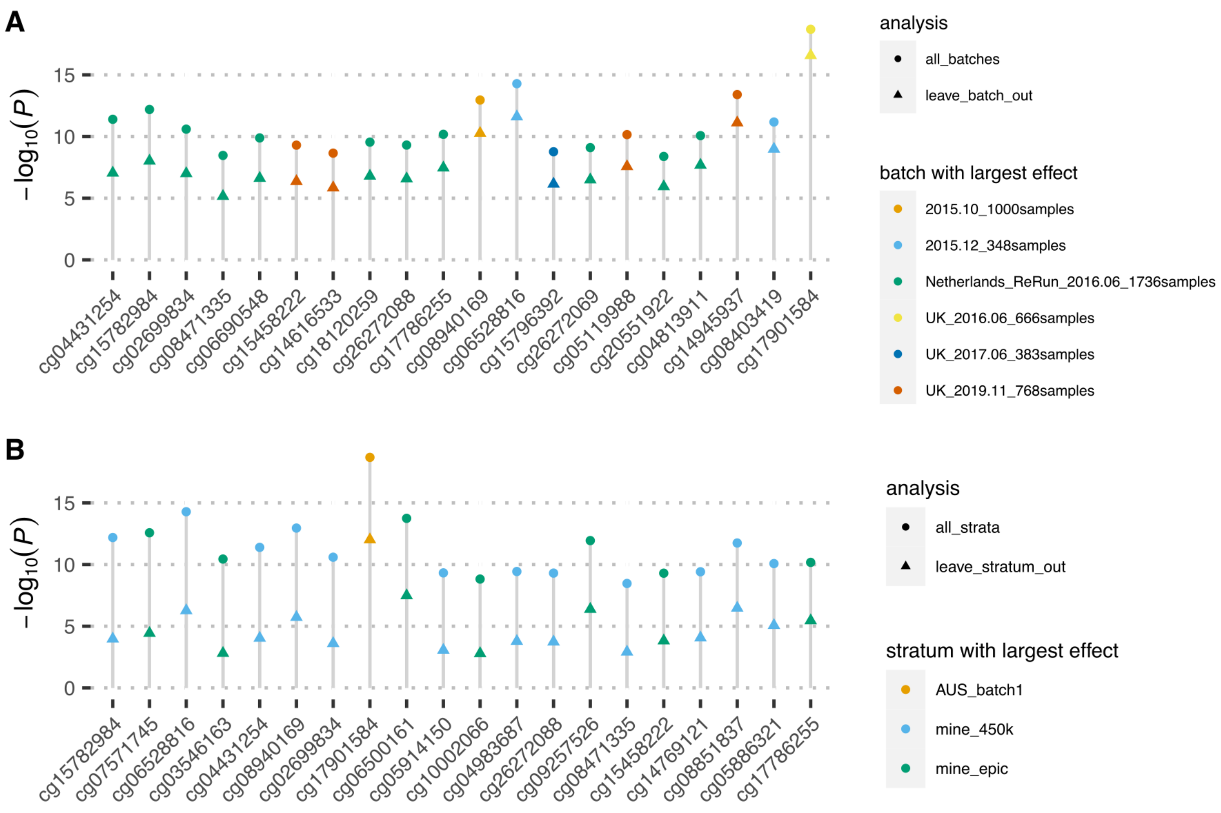


**Supplementary Figure 4.** Leave-one-out analyses testing the influence of specific experimental batches and strata on the results.
**(A)** 20 probes showing the largest *P*-value difference after leaving out one experimental batch. Dots indicate the *P*-value including all experimental batches, triangles indicate the *P*-value after leaving out the experimental batch indicated by the color. The color indicates the experimental batch that had the largest effect on exclusion. Note that *P*-values were not corrected for bias and inflation using *bacon*.
**(B)** 20 probes showing the largest *P*-value difference after leaving out one stratum. Dots indicate the *P*-value including all strata, triangles indicate the *P*-value after leaving out the stratum indicated by the color. The color indicates the stratum that had the largest effect on exclusion. Note that *P*-values were not corrected for bias and inflation using *bacon*.

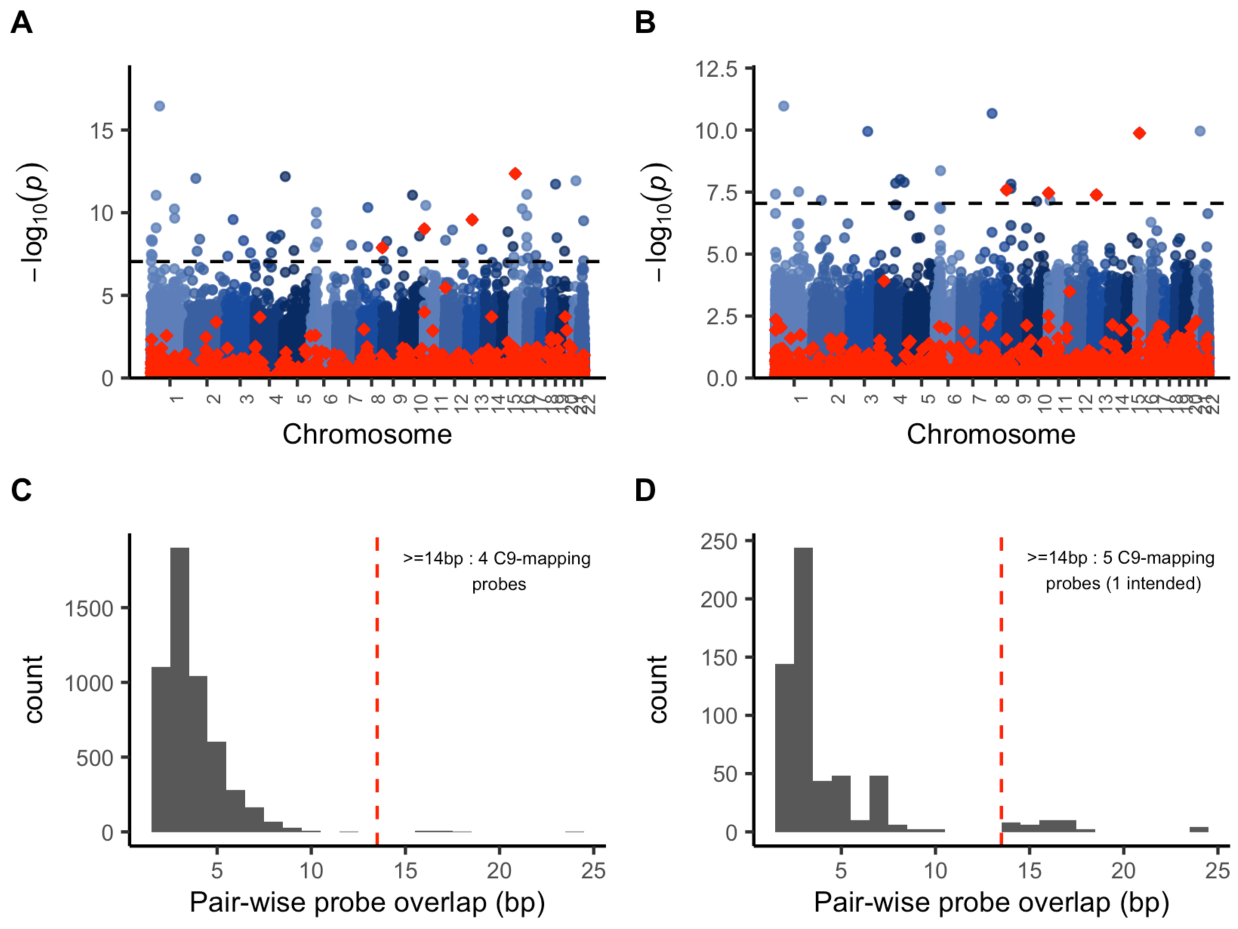


**Supplementary Figure 5. (A)** LB meta-analysis test statistics, probes mapping to the C9 repeat (≥14bp inexact match) are highlighted in red. **(B)** MOA meta-analysis test statistics, probes mapping to the C9 repeat (≥14bp inexact match[11]) are highlighted in red. **(C)** Sequence overlap among significant linear+bacon sites, probes with ≥15bp overlap all map to the C9 repeat. **(D)** Sequence overlap among significant MOA sites, probes with ≥15bp overlap all show homology to the C9 repeat, including one probe (cg01589155) for which the C9 repeat is the intended location.

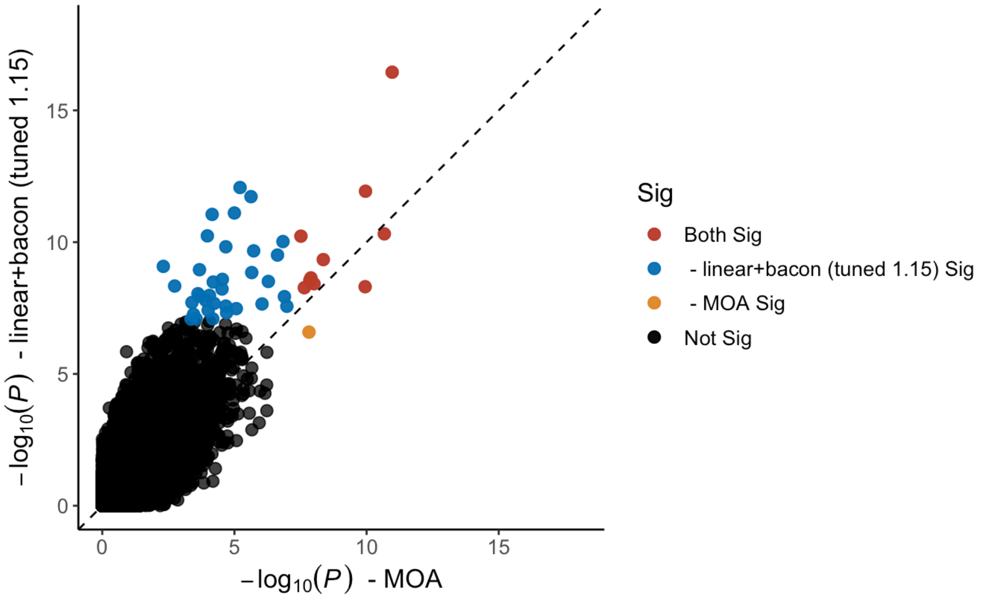


**Supplementary Figure 6.** Comparison of MOA meta-analysis *P*-values (x-axis) with LB meta-analysis *P*-values (y-axis).

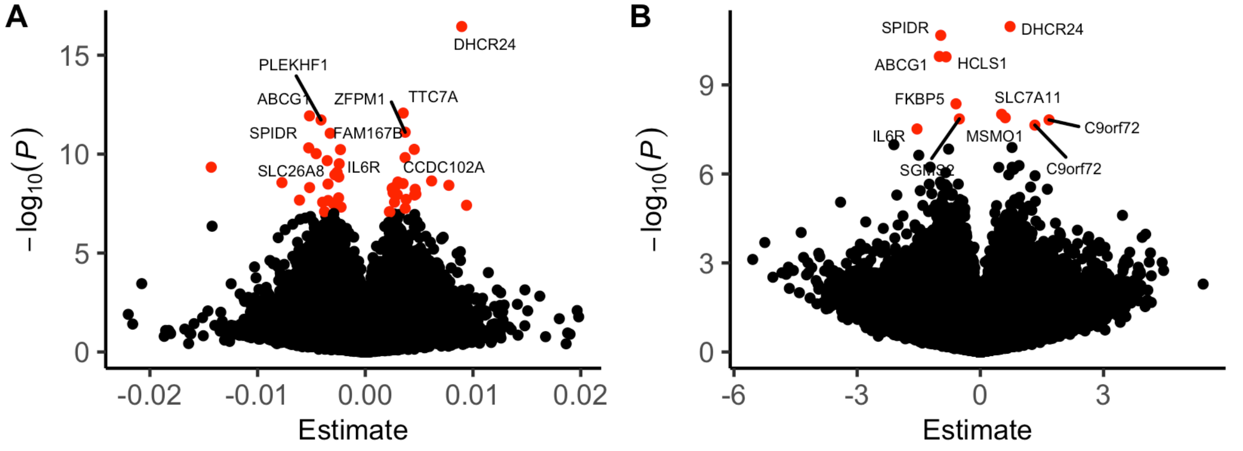


**Supplementary Figure 7.** Volcano plots showing the estimated effect sizes (x-axis) and -$\log_{10}(P)$ (y-axis). **(A)** LB meta-analysis test statistics. Nearest genes are shown for sites where -$\log_{10}(P)$ > 10. **(B)** MOA meta-analysis test statistics. Nearest genes are shown for the significant sites. Red dots indicate sites that passed the genome-wide significance threshold ($9 \times 10^{-8}$).

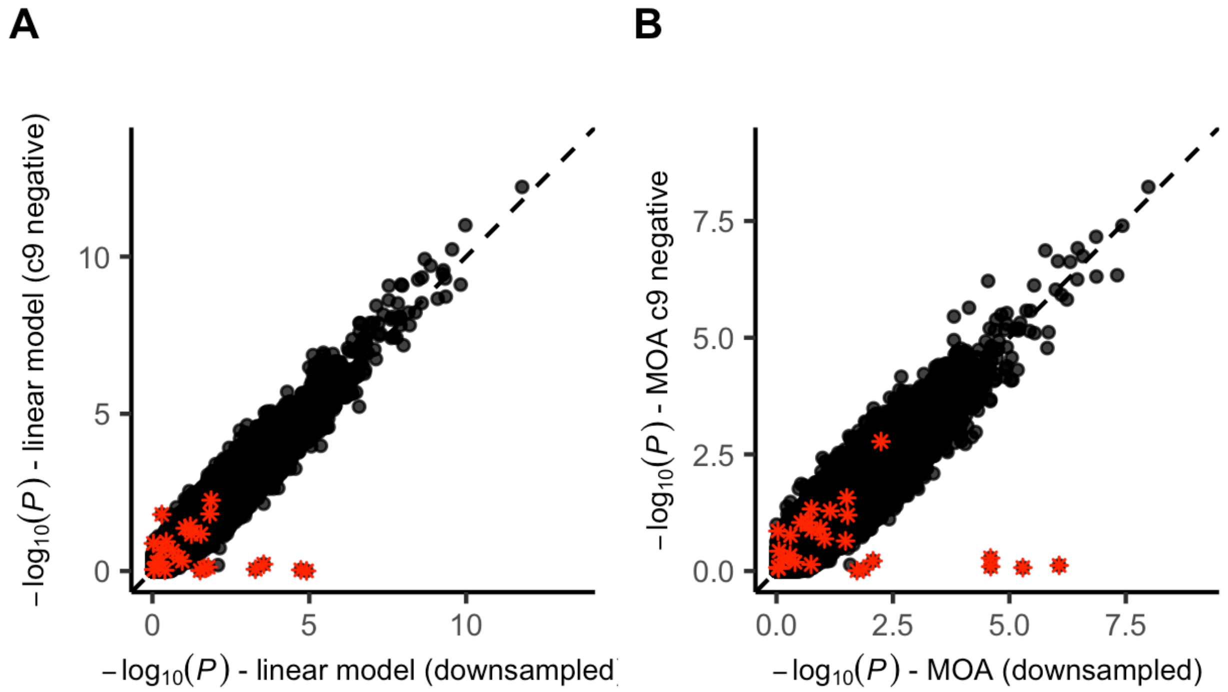


**Supplementary Figure 8.** Sensitivity analyses comparing an EWAS where we excluded individuals carrying the C9 repeat expansion (y-axis), compared to an EWAS including C9-carriers where we randomly downsampled the number of samples to match the sample size of the C9-negative EWAS (x-axis). **(A)** LB test-statistics, **(B)** MOA test-statistics. The red asterisks indicate sites that are located within the *C9orf72* gene.

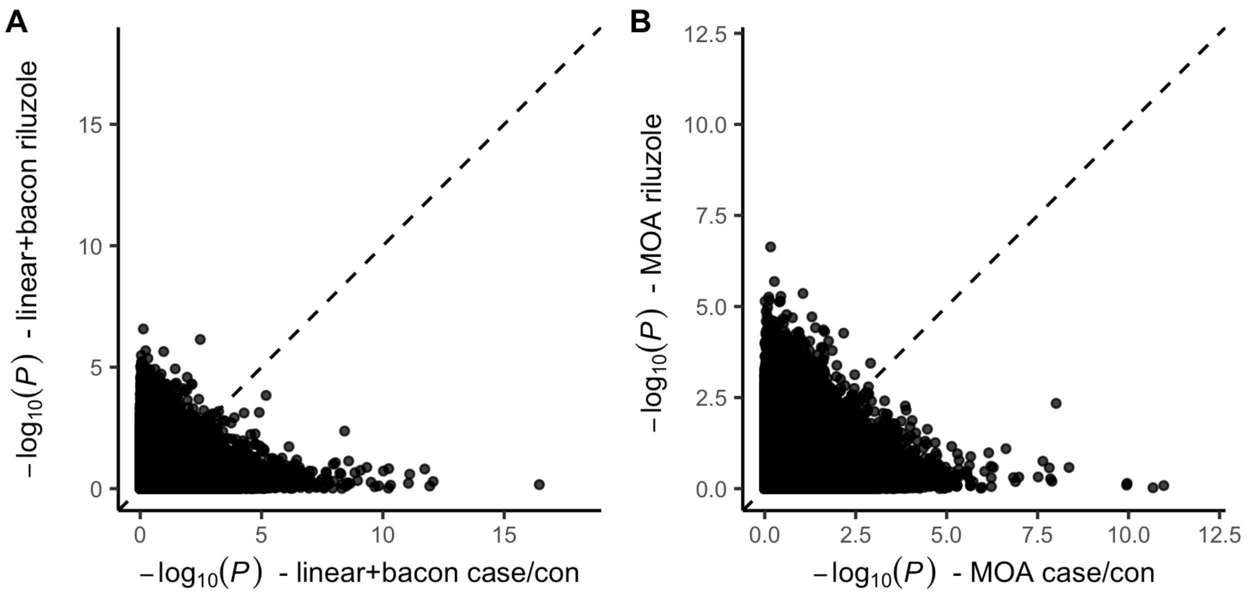


**Supplementary Figure 9.** *P*-values from an EWAS on riluzole usage (y-axis) compared to case/control *P*-value (x-axis). **(A)** LB test-statistics, **(B)** MOA test-statistics.

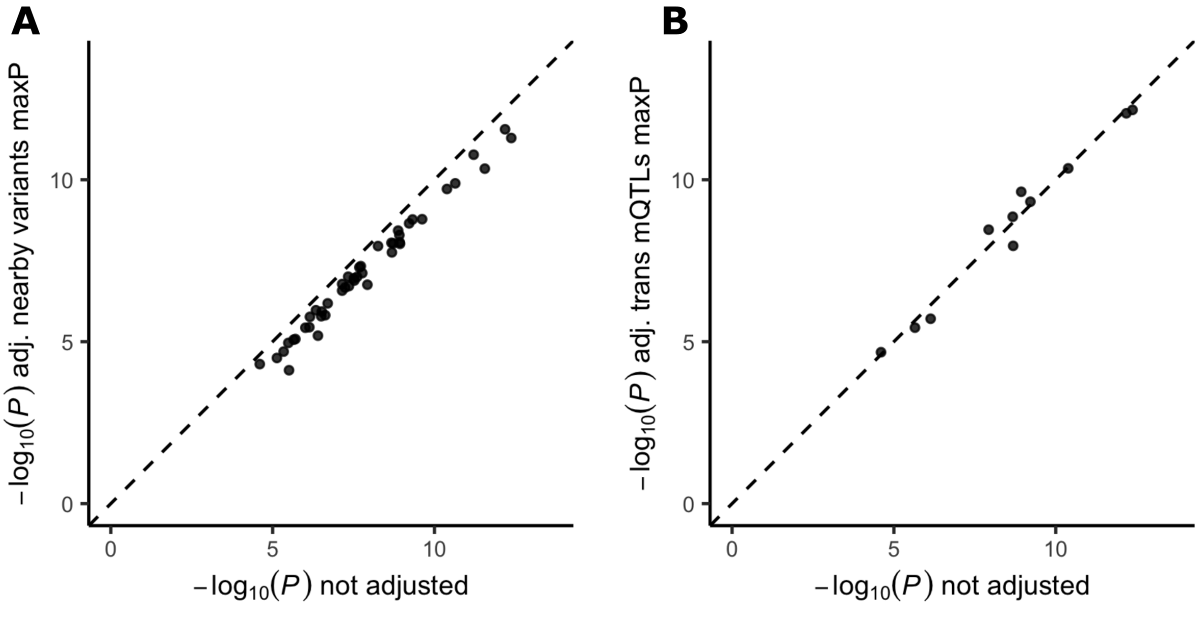


**Supplementary Figure 10.** Maximum P-value observed among iterative tests including one variant at a time on top of the core set of covariates included in the LB algorithm (y-axis), compared to the P-value including only the core set of covariates in all samples for which WGS data was available (x-axis). **(A)** Iteratively including all measured variants *in cis* (<250 Kb); **(B)**

Iteratively including *trans*-mQTLs as reported in the GoDMC consortium[12] (for 11 DMPs at least one trans-mQTL was reported).

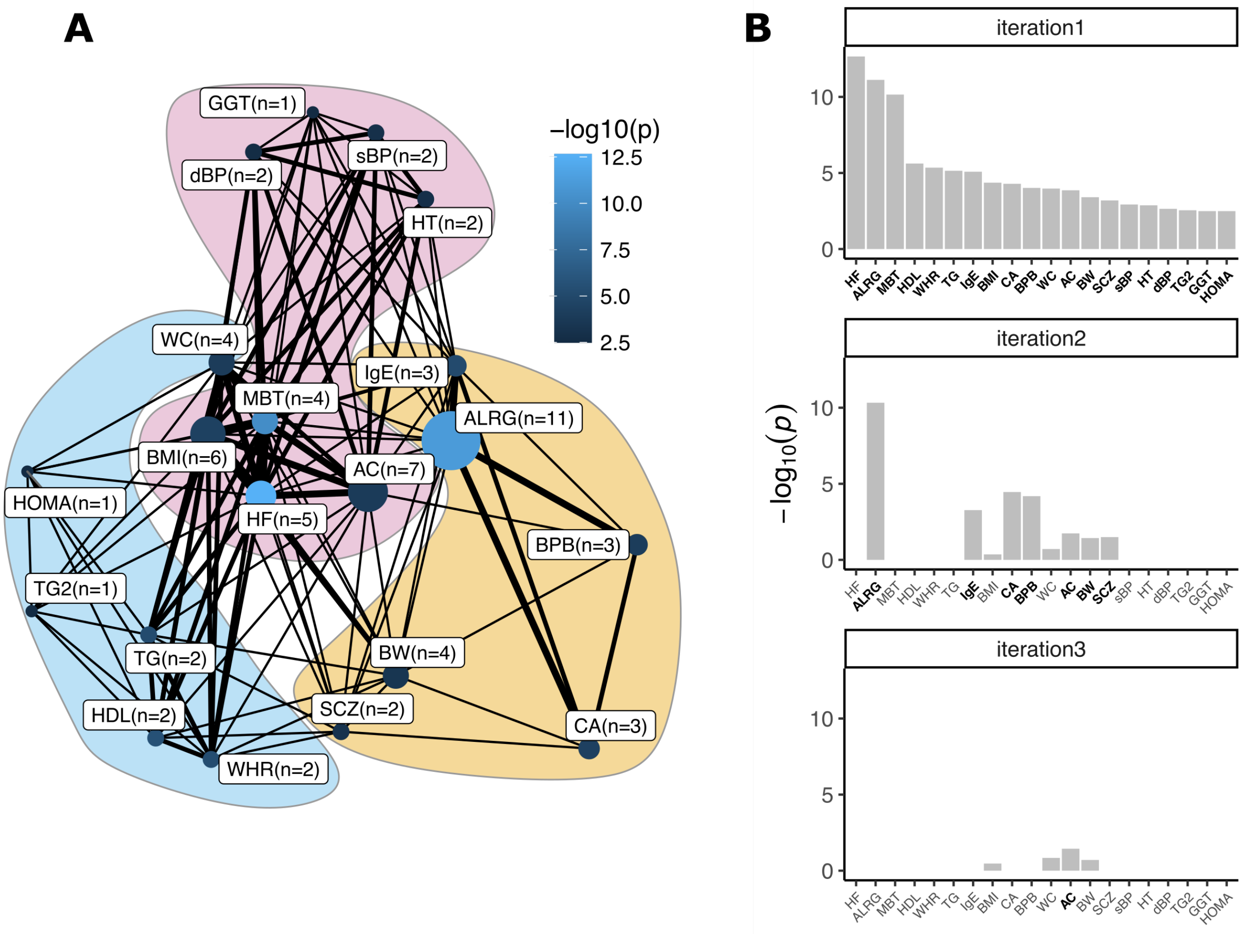


**Supplementary Figure 11.** Significant overlap between traits included in the NGDC EWAS database[13] and ALS-associated sites identified using the LB algorithm. **(A)** Network showing the traits that significantly overlap with the ALS-associated sites. Edges indicate probe overlap between the traits, with thicker lines indicating more overlapping probes.The node color indicates the enrichment -$\log_{10}(P)$, with lighter shades of blue indicating stronger associations. Colored surfaces indicate the clusters identified using the Louvain clustering algorithm. **(B)** Identification of independent clusters of traits. The first iteration shows the traits that significantly overlap with the ALS-associated probes at *FDR* < 0.05. In subsequent iterations the probes belonging to the most significant trait were excluded and enrichments tests were performed using the remaining traits. This algorithm was repeated, retaining traits that were nominally significant ($P < 0.05$, indicated in bold), until at most one trait remained significant.

Abbreviations: HF = hepatic fat, ALRG = allergic sensitization, MBT = metabolic trait, WHR = waist to hip ratio, CA = childhood asthma, BPB = blood protein biomarker levels, WC = waist circumference, AC = alcohol consumption, BW = birth weight, SCZ = schizophrenia, sBP = systolic blood pressure, HT = hypertension, dBP = diastolic blood pressure, TG2 = triglyceride postprandial responses to a high-fat dietary challenge, GGT = serum gamma-glutamyl transferase, HOMA = plasma fasting HOMA-IR levels.

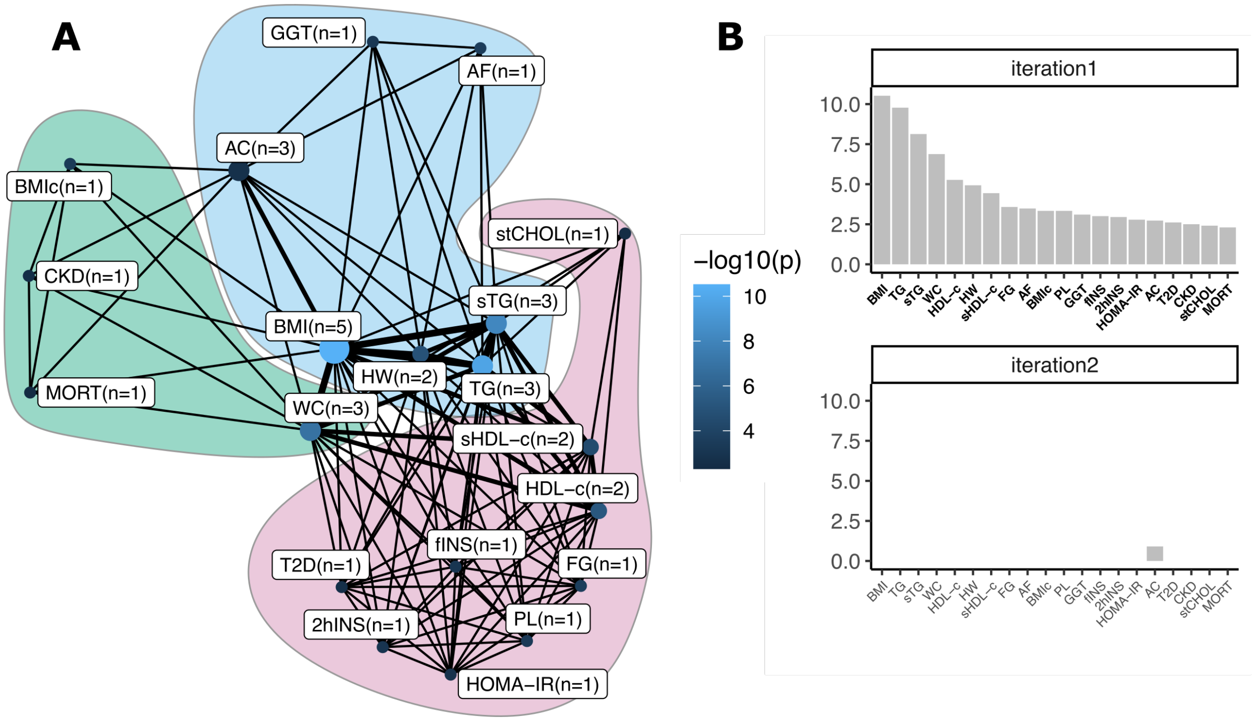


**Supplementary Figure 12.** Significant overlap between traits included in the MRC EWAS database[14] and ALS-associated sites identified using the OSCA MOA algorithm. **(A)** Network showing the traits that significantly overlap with the ALS-associated sites. Edges indicate probe overlap between the traits, with thicker lines indicating more overlapping probes.The node color indicates the enrichment -$\log_{10}(P)$, with lighter shades of blue indicating stronger associations. Colored surfaces indicate the clusters identified using the Louvain clustering algorithm. **(B)** Identification of independent clusters of traits. The first iteration shows the traits that significantly overlap with the ALS-associated probes at $FDR < 0.05$. In subsequent iterations the probes belonging to the most significant trait were excluded and enrichments tests were performed using the remaining traits. This algorithm was repeated, retaining traits that were nominally significant ($P < 0.05$, indicated in bold), until at most one trait remained significant. Abbreviations: TG = triglycerides, sTG = serum triglycerides, WC = waist circumference, HW = Hypertriglyceridemic waist, sHDL-c = serum HDL-c, FG = fasting glucose, AF = atrial fibrillation, BMIc = BMI change, PL = postprandial lipemia, GGT = Gamma-glutamyl transferase, fINS=fasting insulin, 2hINS = 2-hour insulin, HOMA-IR = Homeostatic Model Assessment of Insulin Resistance, AC = alcohol consumption per day, T2D = Type II diabetes, CKD = Chronic kidney disease, stCHOL = serum total cholesterol, MORT = Old-age mortality.

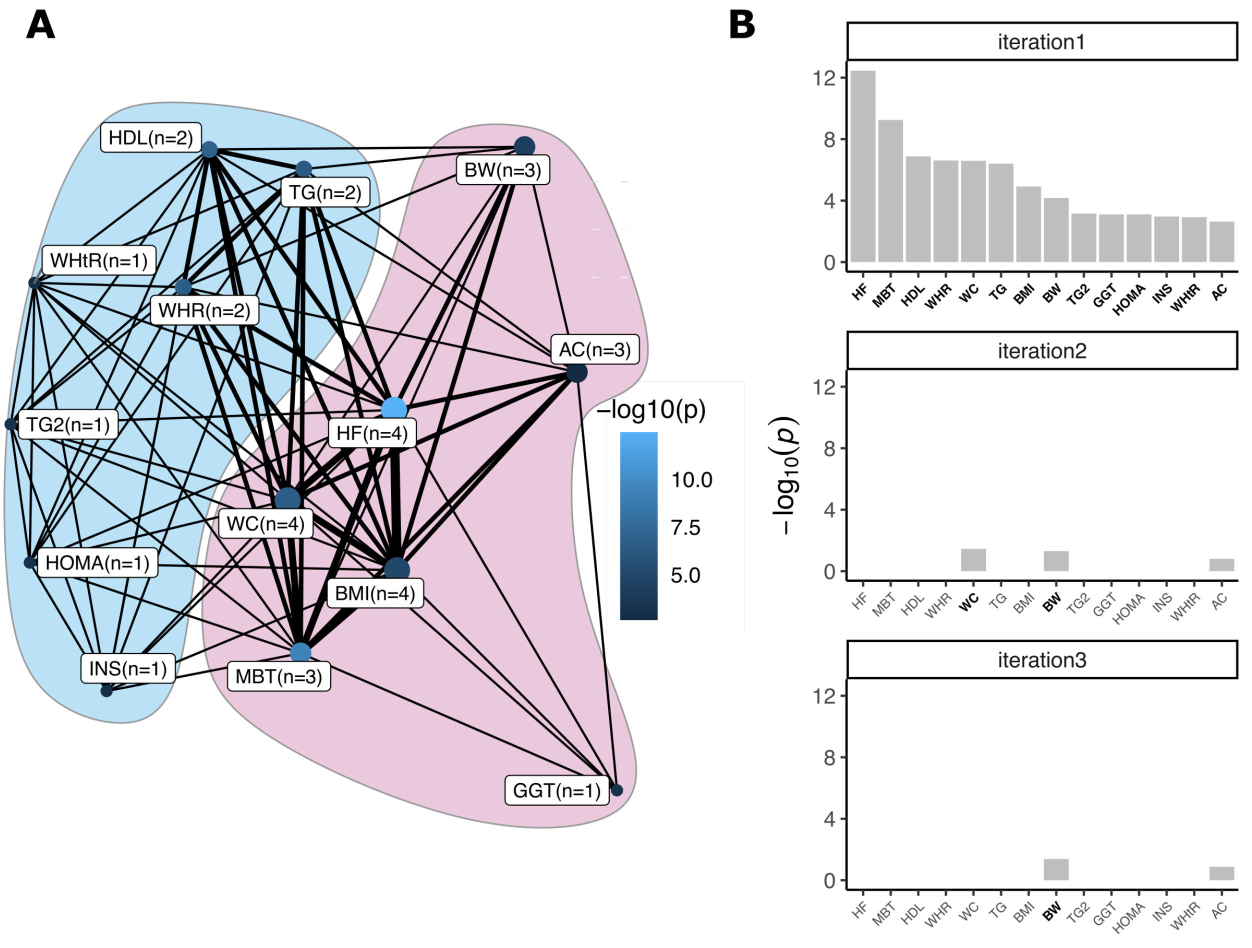


**Supplementary Figure 13.** Significant overlap between traits included in the NGDC EWAS database[13] and ALS-associated sites identified using the OSCA MOA algorithm. **(A)** Network showing the traits that significantly overlap with the ALS-associated sites. Edges indicate probe overlap between the traits, with thicker lines indicating more overlapping probes.The node color indicates the enrichment -$\log_{10}(P)$, with lighter shades of blue indicating stronger associations. Colored surfaces indicate the clusters identified using the Louvain clustering algorithm. **(B)** Identification of independent clusters of traits. The first iteration shows the traits that significantly overlap with the ALS-associated probes at $FDR < 0.05$. In subsequent iterations the probes belonging to the most significant trait were excluded and enrichments tests were performed using the remaining traits. This algorithm was repeated, retaining traits that were nominally significant ($P < 0.05$, indicated in bold), until at most one trait remained significant. Abbreviations: HF = hepatic fat, MBT = metabolic trait, WHR = waist to hip ratio, WC = waist circumference, BW = birth weight, TG2 = triglyceride postprandial responses to a high-fat dietary challenge, GGT = serum gamma-glutamyl transferase, HOMA = plasma fasting HOMA-IR levels, INS = plasma fasting insulin levels, WHtR = waist to height ratio, AC = alcohol consumption.

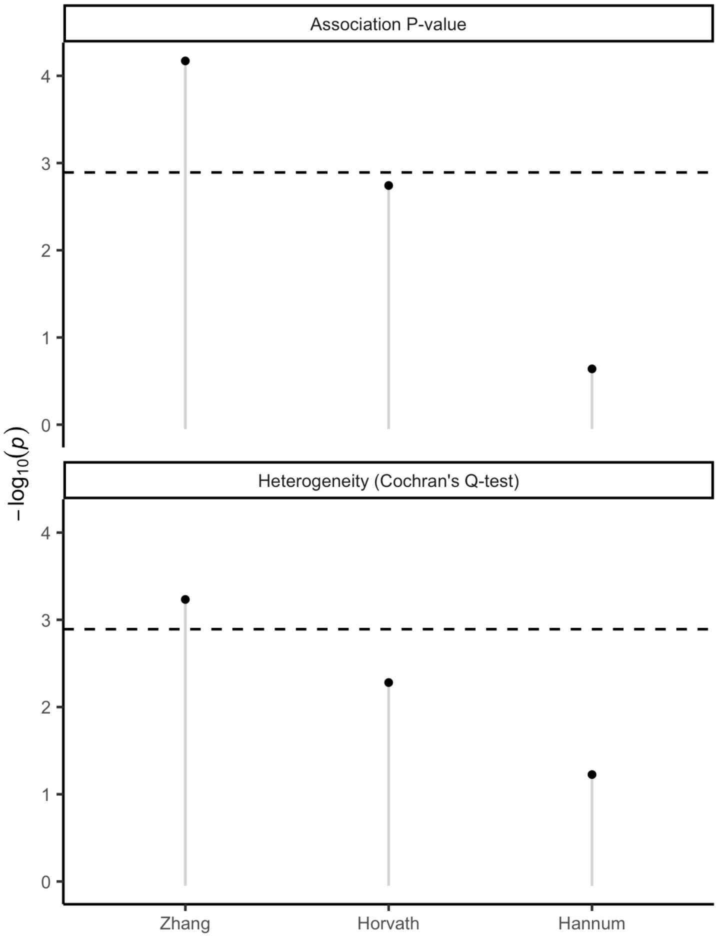


**Supplementary Figure 14.** Age acceleration test-statistics in three epigenetic clocks[15–17]. The upper panel shows the association *P*-values, the lower panel shows the heterogeneity *P*-values (Cochran's Q-test). The dashed line indicates the significance line ($1.3 \times 10^{-3}$).

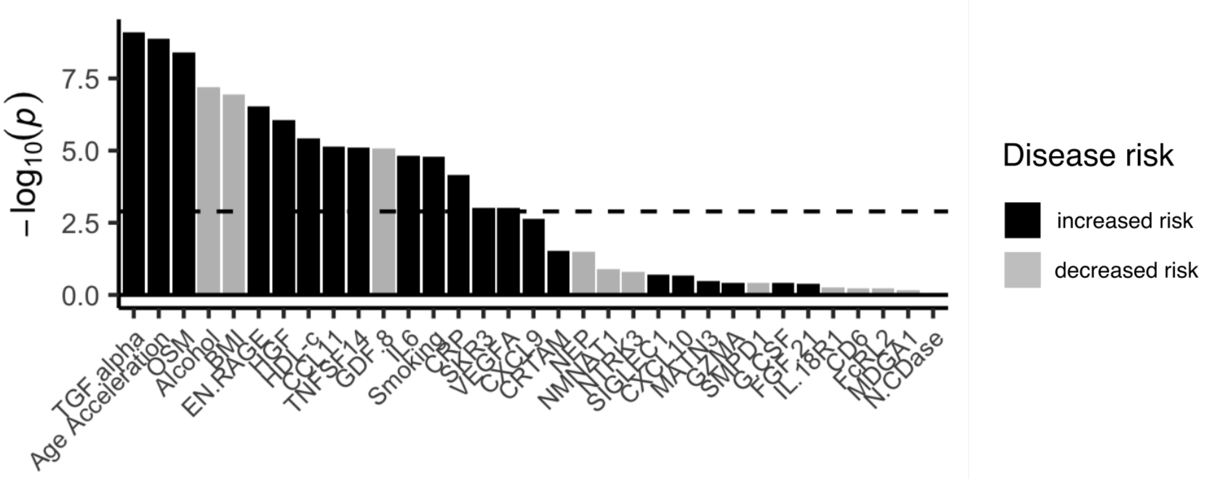


**Supplementary Figure 15.** Non-PC-adjusted PMS association *P*-values ($-\log_{10}(P)$, y-axis) for each PMS (x-axis) colored by whether a higher score is associated with increased (black) or decreased (grey) disease risk.

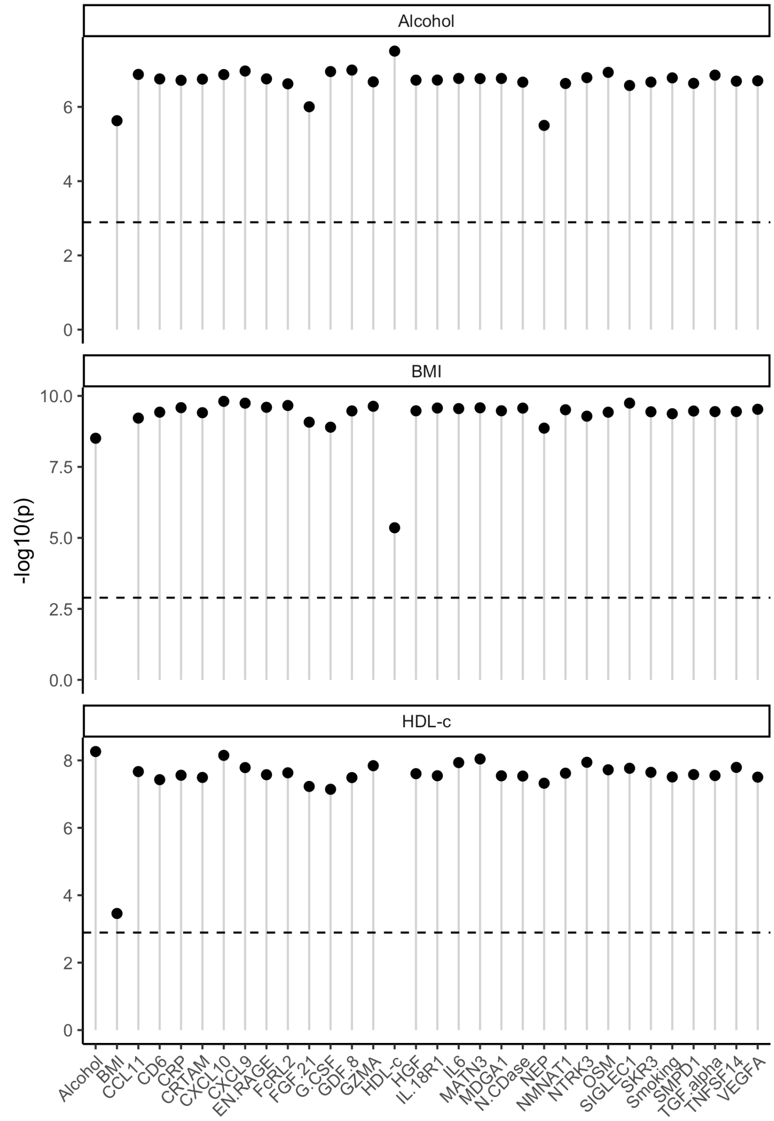


**Supplementary Figure 16.** Conditional analyses showing the -$\log_{10}$($P$) (y-axis) for Alcohol (upper), BMI (middle) and HDL-c (lower) PMS respectively, upon including the PMS indicated on the x-axis to the logistic regression model. The dashed line indicates the significance line (1.3 x $10^{-3}$).

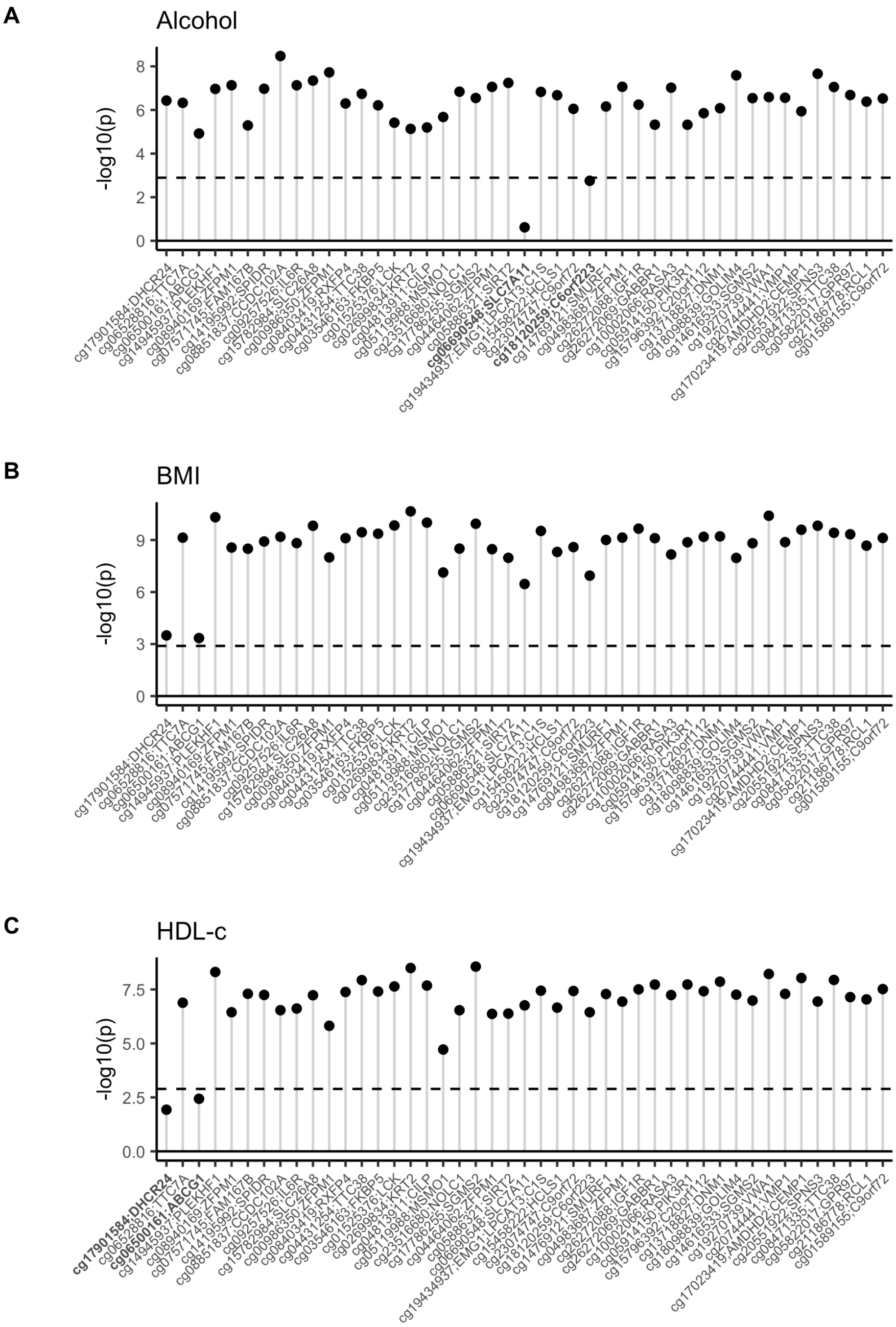


**Supplementary Figure 17.** Conditional analyses showing the -$\log_{10}(P)$ (y-axis) for the **(A)** Alcohol, **(B)** BMI and **(C)** HDL-c PMS respectively, upon including the probe indicated on the x-axis to the logistic regression model. The dashed line indicates the significance line (1.3 x $10^{-3}$).

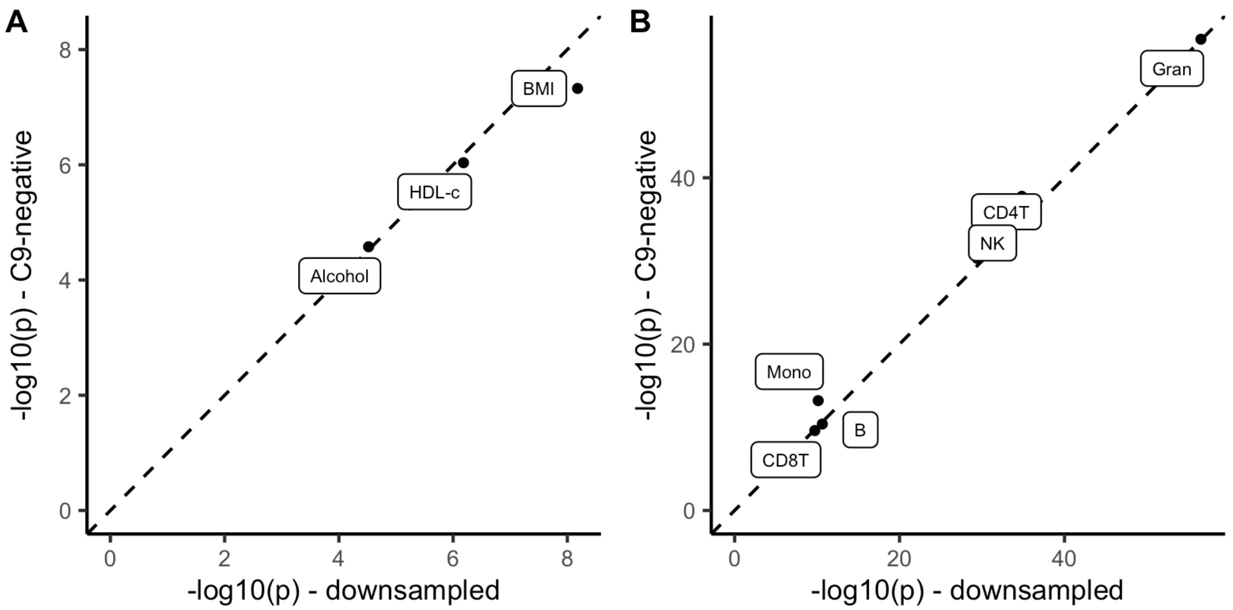


**Supplementary Figure 18.** Sensitivity analyses comparing PMS associations where we excluded individuals carrying the C9 repeat expansion (y-axis), compared to PMS associations including C9-carriers where we randomly downsampled the number of samples to match the sample size of the C9-negative sample set (x-axis).

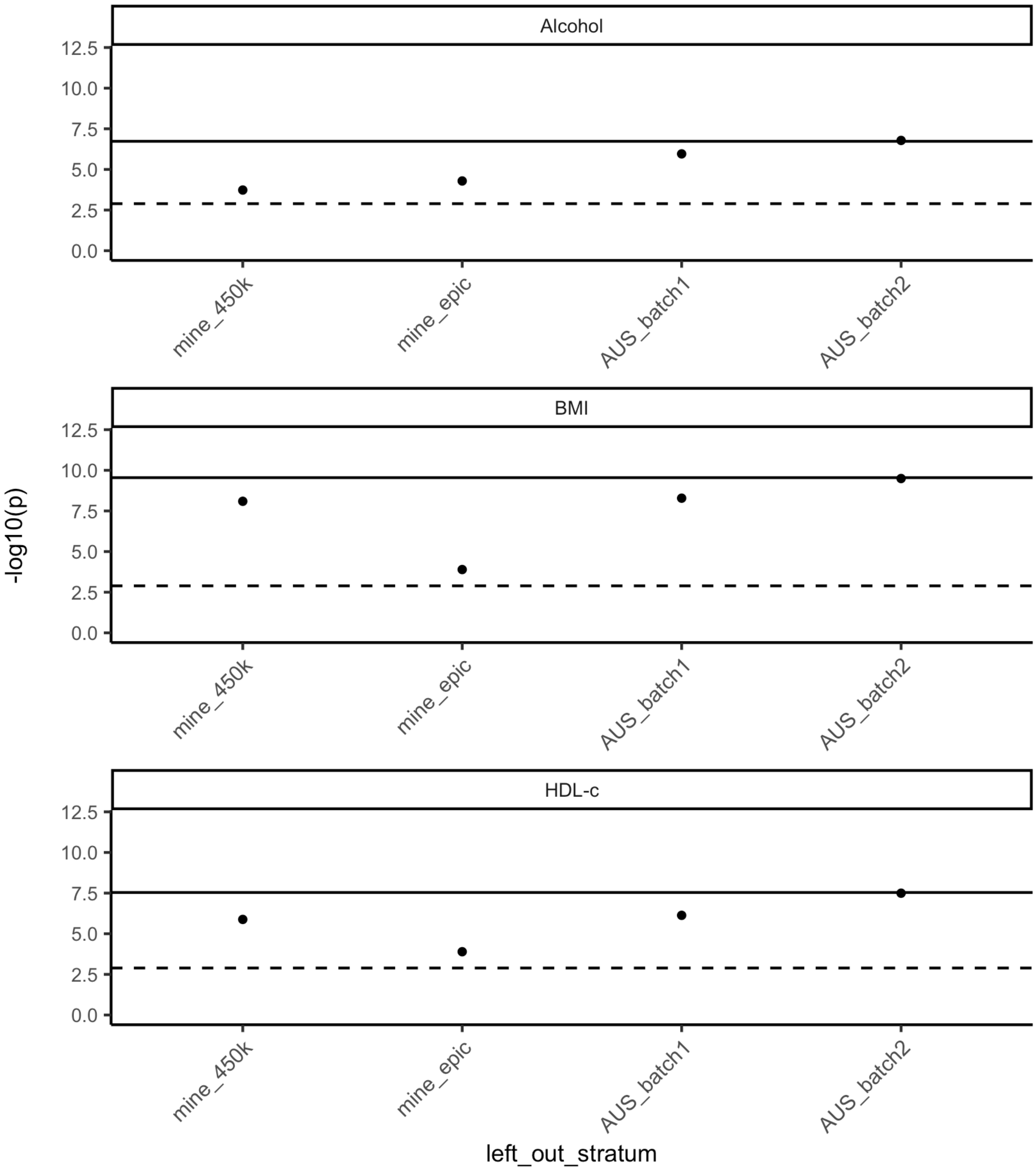


**Supplementary Figure 19.** PMS association *P*-values (-$\log_{10}$(*P*); y-axis) upon excluding strata from the meta-analysis (x-axis).The dashed line indicates the significance line ($1.3 \times 10^{-3}$), the solid line indicates the *P*-value including all strata for the respective PMS.

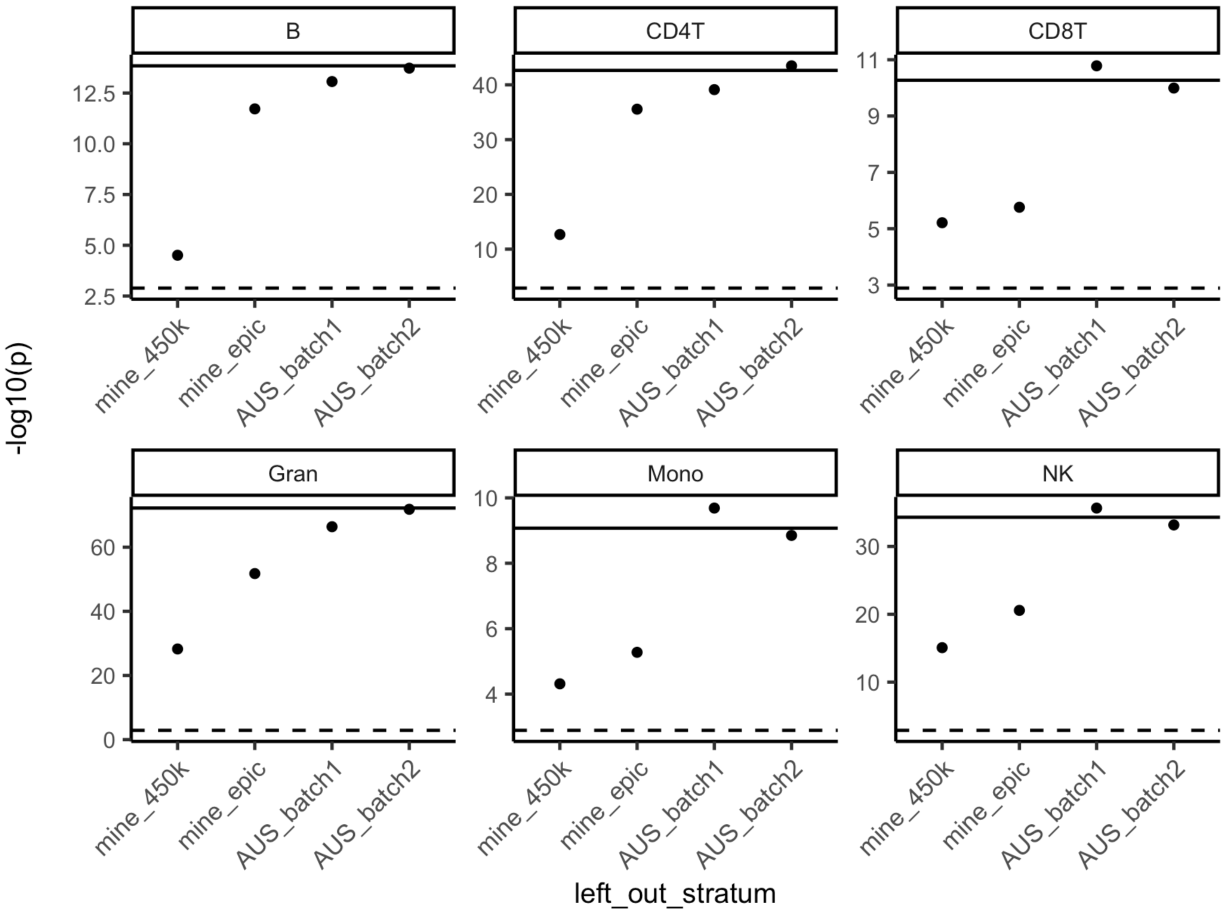


**Supplementary Figure 20.** PMS association *P*-values (-$\log_{10}$(*P*); y-axis) upon excluding strata from the meta-analysis (x-axis).The dashed line indicates the significance line (1.3 x $10^{-3}$), the solid line indicates the *P*-value including all strata for the respective PMS.

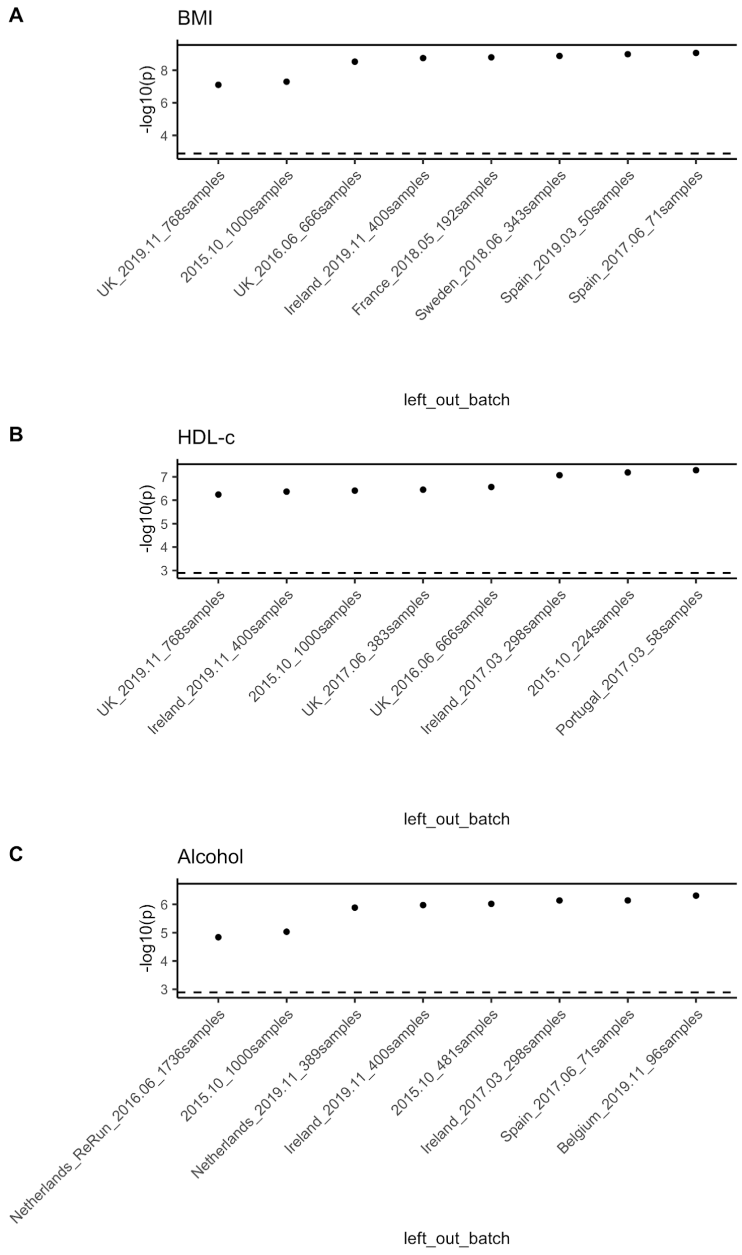


**Supplementary Figure 21.** PMS association *P*-values ($-\log_{10}(P)$; y-axis) upon excluding experimental batches (x-axis). The dashed line indicates the significance line ($1.3 \times 10^{-3}$), the solid line indicates the *P*-value including all cohorts for the respective PMS. The eight experimental batches resulting in the least significant *P*-value upon exclusion are shown.

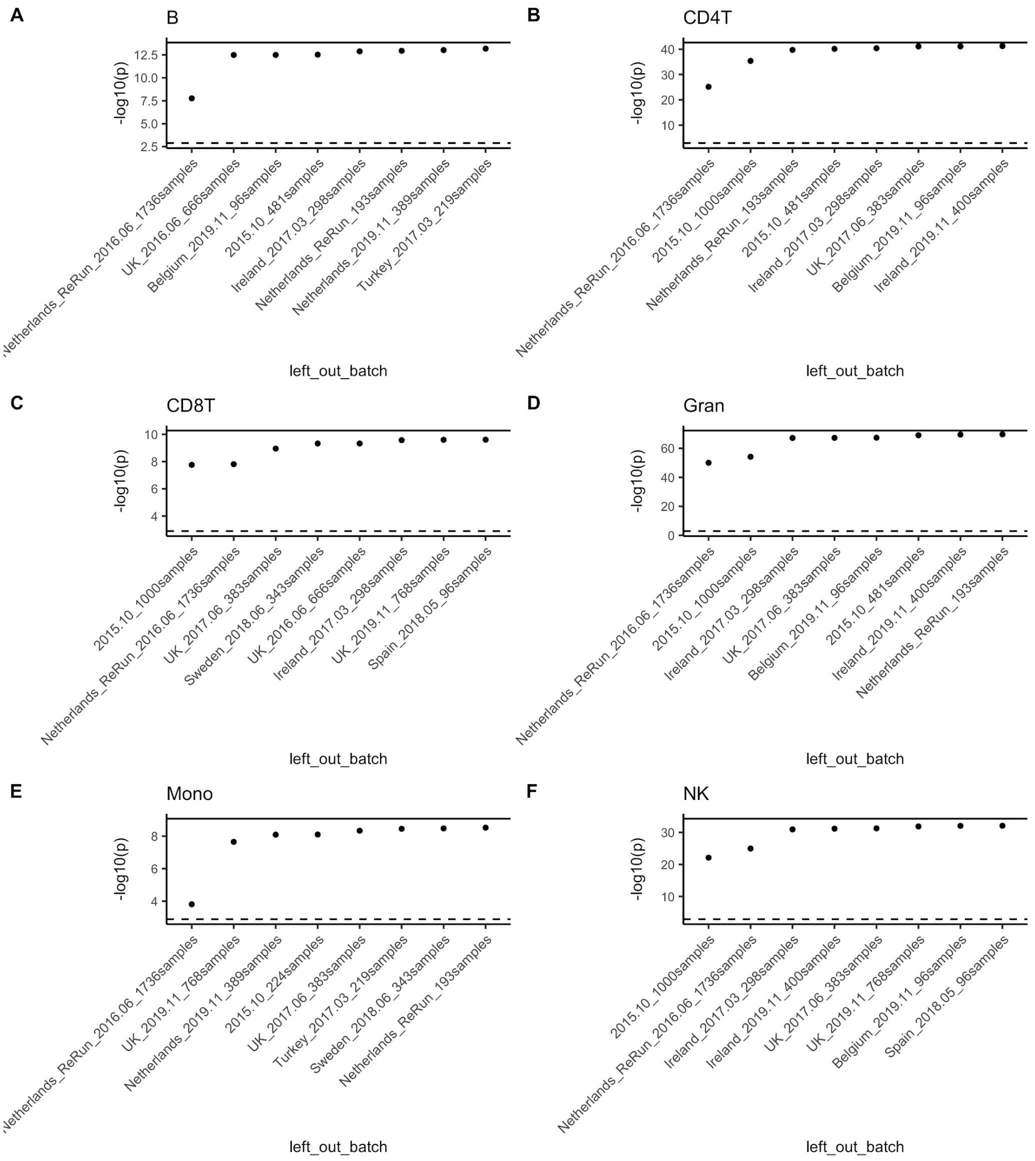


**Supplementary Figure 22.** PMS association *P*-values (-$\log_{10}(P)$; y-axis) upon excluding experimental batches (x-axis).The dashed line indicates the significance line (1.3 x $10^{-3}$), the solid line indicates the *P*-value including all strata for the respective PMS.
The eight experimental batches resulting in the least significant *P*-value upon exclusion are shown.

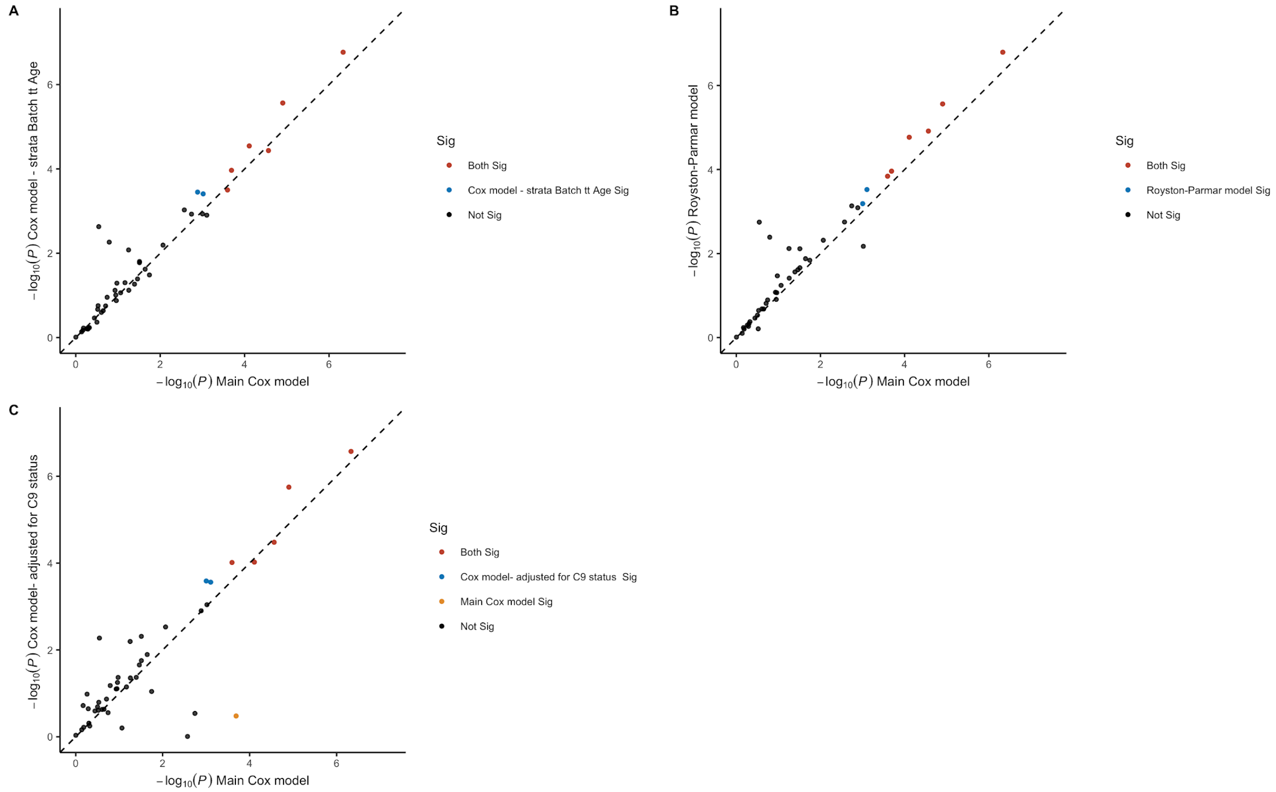


**Supplementary Figure 23.** Association *P*-values (-$\log_{10}$(*P*)) of the main Cox model (x-axis) for all 45 sites to *P*-values from a Cox model that was stratified on experimental batch, and predicted age was time-transformed as described in the Methods section (y-axis). **(B)** Association *P*-values (-$\log_{10}$(*P*)) of the main Cox model (x-axis) compared to *P*-values from a Royston-Parmar spline model with one knot. **(C)** Association *P*-values (-$\log_{10}$(*P*)) of the main Cox model (x-axis) for 45 sites compared to *P*-values from a Cox model where C9 status (normal or expanded) was added as additional covariate (y-axis).

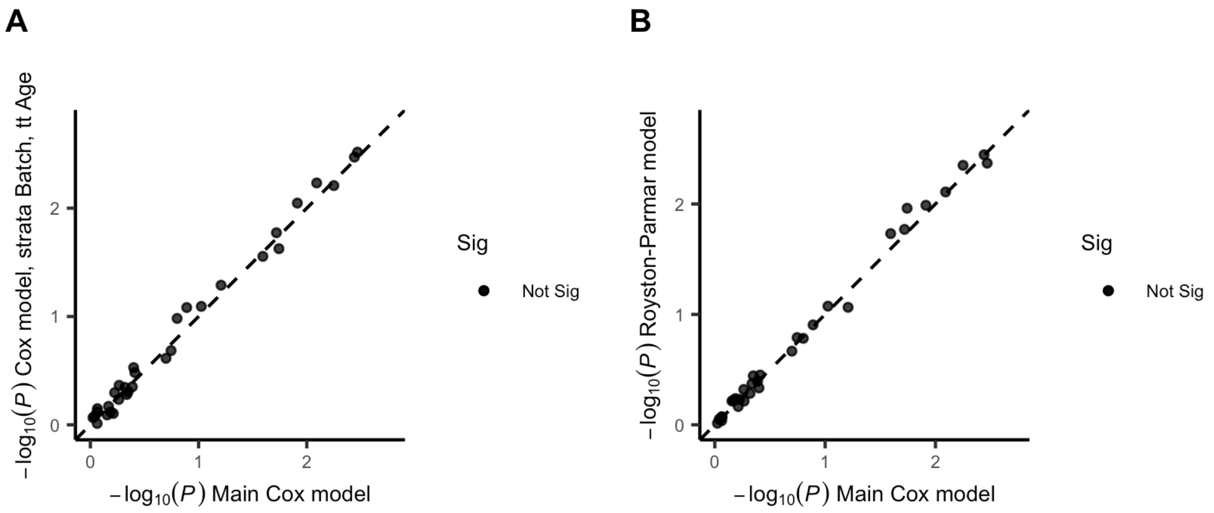


**Supplementary Figure 24. (A)** Association *P*-values (-$\log_{10}$(*P*)) of the main Cox model (x-axis) for all PMSs except WBC compared to *P*-values from a Cox model that was stratified on experimental batch, and where predicted age was time-transformed as described in the Methods section (y-axis). **(B)** Association *P*-values (-$\log_{10}$(*P*)) of the main Cox model (x-axis) compared to *P*-values from a Royston-Parmar spline model with one knot.

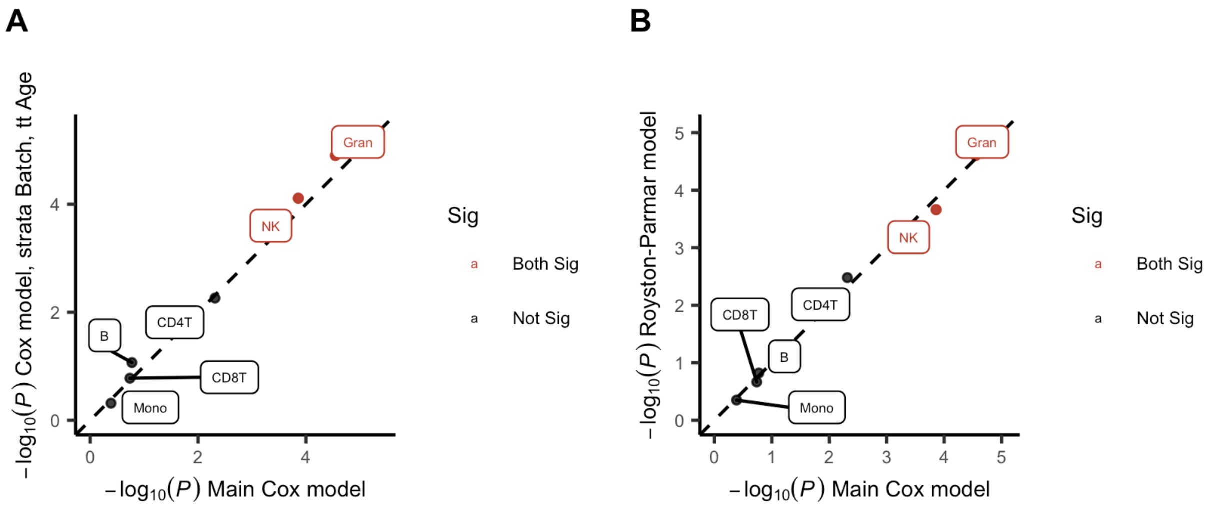


**Supplementary Figure 25.** Association *P*-values (-$\log_{10}$(*P*)) of the main Cox model (x-axis) for the WBC PMSs compared to *P*-values from a Cox model that was stratified on experimental batch, and where predicted age was time-transformed as described in the Methods section (y-axis). **(B)** Association *P*-values (-$\log_{10}$(*P*)) of the main Cox model (x-axis) compared to *P*-values from a Royston-Parmar spline model with one knot.

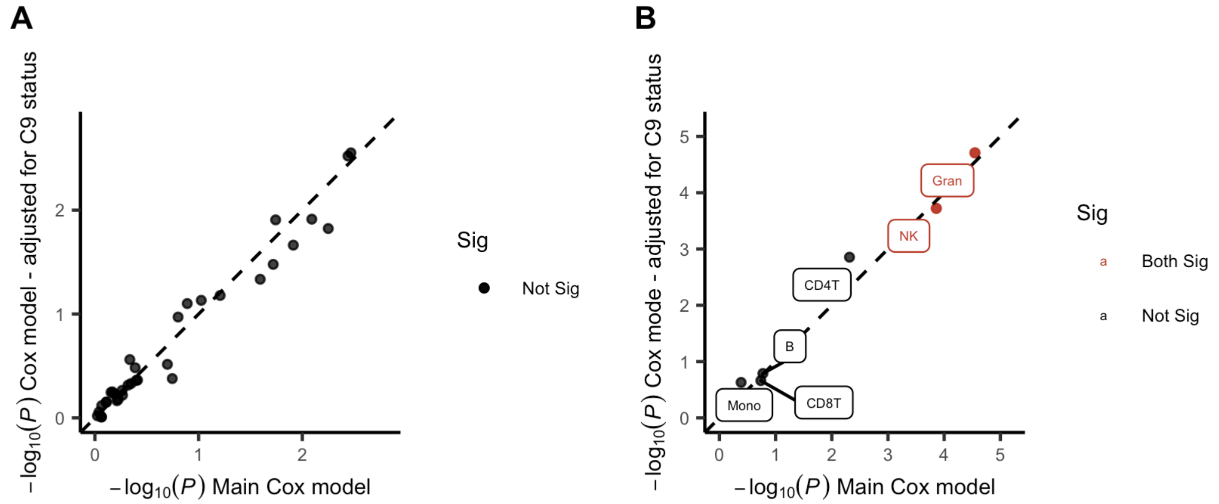


**Supplementary Figure 26.** Association *P*-values (-$\log_{10}$(*P*)) of the main Cox model (x-axis) for all PMSs except WBC compared to *P*-values from a Cox model where C9 status (normal or expanded) was added as additional covariate (y-axis). **(B)** Association *P*-values (-$\log_{10}$(*P*)) of the main Cox model (x-axis) for the WBC PMSs compared to *P*-values from a Cox model where C9 status (normal or expanded) was added as additional covariate (y-axis).

**A**

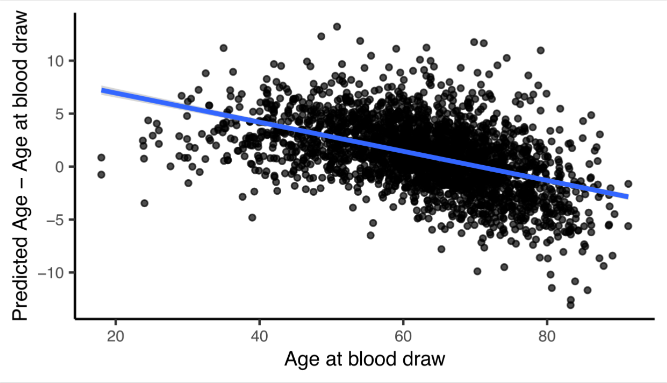


**B**

| Outcome | Predictor | Adjusted for Age at blood draw | b | se | p |
|---|---|---|---|---|---|
| Age at Onset | Predicted Age – Age at blood draw | no | -1.22 | 0.028 | 0 |
| Age at Onset | Predicted Age – Age at blood draw | yes | 0.014 | 0.012 | 0.26 |

**C**

| Outcome | Predictor | Adjusted for Age at blood draw | b | se | HR | p |
|---|---|---|---|---|---|---|
| Survival | Predicted Age – Age at blood draw | no | -0.016 | 0.0056 | 0.98 | 0.0038 |
| Survival | Predicted Age – Age at blood draw | yes | 0.0019 | 0.0067 | 1.0 | 0.78 |

**Supplementary Figure 27. (A)** Association between age at blood draw, and age acceleration defined as: Predicted Age – Age at blood draw. There is a strong negative correlation (b = -1.49, $P = 2.9 \times 10^{-223}$) between Age at blood draw and age acceleration defined in this way[18]. **(B)** Association between Age at Onset and Age acceleration defined as: Predicted Age – Age at blood draw, the same covariates as in the LB EWAS were included. There is a strong association without age adjustment, this association disappears when adjusting for age at blood draw. **(C)** Association between survival (Cox proportional hazards model) and Age acceleration defined as: Predicted Age – Age at blood draw, the same covariates as in the LB EWAS were included. There is a significant association without age adjustment, this association disappears when adjusting for age at blood draw.
