## Supplemental file 1 for "Genome-wide study of DNA methylation in Amyotrophic Lateral Sclerosis identifies differentially methylated loci and implicates metabolic, inflammatory and cholesterol pathways"

### Quality Control

March 6, 2021

#### Contents

**1 MinE 450k** **2**

**2 MinE EPIC** **9**

**3 AUS1** **16**

**4 AUS2** **23**

**5 Definition of AUS1 and AUS2 strata** **29**

### 1 MinE 450k

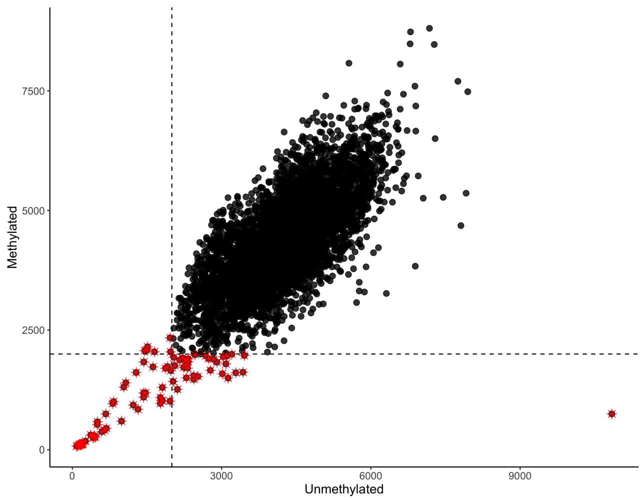


**QC figure 1:** Median unmethylated vs. median methylated signal.

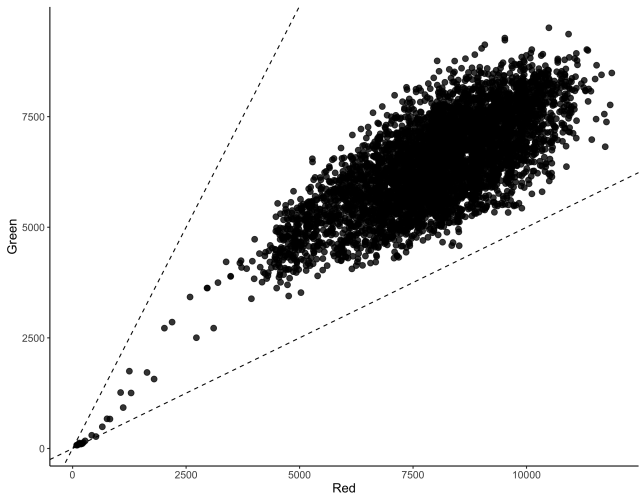


**QC figure 2:** Median red vs. median green signal, calculated in type I probes.

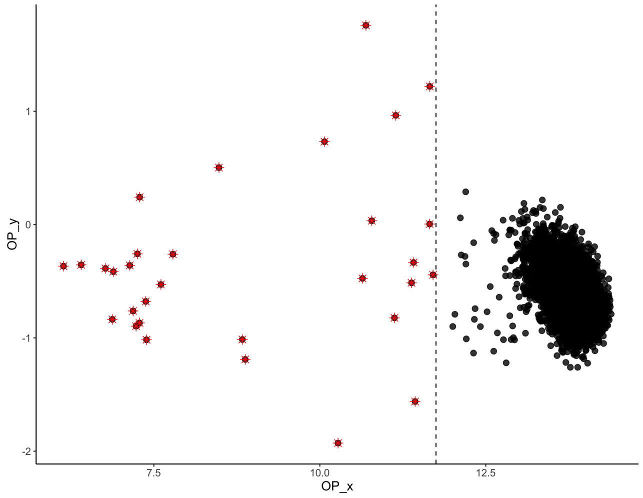


**QC figure 3:** Rotated OP (Overall quality) plot based on non-polymorphic control probes as implemented in the *MethylAid* package [1].

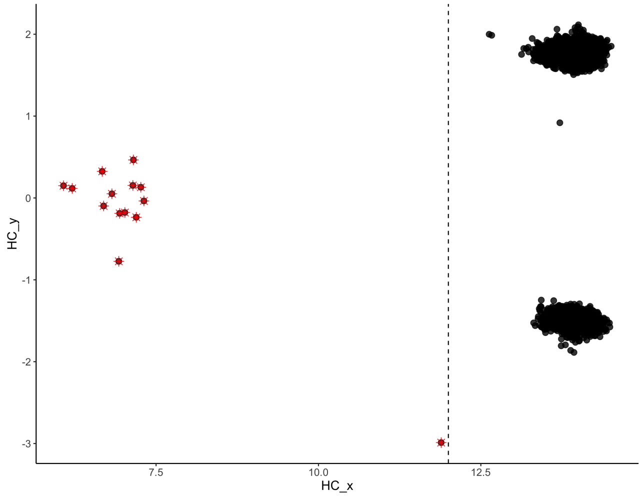


**QC figure 4:** Rotated HC plot based on hybridization control probes as implemented in the *MethylAid* package [1].

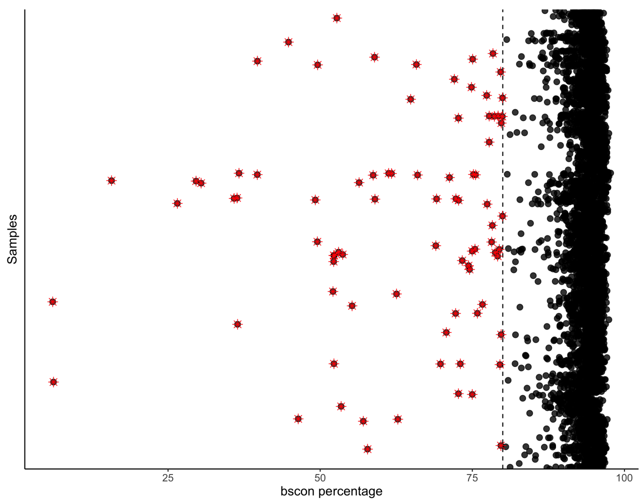


**QC figure 5:** bscon metric from the *wateRmelon* package, which measures bisulfite conversion efficiency [2].

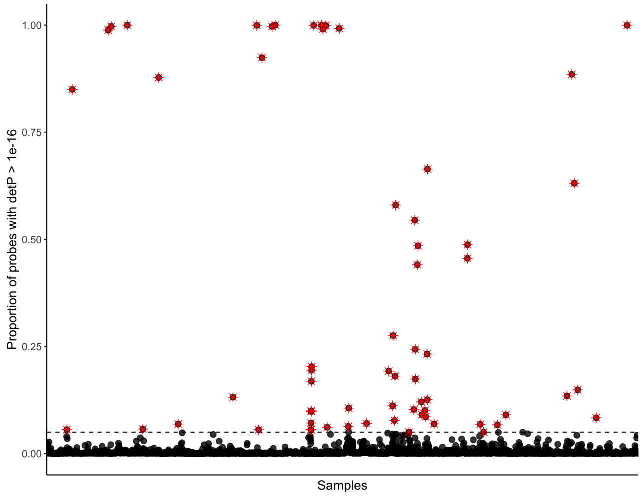


**QC figure 6:** Percentage of probes with detection P-value $> 1 \times 10^{-16}$

.

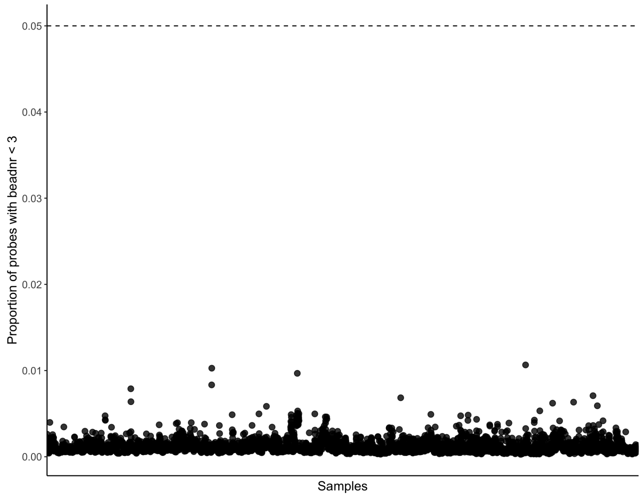


**QC figure 7:** Percentage of probes with beadNr $< 3$.

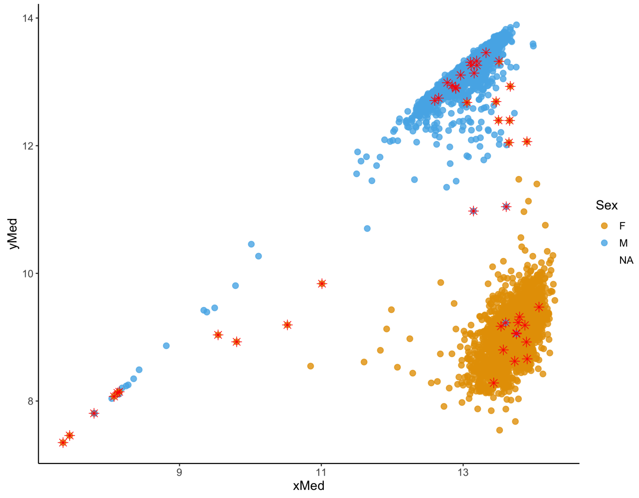


**QC figure 8:** Median X and Y copy number, as implemented in the *minfi* package [3].

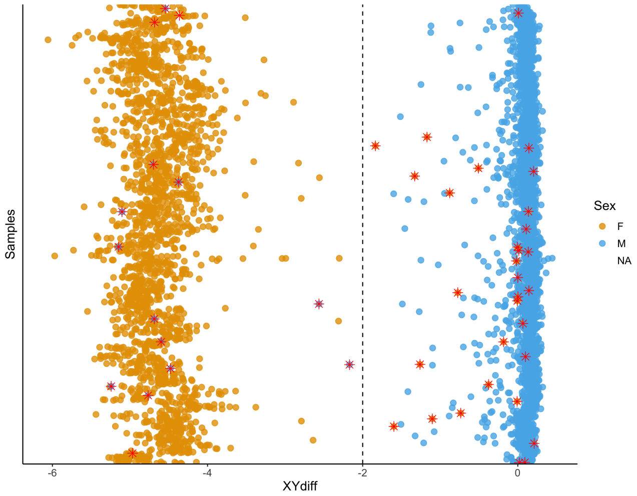


**QC figure 9:** Difference between median Y-chromosomal and median X-chromosomal signal intensity.

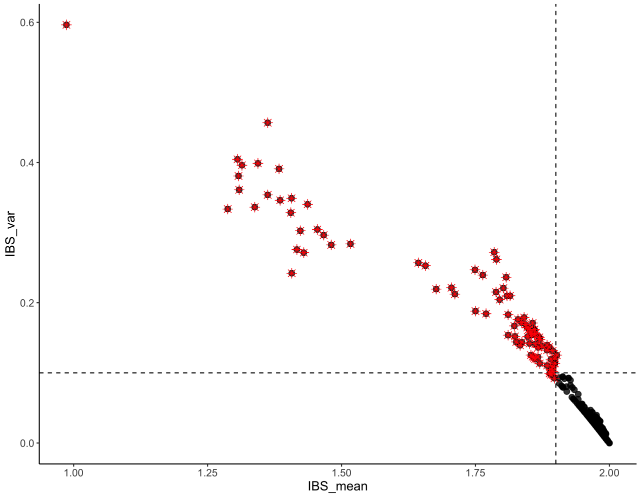


**QC figure 10:** Identity-by-descent (IBS) comparing DNAm-inferred SNPs and SNPs measured using whole-genome sequencing data.

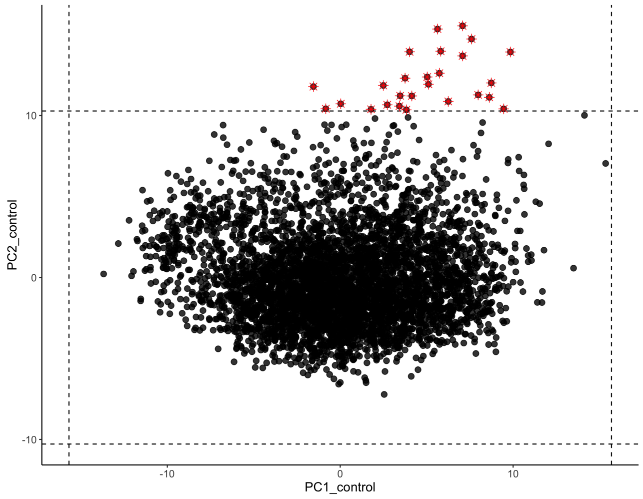


**QC figure 11:** Control probe principal components, after first round of QC.

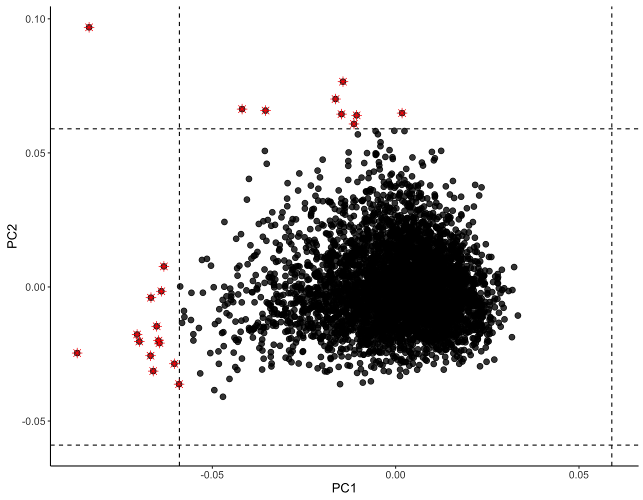


**QC figure 12:** Array-wide principal components, after first round of QC.

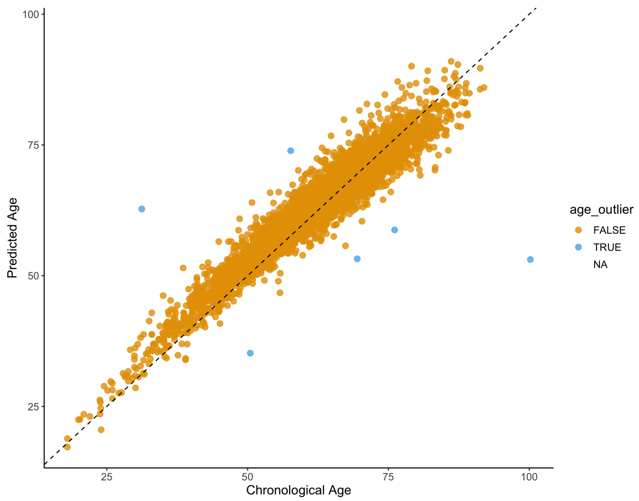


**QC figure 13:** Chronological age vs. Predicted Age based on the Zhang *et al.* age predictor [4].

#### 2 MinE EPIC

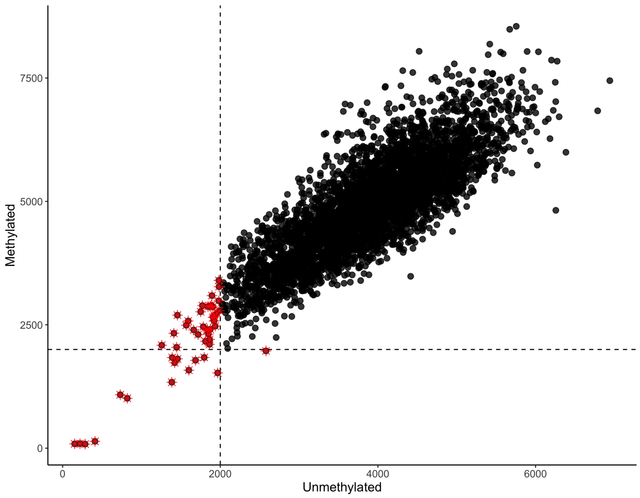


**QC figure 14:** Median unmethylated vs. median methylated signal.

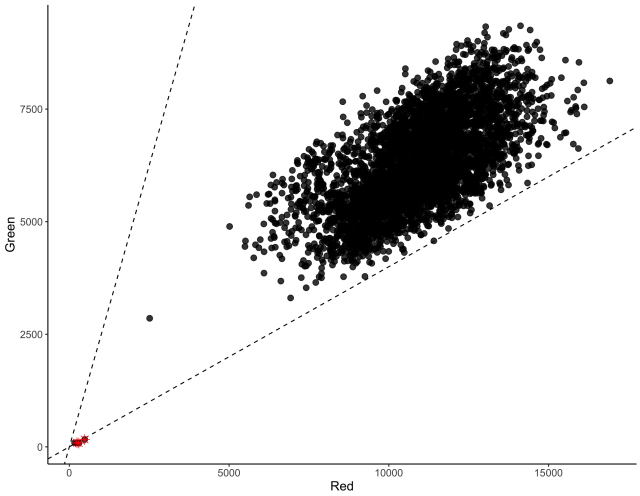


**QC figure 15:** Median red vs. median green signal, calculated in type I probes.

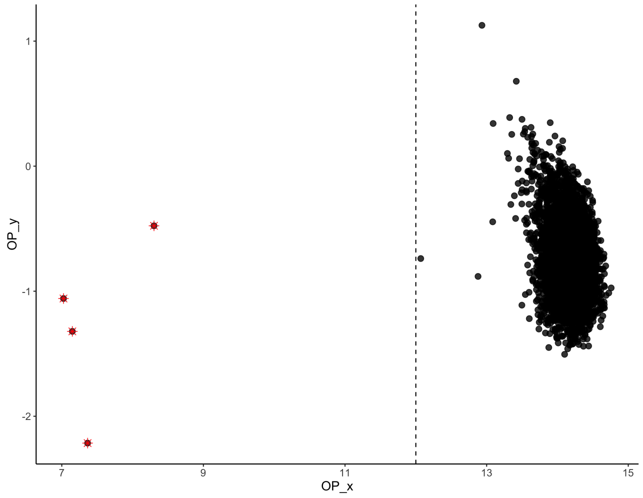


**QC figure 16:** Rotated OP (Overall quality) plot based on non-polymorphic control probes as implemented in the *MethylAid* package [1].

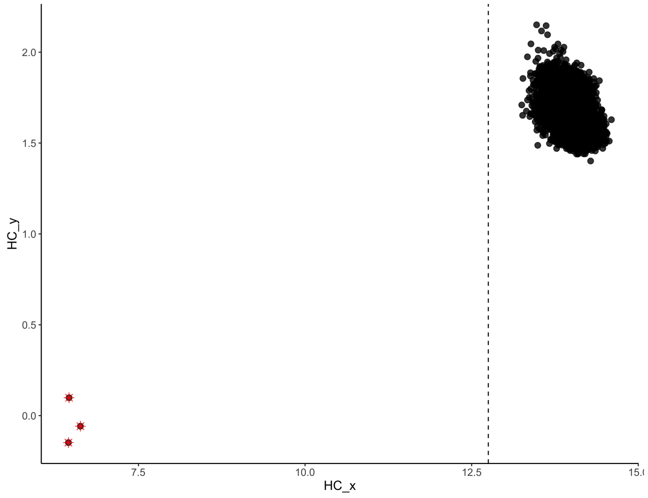


**QC figure 17:** Rotated HC plot based on hybridization control probes as implemented in the *MethylAid* package [1].

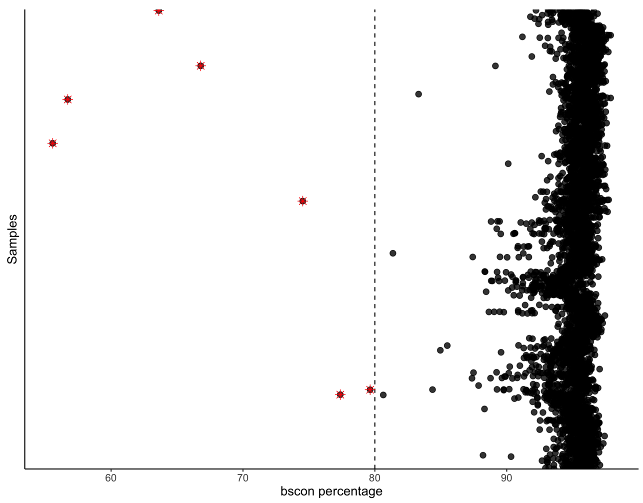


**QC figure 18:** bscon metric from the *wateRmelon* package, which measures bisulfite conversion efficiency [2].

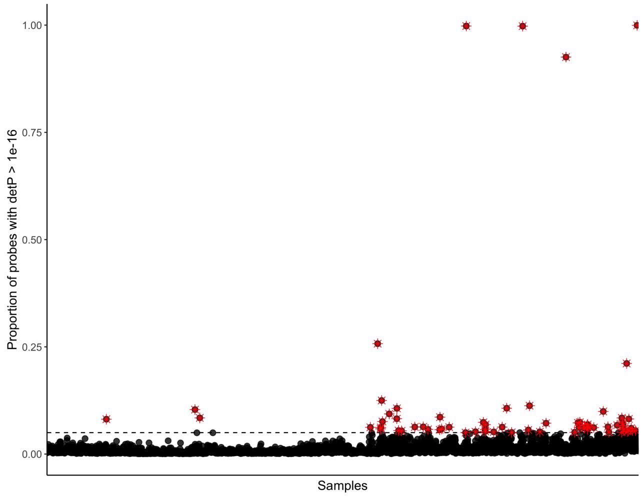


**QC figure 19:** Percentage of probes with detection P-value $> 1 \times 10^{-16}$

.

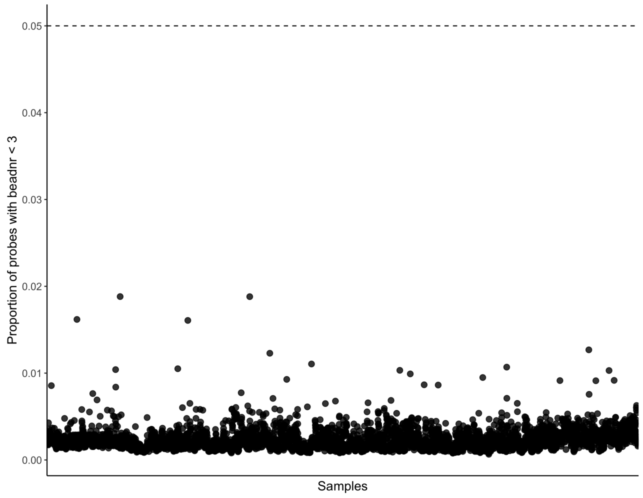


**QC figure 20:** Percentage of probes with beadNr $<3$.

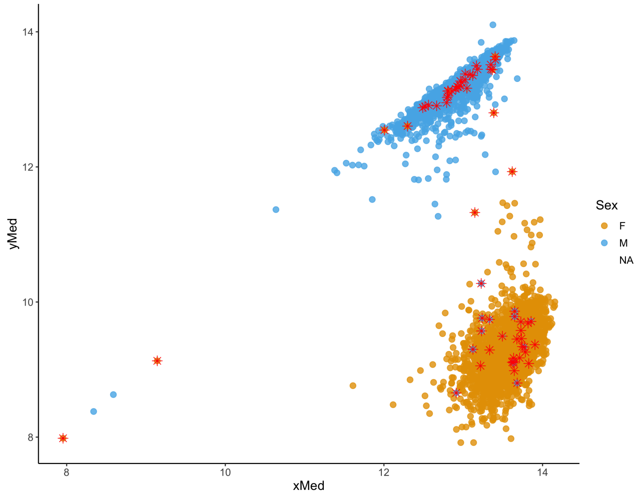


**QC figure 21:** Median X and Y copy number, as implemented in the *minfi* package [3].

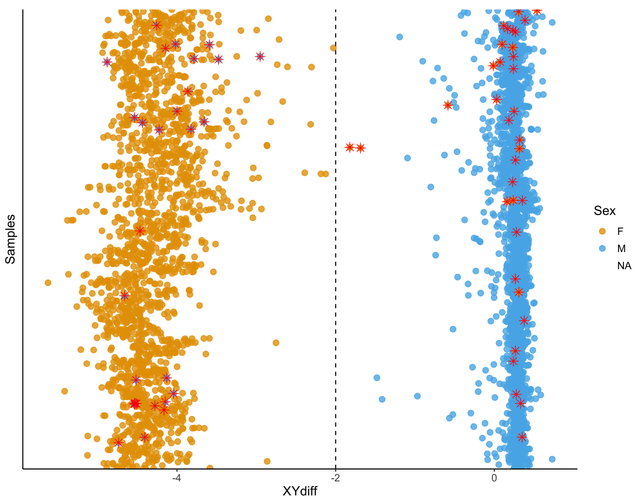


**QC figure 22:** Difference between median Y-chromosomal and median X-chromosomal signal intensity.

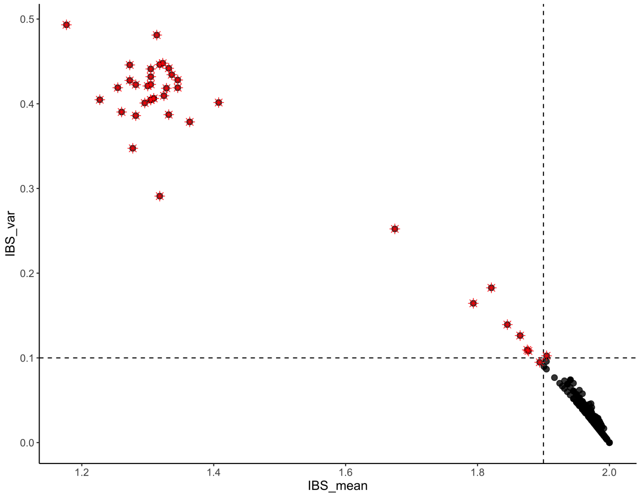


**QC figure 23:** Identity-by-descent (IBS) comparing DNAm-inferred SNPs and SNPs measured using whole-genome sequencing data.

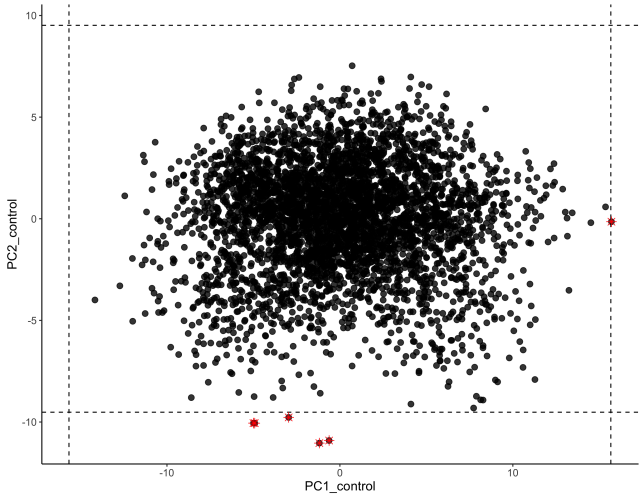


**QC figure 24:** Control probe principal components, after first round of QC.

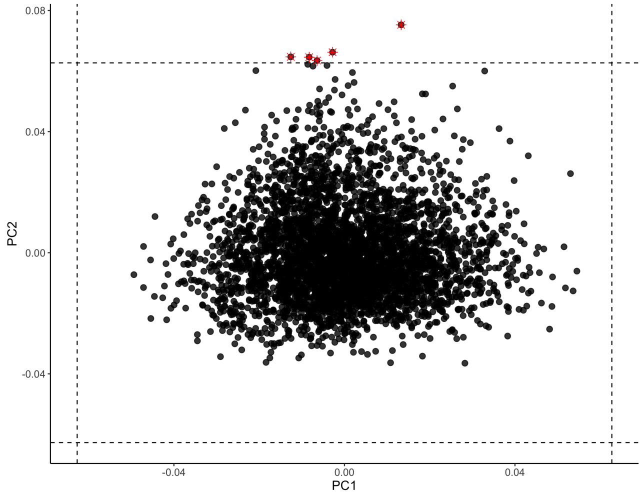


**QC figure 25:** Array-wide principal components, after first round of QC.

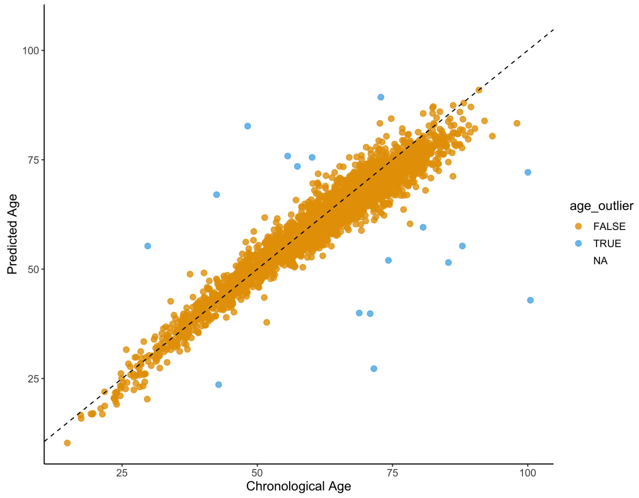


**QC figure 26:** Chronological age vs. Predicted Age based on the Zhang *et al.* age predictor [4].

### 3 AUS1

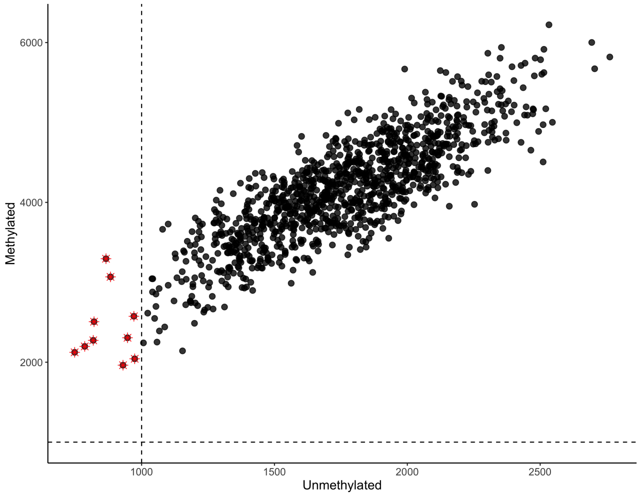


**QC figure 27:** Median unmethylated vs. median methylated signal.

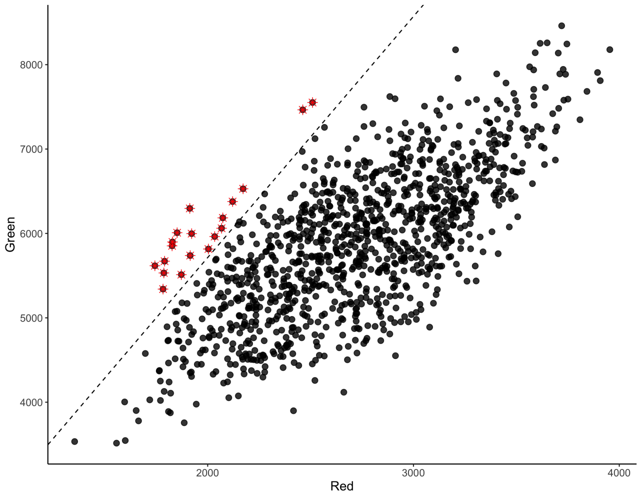


**QC figure 28:** Median red vs. median green signal, calculated in type I probes.

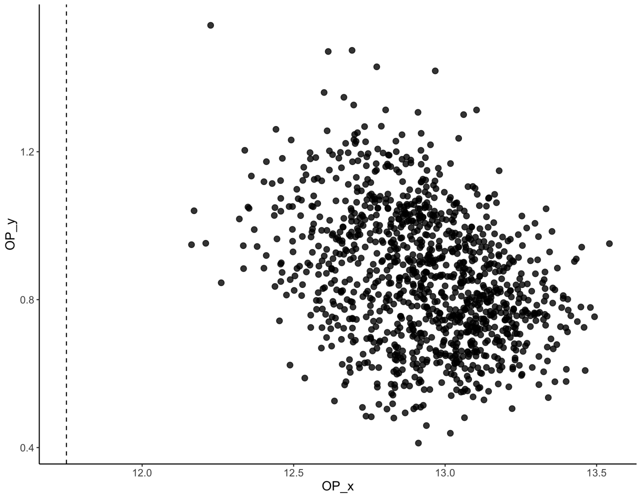


**QC figure 29:** Rotated OP (Overall quality) plot based on non-polymorphic control probes as implemented in the *MethylAid* package [1].

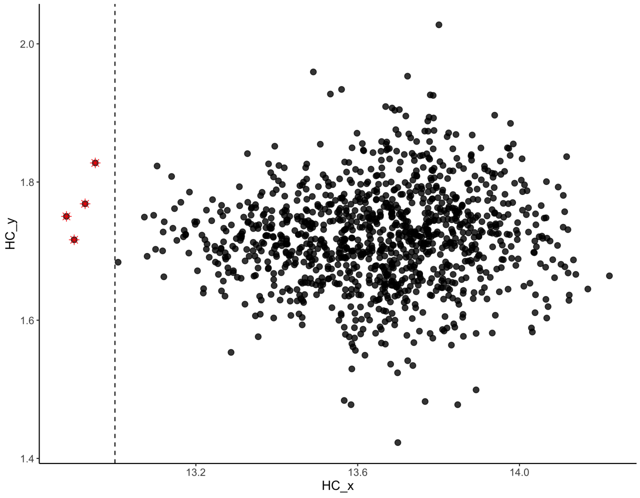


**QC figure 30:** Rotated HC plot based on hybridization control probes as implemented in the *MethylAid* package [1].

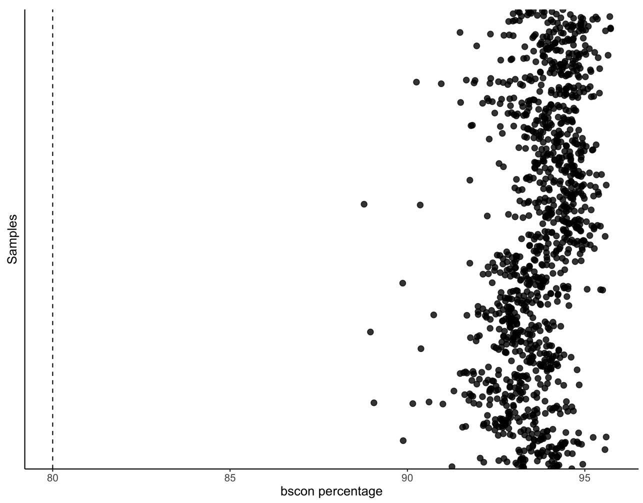


**QC figure 31:** bscon metric from the *wateRmelon* package, which measures bisulfite conversion efficiency [2].

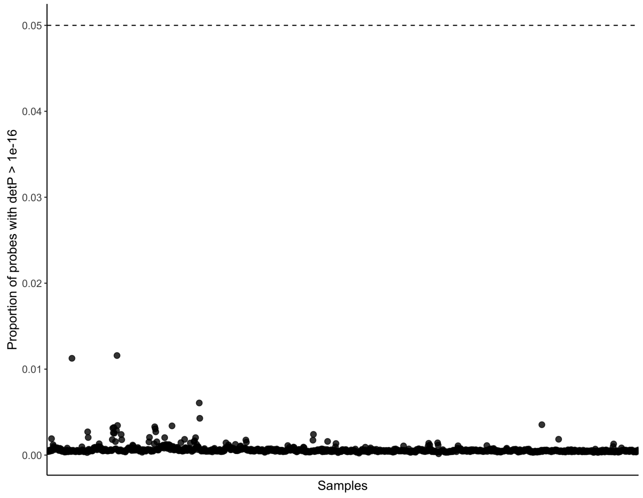


**QC figure 32:** Percentage of probes with detection P-value $> 1 \times 10^{-16}$

.

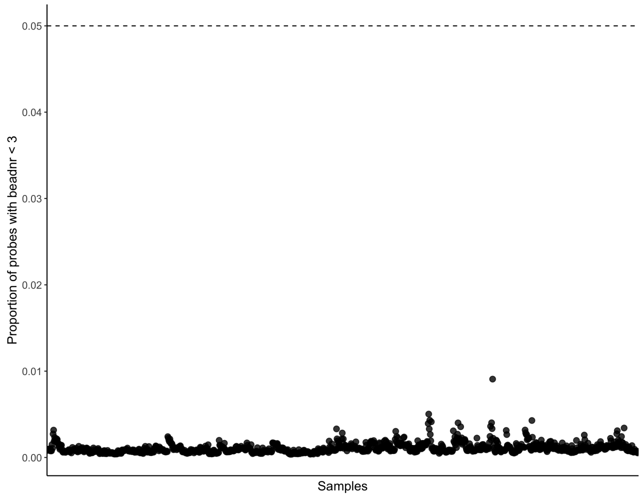


**QC figure 33:** Percentage of probes with beadNr $< 3$.

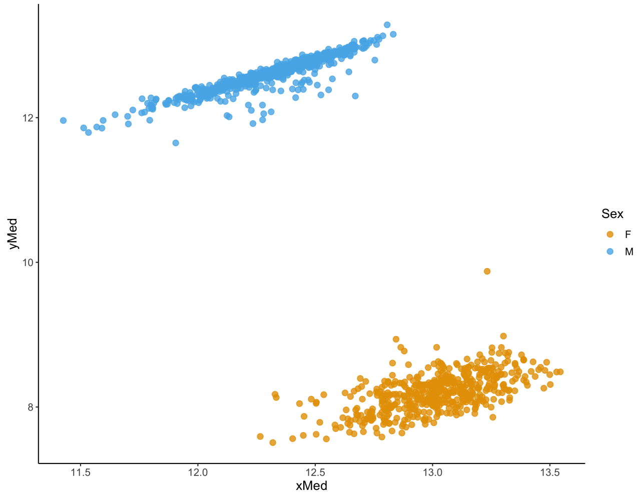


**QC figure 34:** Median X and Y copy number, as implemented in the *minfi* package [3].

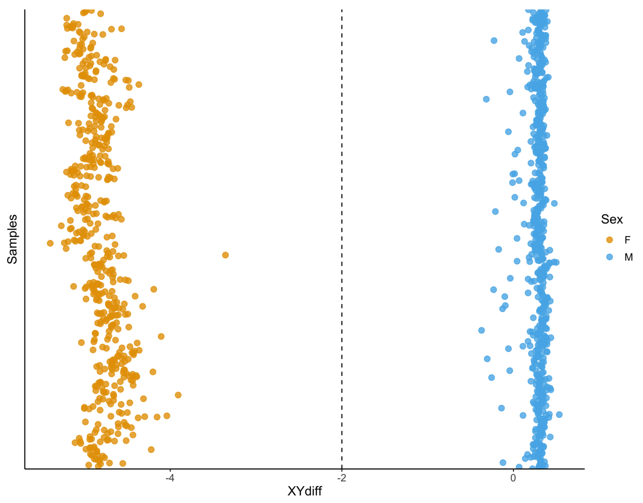


**QC figure 35:** Difference between median Y-chromosomal and median X-chromosomal signal intensity.

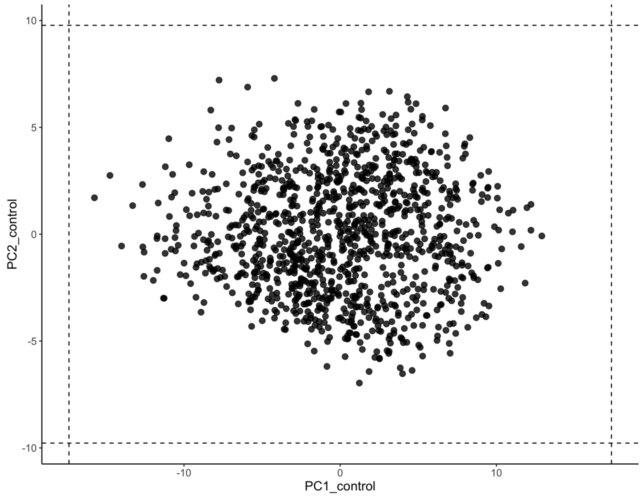


**QC figure 36:** Control probe principal components, after first round of QC.

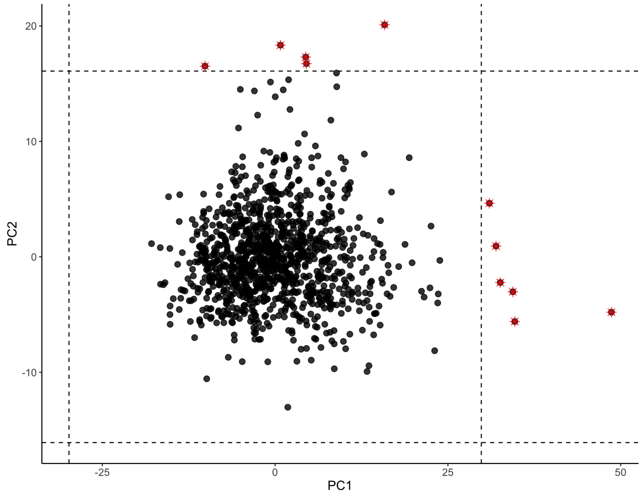


**QC figure 37:** Array-wide principal components, after first round of QC.

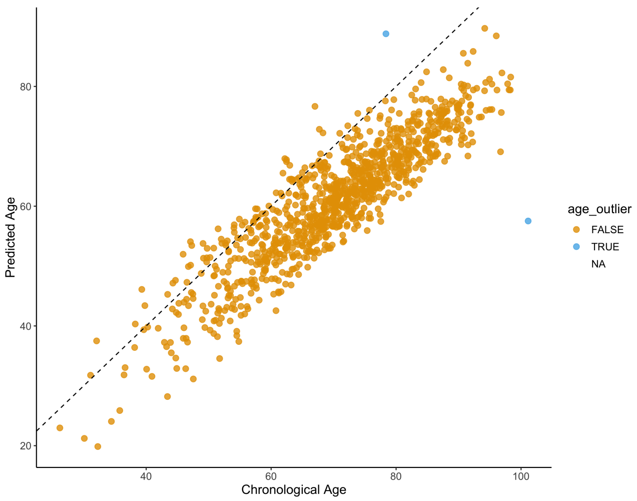


**QC figure 38:** Chronological age vs. Predicted Age based on the Zhang *et al.* age predictor [4].

### 4 AUS2

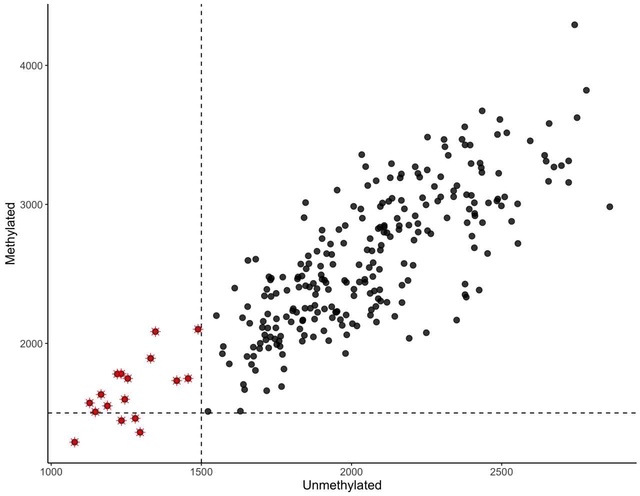


**QC figure 39:** Median unmethylated vs. median methylated signal.

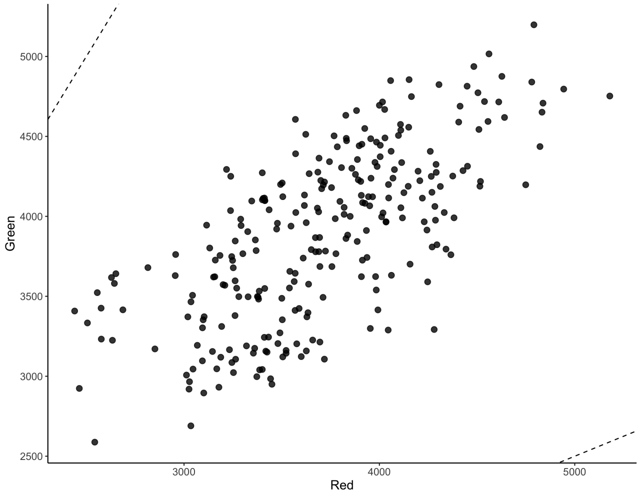


**QC figure 40:** Median red vs. median green signal, calculated in type I probes.

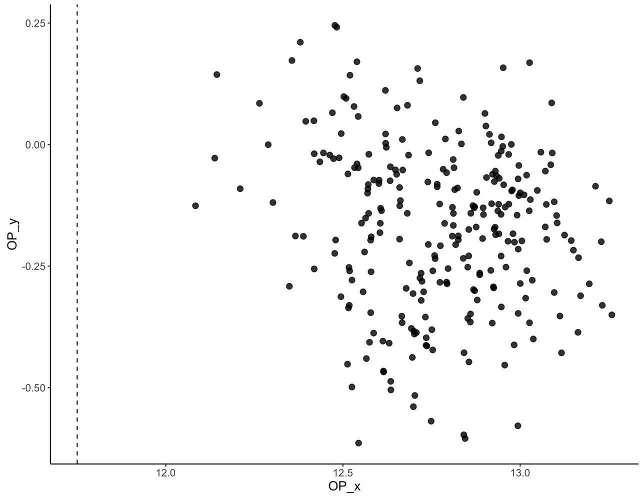


**QC figure 41:** Rotated OP (Overall quality) plot based on non-polymorphic control probes as implemented in the *MethylAid* package [1].

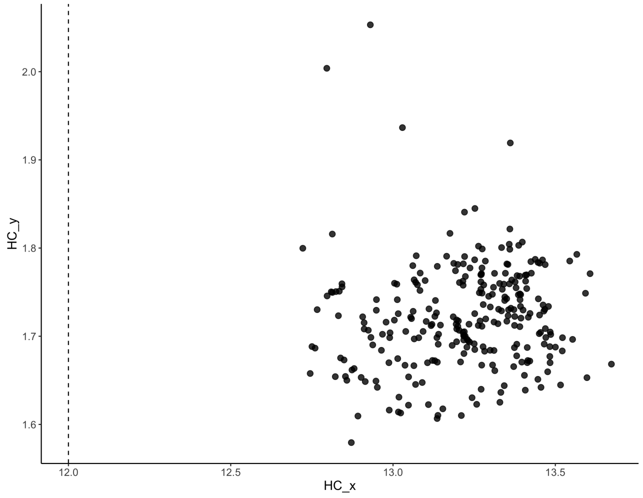


**QC figure 42:** Rotated HC plot based on hybridization control probes as implemented in the *MethylAid* package [1].

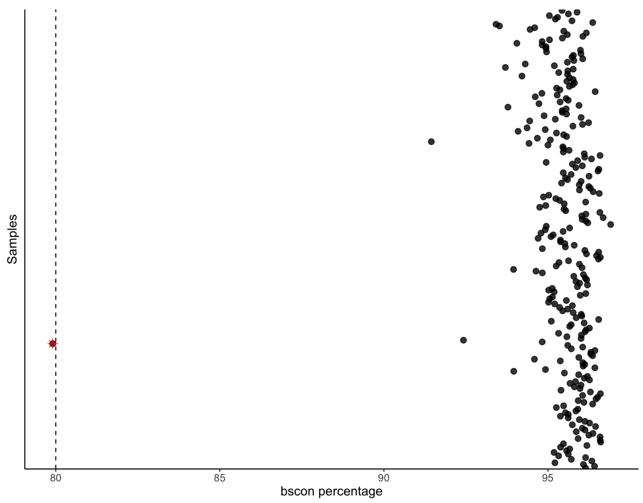


**QC figure 43:** bscon metric from the *wateRmelon* package, which measures bisulfite conversion efficiency [2].

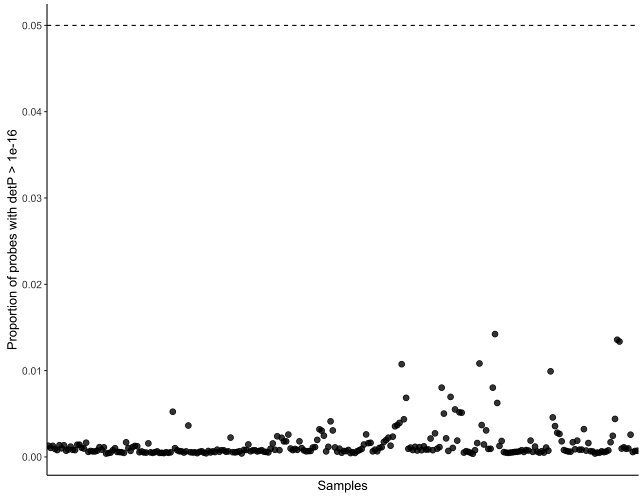


**QC figure 44:** Percentage of probes with detection P-value $> 1 \times 10^{-16}$

.

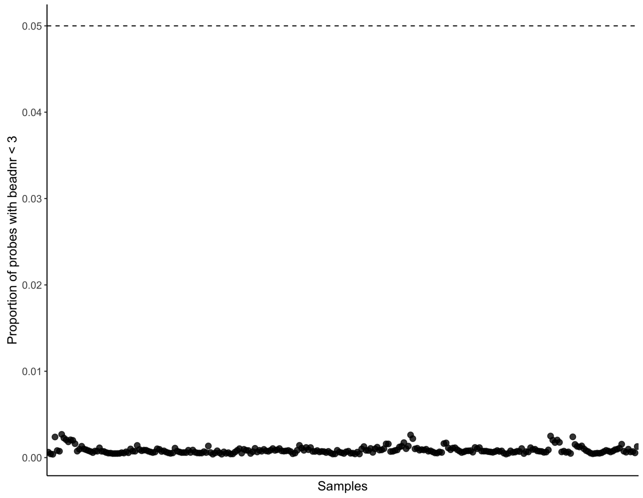


**QC figure 45:** Percentage of probes with beadNr $<$ 3.

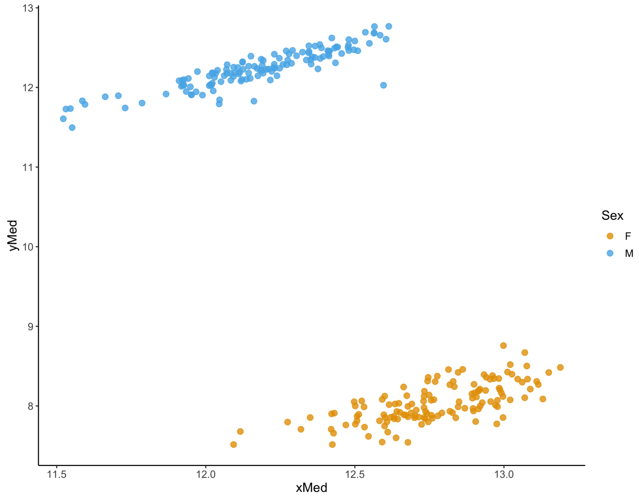


**QC figure 46:** Median X and Y copy number, as implemented in the *minfi* package [3].

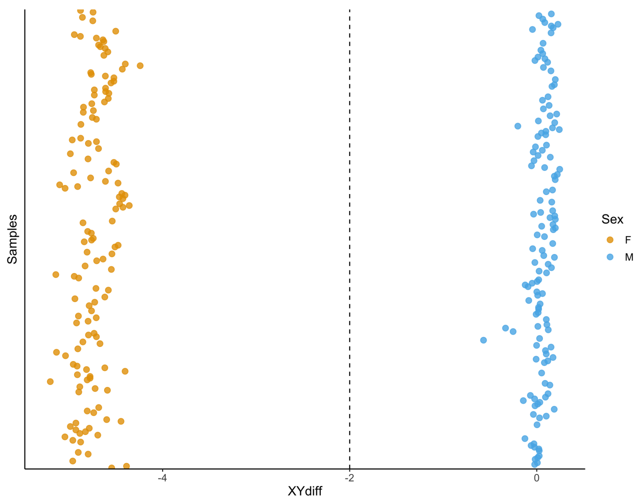


**QC figure 47:** Difference between median Y-chromosomal and median X-chromosomal signal intensity.

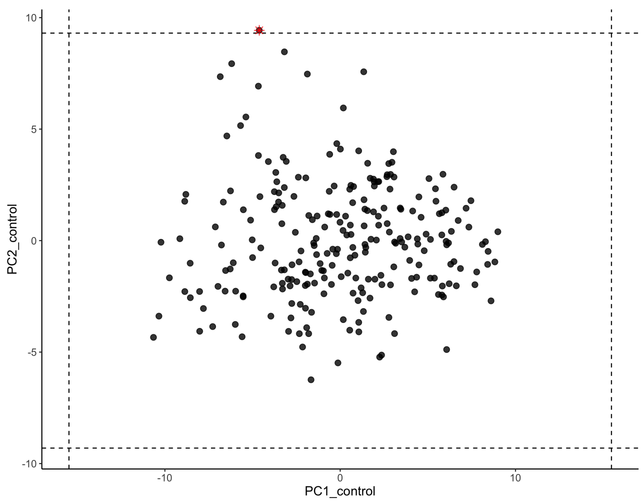


**QC figure 48:** Control probe principal components, after first round of QC.

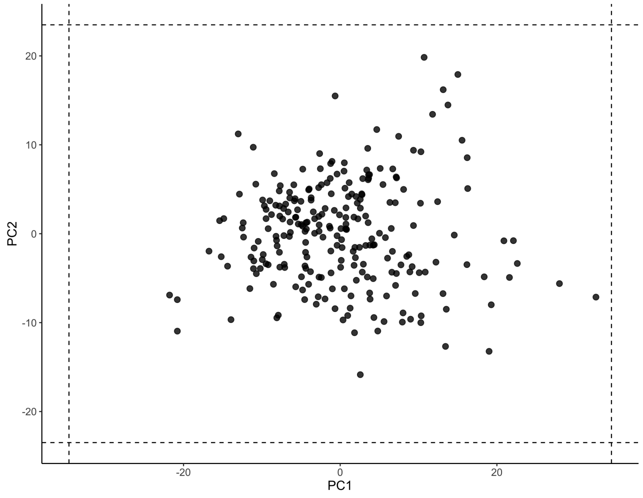


**QC figure 49:** Array-wide principal components, after first round of QC.

### 5 Definition of AUS1 and AUS2 strata

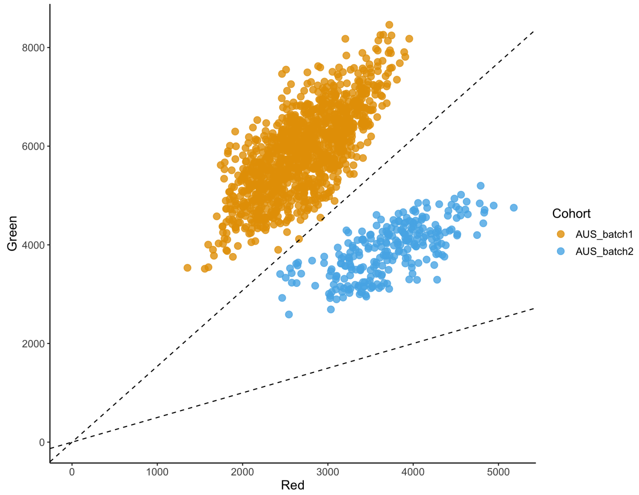


**QC figure 50:** Median red vs. median green signal in all Australian data, calculated in type I probes. AUS1 samples were defined based on an RG ratio <0.65, and AUS2 samples were defined based on an RG ratio >0.65.
